# Antibiotic Review Kit for Hospitals (ARK-Hospital): a stepped wedge cluster randomised controlled trial

**DOI:** 10.1101/2022.06.13.22275007

**Authors:** Martin J Llewelyn, Eric P Budgell, Magda Laskawiec-Szkonter, Elizabeth LA Cross, Rebecca Alexander, Stuart Bond, Phil Coles, Geraldine Conlon-Bingham, Samantha Dymond, Morgan Evans, Rosemary Fok, Kevin J Frost, Veronica Garcia-Arias, Stephen Glass, Cairine Gormley, Katherine Gray, Clare Hamson, David Harvey, Tim Hills, Shabnam Iyer, Alison Johnson, Nicola Jones, Parmjit Kang, Gloria Kiapi, Damien Mack, Charlotte Makanga, Damian Mawer, Bernie McCullagh, Mariyam Mirfenderesky, Ruth McEwen, Sath Nag, Aaron Nagar, John Northfield, Jean O’Driscoll, Amanda Pegden, Robert Porter, Neil Powell, David Price, Elizabeth Sheridan, Mandy Slatter, Bruce Stewart, Cassandra Watson, Immo Weichert, Katy Sivyer, Sarah Wordsworth, Jack Quaddy, Marta Santillo, Adele Krusche, Laurence SJ Roope, Fiona Mowbray, Kieran S Hand, Melissa Dobson, Derrick Crook, Louella Vaughan, Susan Hopkins, Lucy Yardley, Timothy EA Peto, Ann Sarah Walker, the ARK trial team

## Abstract

**Background:** Strategies to reduce antibiotic overuse in hospitals depend on clinicians taking decisions to stop unnecessary antibiotics. There is a lack of evidence on how support clinicians do this effectively. We evaluated a multifaceted behaviour change intervention (ARK) which aims to reduce antibiotic consumption in hospitals by increasing decisions to stop antibiotics at clinical review.

**Methods:** We performed a stepped-wedge, hospital-level, cluster-randomised controlled trial using computer-generated sequence randomisation of 39 acute hospitals to 7 calendar-time blocks (12/February/2018–01/July/2019). Co-primary outcomes were monthly antibiotic defined-daily-doses (DDD) per acute/medical admission (organisation-level, superiority) and all-cause 30-day mortality (patient-level, non-inferiority, margin 5%). Clusters were eligible if they admitted non-elective medical patients, could identify an intervention “champion” and provide pre-intervention data from February/2016. Sites were followed up for a minimum of 14 months. Intervention effects were assessed using interrupted time series analyses in each cluster. Overall effects were derived through random-effects meta-analysis, using meta-regression to assess heterogeneity in effects across prespecified factors. Trial registration was ISRCTN12674243.

**Findings:** Adjusted estimates showed a year-on-year reduction in antibiotic consumption (−4.8%, 95%CI: -9.1%,-0.2%, p=0.042) following the ARK intervention. Among 7,160,421 acute/medical admissions, we observed a -2.7% (95%CI: -5.7%,+0.3%, p=0.079) immediate and +3.0% (95%CI: - 0.1%,+6.2%, p=0.060) sustained change in adjusted 30-day mortality. This mortality trend was not related to the magnitude of antibiotic reduction achieved (Spearman’s ρ=0.011, p=0.949). Whilst 90-day mortality odds appeared to increase over time (+3.9%, 95%CI:+0.5%,+7.4%, p=0.023), this was not observed among admissions before COVID-19 onset (+3.2%, 95%CI:-1.5%,+8.2%, p=0.182). Length of hospital stay was unaffected.

**Interpretation:** The weak, inconsistent effects of the intervention on mortality are likely to be explained by the COVID-19 pandemic onset during the post-implementation phase. We conclude that the ARK-intervention resulted in sustained, safe reductions in hospital antibiotic use.

**Funding:** NIHR Programme Grants for Applied Research, RP-PG-0514-20015.

**Research in context:** *Evidence before this study:* Acutely ill patients often need to receive antibiotics before full diagnostic information is available. Consequently, reducing overuse of antibiotics in hospitals requires clinicians to review and where appropriate, stop unnecessary antibiotic prescriptions. Evidence-based tools to support clinicians stop unnecessary antibiotics do not exist. We searched PubMed, with no language or date restrictions, on 31/January/2022 for clinical studies focused on improving antibiotic use for hospitalised adults using the terms “anti-bacterial agents therapeutic use” AND “antibiotic stewardship”. Among the 427 studies found, the great majority were uncontrolled evaluations of different approaches to education, decision support and feedback. These included one before-after study, which found no impact of unsupported clinician-led prescription review. Three small, hospital-level cluster-randomised trials were identified. One evaluated different approaches to feedback, one compared different hospital specialties and one found intense feedback to be effective. All were small and none considered clinical outcomes or sustainability. There is a need for research to deliver proven interventions ready for implementation into practice.

*Added value of this study:* We evaluated a multifaceted “Antibiotic Review Kit” (ARK) intervention to support prescribers to appropriately stop antibiotics at clinical review. ARK comprises a prescription decision-aid supported by a brief online training tool, guidance on implementation (including regular data collection and feedback) and a patient information leaflet. We found that the intervention was associated with a sustained reduction in hospital-level antibiotic use overall and of oral and narrow-spectrum antibiotics specifically. Weak trends were observed for 30-day mortality in opposite directions for immediate and sustained impact. Although there was a sustained increase in 90-day mortality after the intervention, this was only seen when analyses included patients admitted after the start of the COVID-19 pandemic. Taken together we conclude that these mortality effects are unrelated to the intervention.

*Implications of all available evidence:* The ARK intervention is safe and effective in reducing antibiotic use among adult medical hospital admissions. The tools used are now freely available for adoption into practice.

## Introduction

The impact of antimicrobial resistance (AMR) on global public health is comparable to malaria and HIV, associated with an estimated 4.95 million deaths in 2019^1^. AMR places increased demands on healthcare systems, with substantial economic consequences.^2,3^ Human consumption of antibiotics is a major driver of AMR.^4^ Antibiotic use varies widely between and within healthcare systems, and greater antibiotic use drives resistance, both at a population-level and an individual patient-level.^5^ Despite this, there is no evidence that clinical outcomes are influenced by the wide organisational-level differences in antibiotic use that exist between acute hospitals in England.^6^

Antimicrobial stewardship (AMS) aims to minimise resistance selection by ensuring antibiotics are only prescribed when clinically indicated and that narrow-spectrum agents are used whenever appropriate.^7^ In primary care settings, AMS strategies that avoid or delay antibiotics are demonstrably safe and effective in controlling antibiotic overuse.^8^ In hospitals, the need to ensure patients with serious bacterial infections are treated promptly before a diagnosis is confirmed, means that ongoing review and revision of a patient’s need for antibiotic treatment is required to safely minimise unnecessary use. In England, this approach is set out in the Department of Health’s guidance “Start Smart, then Focus”^9^ which requires prescribers to review and revise antibiotic prescriptions at 48-72 hours. In the United States, the analogous term “antibiotic timeouts” is used, but revised Centers for Disease Control and Prevention (CDC) guidance in 2019^10^ prioritised pharmacist-led audit and feedback to prescribers, as evidence is lacking that prescriber-led reviews reduce overall consumption.^11^

After the introduction of “Start Smart, then Focus” in 2011, antibiotic consumption in English hospitals continued to rise year-on-year until the onset of the COVID-19 pandemic in 2020.^12^ This was against a background of falling consumption in primary care and despite financial incentives to reduce hospital prescribing, first through a Commissioning for Quality and Innovation (CQUIN) in 2016-18^13^ and then through its incorporation into the NHS Standard Contract for acute hospitals. Although high rates of prescription review were achieved,^14^ anecdotally, the great majority of review decisions were to adjust rather than stop antibiotics.

AMS interventions that enable better prescribing are more acceptable and effective than restrictive interventions, particularly if supported by audit and feedback^15^. In primary care, resources such as the UK’s TARGET toolkit help promote antibiotic stewardship^16^. In contrast, translation of hospital stewardship research into practice has been hampered by weaknesses in both the intervention design process, which is often not evidence-based, and the study designs used, which are usually not experimental and do not consider clinical outcomes.^17,18^

The Antibiotic Review Kit (ARK) Hospital programme set out to develop and evaluate a multifaceted behaviour change intervention to reduce antibiotic use by increasing the number of decisions to stop antibiotics at clinical review.^19^ The intervention comprised four elements: a novel prescribing decision aid, an online training tool supporting the use of the decision aid, guidance for implementing audit and feedback, and a patient leaflet.^20^ Following a feasibility evaluation at one acute NHS hospital,^21^ the intervention was evaluated at a further 39 hospitals (three pilot trial sites, 36 full trial sites) across all nations of the UK in a stepped-wedge cluster randomised controlled trial (ISRCTN12674243).^19^ A cluster design was essential as the intervention had to be implemented at the level of a healthcare organisation to avoid contamination. A stepped-wedge design was essential because of the limited number of secondary care organisations that could be randomised. Here we report the immediate and sustained impact of the intervention on hospital-level antibiotic consumption and patient-level clinical outcomes.

## Methods

### Objective

The primary objective was to evaluate whether the ARK-hospital intervention could safely reduce total antibiotic use in acute/general medical inpatients.

### Design

The intervention targeted healthcare professionals involved in antibiotic prescribing among patients admitted to acute/general medical specialties.^20^ It comprised: 1) a decision aid intended to be embedded in the prescription process which classified antibiotic prescriptions initially as either “possible risk from infection” or “probable risk of infection” and then “finalised” when a clear indication for ongoing antibiotic treatment was established at a 48-72h review; 2) online training to motivate and support use of the decision aid; 3) implementation guidance, including audit and feedback tools; and 4) a patient leaflet.^22^ All the tools developed within the ARK programme are freely available through the British Society for Antimicrobial Chemotherapy (BSAC) at: <u>antibioticreviewkit.org.uk</u>. Fidelity of intervention implementation was assessed using eight pre-defined criteria measuring staff engagement, uptake of the different intervention components, and timely submission of study data (**Table S1**).

The unit of observation was a hospital organisation, a single or group of hospital(s) with one executive board and governance framework (NHS Trusts in England; Health Boards in Northern Ireland, Wales and Scotland). The evaluation had three phases: a single-site feasibility study (phase I),^21^ a pilot study at three hospital organisations, and a stepped wedge cluster-randomised controlled trial at 36 hospital organisations. To be eligible, sites needed to admit adult general/medical inpatients, have a local ‘champion’ willing to lead intervention implementation, and be able to provide the required study data. Sites were randomised using a computer-generated list by the Trial Statistician (ASW) in seven blocks of six sites, to implement every 1-2 weeks excluding over holidays or where a pause on randomisation was requested by the funder (**Figure S1**). Allocation was concealed until the point of randomisation when sites were told by the Trial Statistician that their randomised implementation date was 12 weeks in the future. Ethical approval was obtained from the South Central Oxford C Research Ethics Committee (17/SC/ 0034) and the Confidentiality Advisory Group (17/CAG/0015) without individual patient consent since electronic health records were pseudonymised and no personal identifiable data was collected other than date of death.

### Co-primary outcomes

The trial had two co-primary outcomes: (1) antibiotic defined daily doses (DDDs) per acute/general medical admission (superiority), and (2) all-cause mortality within 30 days of admission (in/out of hospital) (non-inferiority). With a minimum of 36 organisations, the stepped-wedge cluster randomised design had >85% power to exclude a 5% relative increase in 30-day mortality and to detect a 15% relative reduction in antibiotic use (details in **Supplementary Methods**).^19^ As the intervention was consistent across sites in the pilot and main trial phases, following the approved protocol, the primary analysis included both to maximise power (n=39 organisations).

### Secondary outcomes

Secondary antibiotic (superiority) outcomes were total antibiotic DDDs per acute/general medical bed-day, and DDDs/admission for specific antibiotic groups (carbapenems, parenteral and oral, broad-spectrum and narrow-spectrum, and the UK Health Security Agency’s interpretations of Access, Watch, and Reserve (AWaRe) from the World Health Organisation’s Essential Medicines List^23^ (**Table S2**)). Admissions, rather than bed-days, were used as the main denominator for antibiotic use because bed-days may be influenced by non-medical reasons for prolonged hospital stays. Secondary non-inferiority outcomes were 90-day mortality, admission to an intensive care unit (ICU), length-of-stay, emergency hospital readmission (to any speciality) within 30 days of discharge and *Clostridioides difficile* infection or colonisation.

All outcomes were assessed using pseudonymised electronic health records from acute/general medical admissions (denoted “spells”, see **Supplementary Methods**) and consultant care episodes, bulk antibiotic dispensing on the wards that implemented the ARK intervention (as most organisations did not have electronic individual patient-level prescribing systems), and *C. difficile* test results. Date of death within 90 days of admission (in/out of hospital) was obtained by sites through linkage with the national registries. Since the intervention targeted acute/general medical wards, the study population was defined using the consultant specialty codes most often used to admit adult general medicine inpatients (**Figure S2;** rationale in^19^). Data were requested from February 2016 to October 2020 inclusive to span from two years before the main trial started to 15 months after the last site implemented, to ensure sufficient data were available to estimate pre and post-implementation trends (included time periods for co-primary outcomes in **Figure S1**). The randomised date of implementation was used as the time of implementation in an intention-to-treat analysis, modified following the Statistical Analysis Plan to exclude 7 sites that withdrew after randomisation but before implementation. As mortality was a non-inferiority comparison, it was more important to replace these sites than use resources collecting data from sites that never implemented the intervention and hence would show no effect of the (not implemented) intervention on mortality. See **Supplementary Methods** for details, including data cleaning steps (shown for 30-day mortality in **Figure S3**).

### Statistical analysis

An interrupted time series analysis estimated the intervention’s immediate impact (‘step change’) and sustained impact on year-on-year trends post versus pre-implementation in each hospital organisation. Overall intervention effects were then obtained using random effects meta-analysis, using meta-regression to assess heterogeneity in effects across prespecified factors. Monthly antibiotic DDDs per admission were modelled using negative binomial regression, and binary outcomes per admission using logistic regression. Length-of-stay (days) was modelled using subhazard regression, treating inpatient deaths as a competing risk and censoring at 90 days, using 0.1 days for those admitted and discharged on the same day, as was emergency 30-day readmission in a sensitivity analysis, with out-of-hospital deaths as the competing event. Due to low event rates (<4% in all sites), sensitivity analyses did not model ICU admission and *C. difficile* infection/colonisation using subhazard regression. All models included a robust variance adjustment by patient.

Since the COVID-19 pandemic profoundly affected both the trial’s primary outcomes, all models included a binary indicator for March-June 2020 unless otherwise noted. Sensitivity analyses dropped admissions after February 2020, and 12 sites with <12 months post-implementation data as a result. Antibiotic models additionally adjusted for seasonal effects by including day of year as a sin() + cos() function to ensure a smooth transition in risk from year to year. Non-antibiotic models also adjusted for individual admission-level covariates, regardless of statistical significance (based on^24^), including: sex, age, immunosuppression, deprivation percentile, Charlson comorbidity index and its interaction with age, admission method, admission source, admission specialty, patient classification, admission day of the week (weekend vs weekday), admission day of year and time of day (and its interaction with admission day of the week), and number of overnight admissions and any previous overnight complex (>1 consultant episode, excluding episodes in the emergency department and rehabilitation) admission in the past year. Ethnicity was missing for a median 8.8% of spells (IQR: 4.5-18.4%) per site so was not adjusted for. See **Supplementary Methods** for further details.

All analyses used Stata/MP 17. The Data Monitoring Committee reviewed outcome data three times during the trial, using a Haybittle-Peto statistical rule for early stopping.

## Results

Three pilot sites implemented the intervention between 25 September 2017 and 20 November 2017; 36 further sites were randomised to implement the intervention between 12 February 2018 and 1 July 2019. Sites were informed 12 weeks ahead of their randomised implementation date to allow for implementation preparation.

Thirteen sites were classed as large (>850 beds available, median: 991), 14 medium (551-850 beds, median: 670) and 12 small (≤550 beds, median: 487); 13 were in the South of England, 10 in the North, 6 in the Midlands and East of England, 3 in London, 4 in Northern Ireland, 2 in Wales, and 1 in Scotland. The Champion was a Microbiologist in 19 sites, was trained jointly in Microbiology and Acute Medicine or Infectious Diseases in 3, in Acute Medicine alone in 7, and was a Pharmacist in 10. At implementation, prescribing was done on paper in 25 sites and electronically in 14 (6 Cerner™, 3 JAC™, 5 other). Twenty-one (54%) sites implemented the decision aid with a “hard stop” to the initial prescription unless revised by 72 hours, 9 (23%) as a “soft stop” highlighting the need to stop or finalise within 72 hours, and 9 (23%) did neither.

Antibiotic use in the 12 months before randomised implementation varied very widely both in terms of total DDDs/admission (median: 2.9, IQR: 2.1-4.7, range: 0.4-11.3) and specific agents, classes, and AWaRe categories (**Figure S4**). Access antibiotics accounted for 30.7-85.2% of total DDDs, Watch for 4.8-44.1%, and Reserve for 0.3-5.7%.

### Adherence to intervention

Site champions named a median 19 (IQR: 14-34, range: 5-72) people as essential for doing the online training; median 78% (IQR: 63-90%) completed the training by 12 weeks, with 16 (41%) sites below the ≥70% target (**Figure 1A**). The total number of staff completing training also varied substantially by site size, with median 28 (IQR: 20-44) staff trained per 100 acute beds and 9 (22%) sites achieving <20 (**Figure 1B&C**). Actual implementation was delayed at 9 sites, by median 7.4 weeks after the randomised date (IQR: 6.1-13, range: 5.3-25), typically because of delays implementing the decision aid into the prescribing processes. Sites achieved a median 6 (IQR: 5-7, range: 2-8) of the eight implementation fidelity criteria (**Table S1**); 9 (23%) sites achieved 4 or fewer.

**Figure 1:**
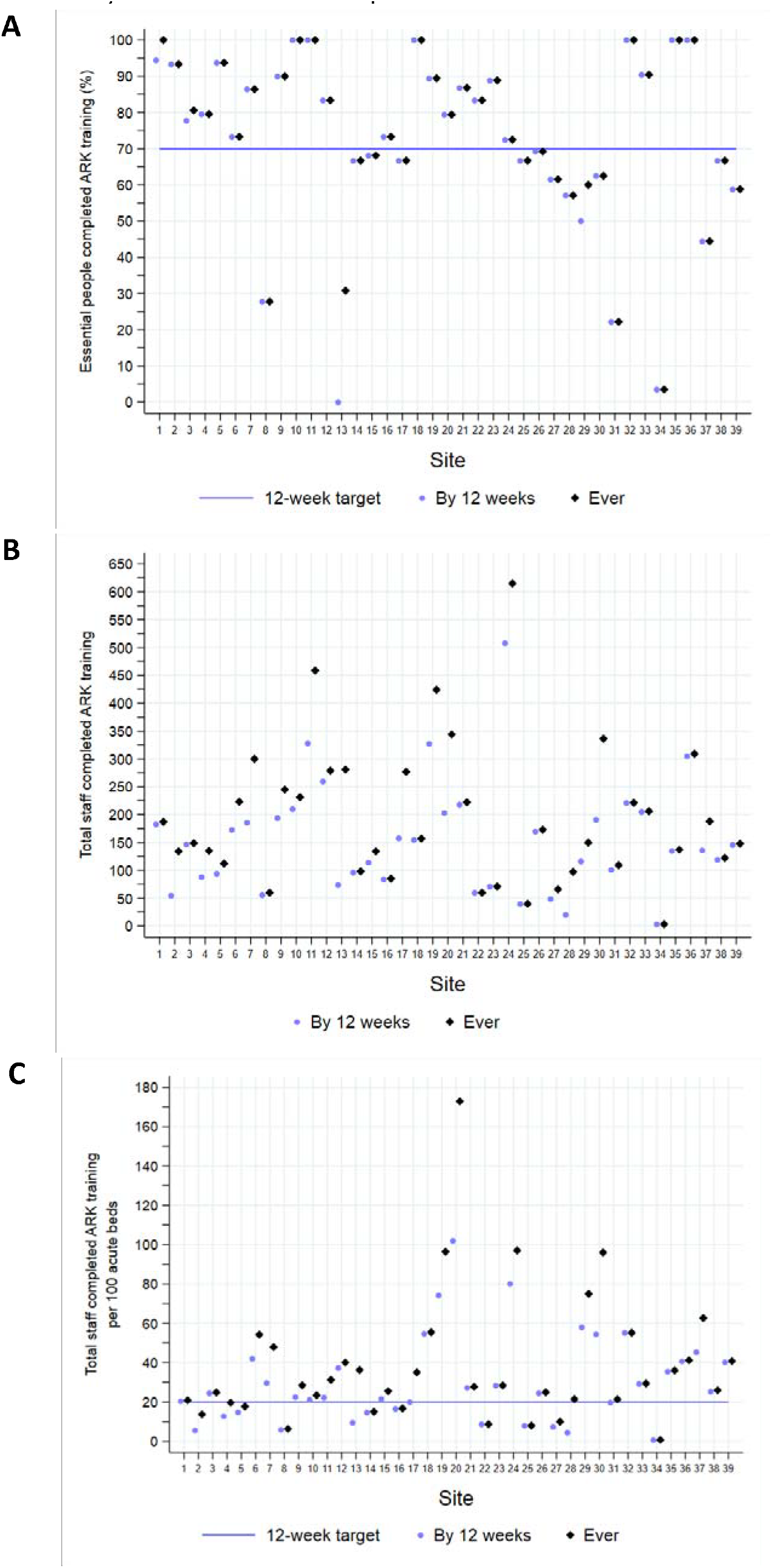
Intervention adherence by the end of the 12-week implementation phase and ever: Percentage of essential people completing ARK training (A), staff completing ARK training (B) and total staff completing training per 100 acute beds (C). Sites are identified numerically by the order in which they were randomised to implement.

Post-implementation audit data were available for 37 sites, of which 31 provided baseline audit data (**Figure 2**). In the 12 weeks following randomised implementation, across sites a median 51.6% (IQR: 31.4-75.9%) of regularly audited antibiotic prescriptions were categorised using the decision aid at the initial prescription. At 12 weeks a median 89.9% (IQR: 80.8-96.5%) of audited prescriptions were reviewed versus 91.0% (IQR: 78.6-95.8%) at baseline (Wilcoxon matched-pairs p=0.209), and a median 16.2% (IQR: 12.8-23.3%) were stopped at ‘review and revise’ versus 12.7% (IQR: 5.4-21.4%) at baseline (p=0.006).

**Figure 2:**
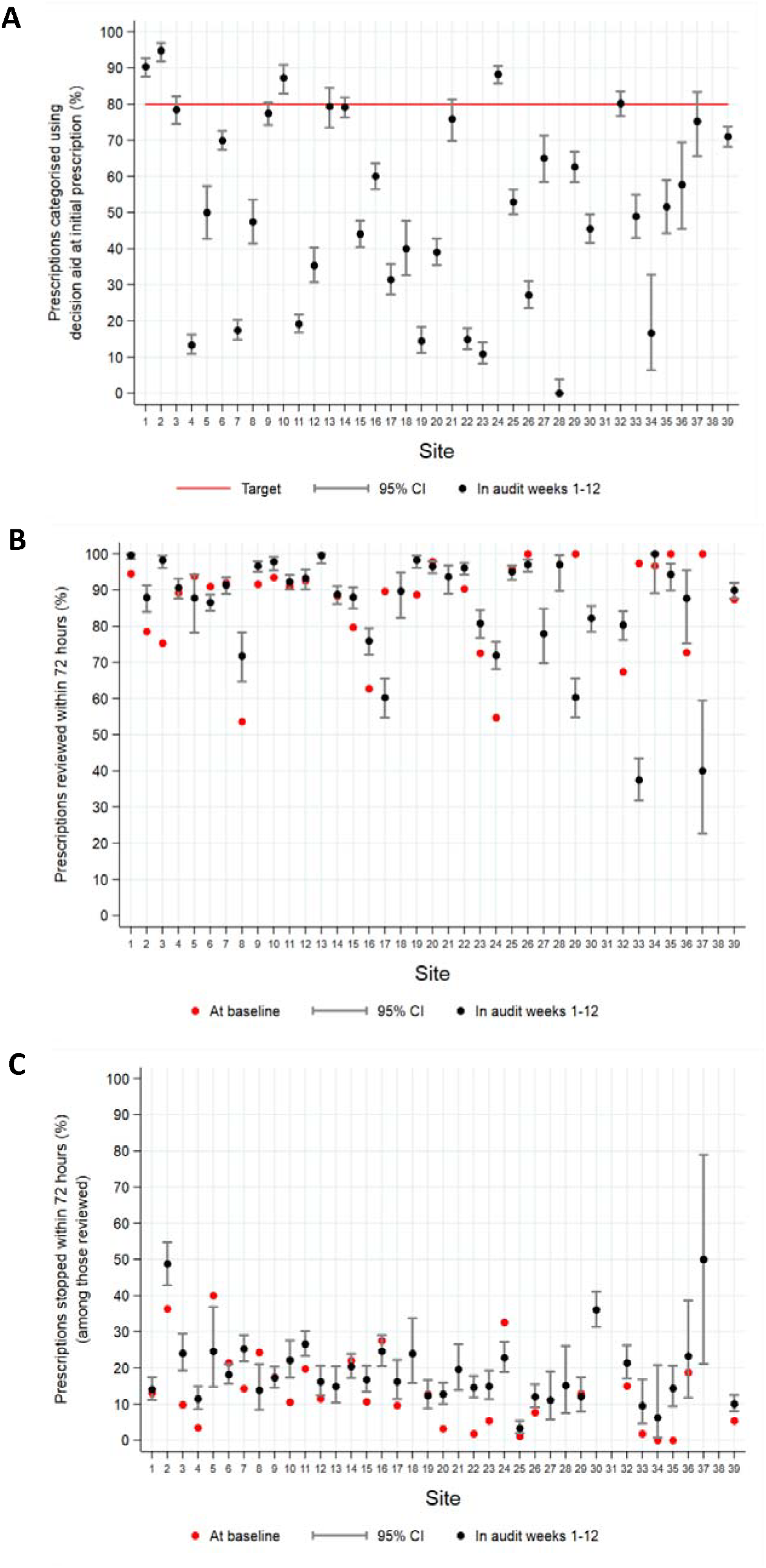
Audits of antibiotic prescriptions in the 12 weeks following randomised implementation versus baseline; percent categorised using the decision aid at the initial prescription (A), percent reviewed (B), and percent stopped at ‘review and revise’ (C)

### Total DDD per acute/general medical admission (co-primary outcome)

Sites contributed a median 23 months (range: 14-37) of antibiotic data post-implementation (**Figure S1**). In the final model, adjusted for the impact of COVID-19 as well as standard interrupted time series trends (shown by site in Figure S5), the intervention was associated with a -1.0% immediate change in total antibiotic DDDs/admission (95% CI: -4.0%,+2.1%, p=0.540) and a sustained -4.8% reduction in year-on-year trends post vs pre-implementation (95% CI: -9.1%,-0.2%, p=0.042) (**Figures 3&4**), with little association between the immediate and longer-term intervention effects across sites (Spearman’s rho: -0.088, p=0.599, **Figure S6**). There was substantial heterogeneity in trajectories of DDDs/admission pre and post-intervention (**Figure S7**). Intervention effects were similar unadjusted (**Figure 3**) and excluding all follow-up from March 2020 (**Table S3**).

**Figure 3:**
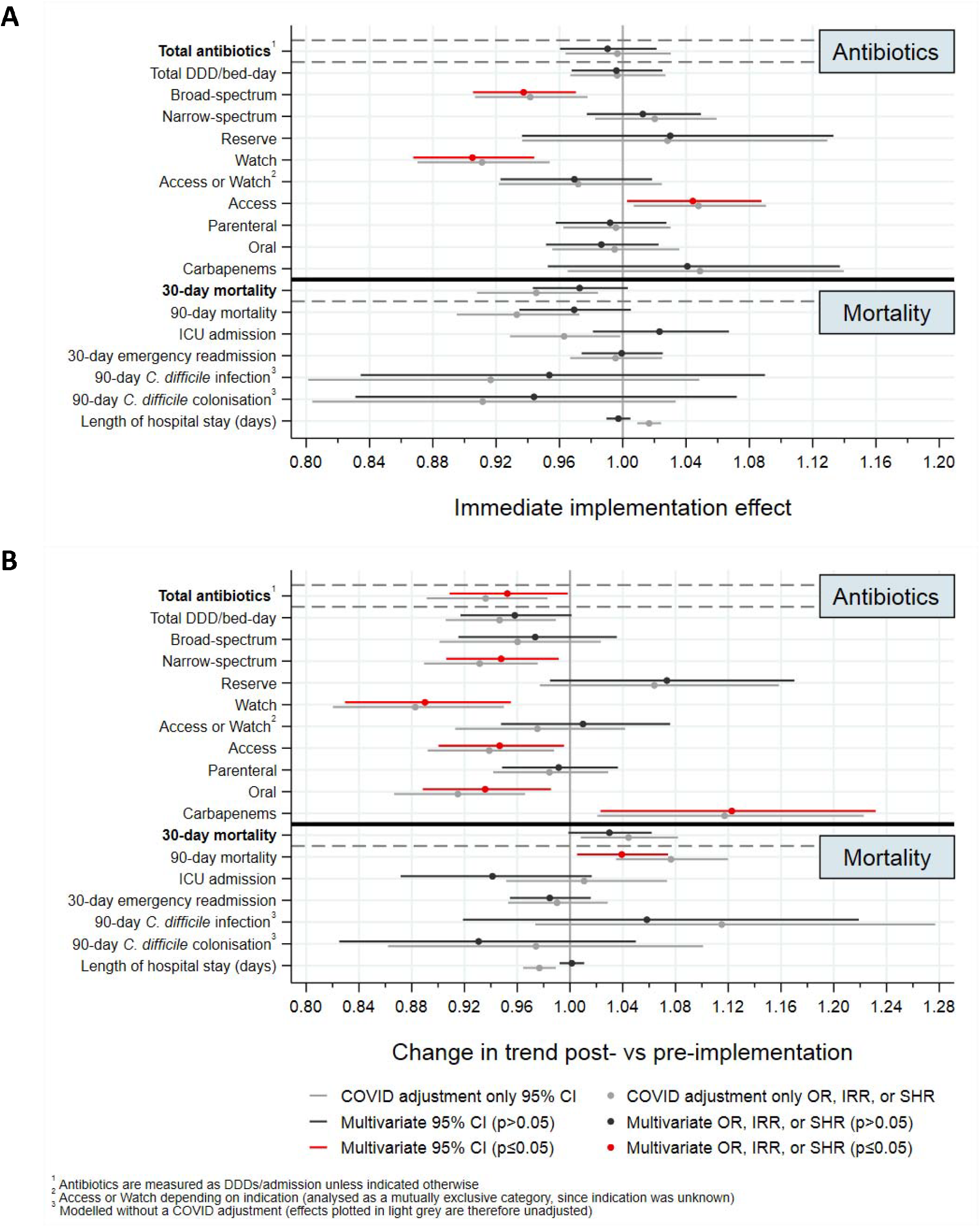
Impact of the ARK intervention: immediate effect at implementation (A) and effect on year-on-year trend post-vs pre-implementation (B). top part of each panel shows antibiotic primary (bold) and secondary outcomes, and bottom part shows clinical primary (bold) and secondary outcomes. Effects only adjusted for the effects of COVID shown in grey, and fully adjusted effects (see Methods) in red where evidence of an association or otherwise in black. OR = Odds ratio (logistic regression), IRR = Incidence rate ratio (negative binomial regression), SHR = subhazard ratio (competing risks regression)

There was no evidence that immediate effects at implementation and on year-on-year trends post vs pre-intervention were associated with overall implementation fidelity (per unit higher -0.5%, 95% CI: -2.7%,+1.7%, p=0.640; and -1.4%, 95% CI: -4.9%,+2.3%, p=0.448, respectively) (**Figure 4**). There was evidence of greater reductions in total DDDs/admission at implementation among sites with a process in place for ongoing audit and feedback by the implementation date (incidence rate ratio (IRR): -16.6%, 95% CI: -28.5%,-2.8%, heterogeneity p=0.022), and among sites that submitted post-implementation audit data within 4 weeks following implementation (IRR: -8.3%, 95% CI: -15.1%,- 1.0%, heterogeneity p=0.027), with the latter explained by the former in multivariable models (**Table S4**). There was also a trend towards greater reductions in the year-on-year trends in total DDDs/admission post vs pre-implementation among sites that introduced the ARK categories into the prescribing process by the implementation date (by -11.5%, 95% CI: -22.9%,+1.7%, heterogeneity p=0.083) and among sites with greater uptake of the online learning (≥20 people per 100 acute beds) by the implementation date (by -9.9%, 95% CI: -19.7%,+1.1%, heterogeneity p=0.075). Medium-sized sites also tended to have greater reductions in DDDs at implementation (heterogeneity p=0.064), but then less year-on year reduction subsequently (heterogeneity p=0.049) (**Table S4**).

**Figure 4:**
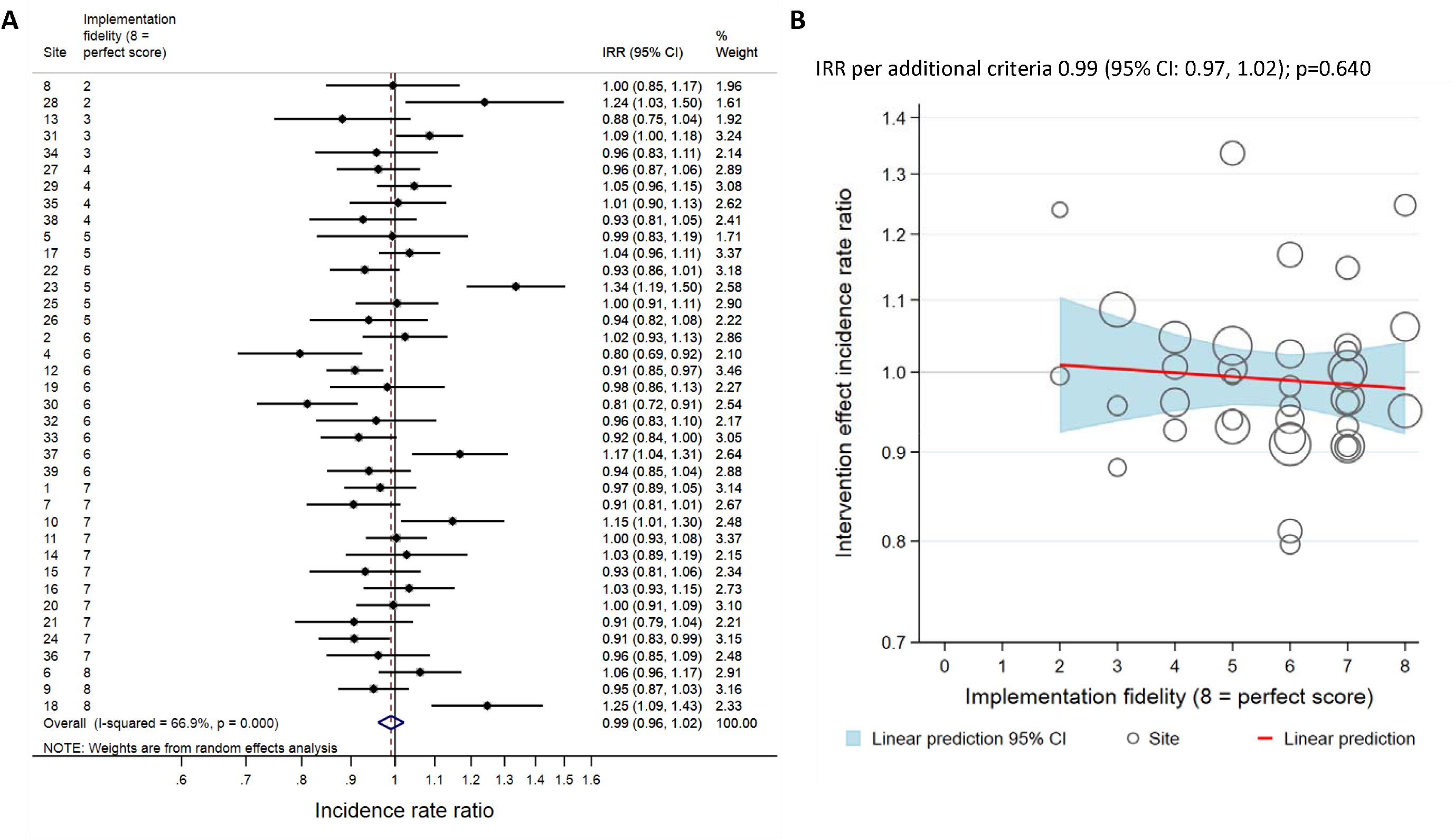
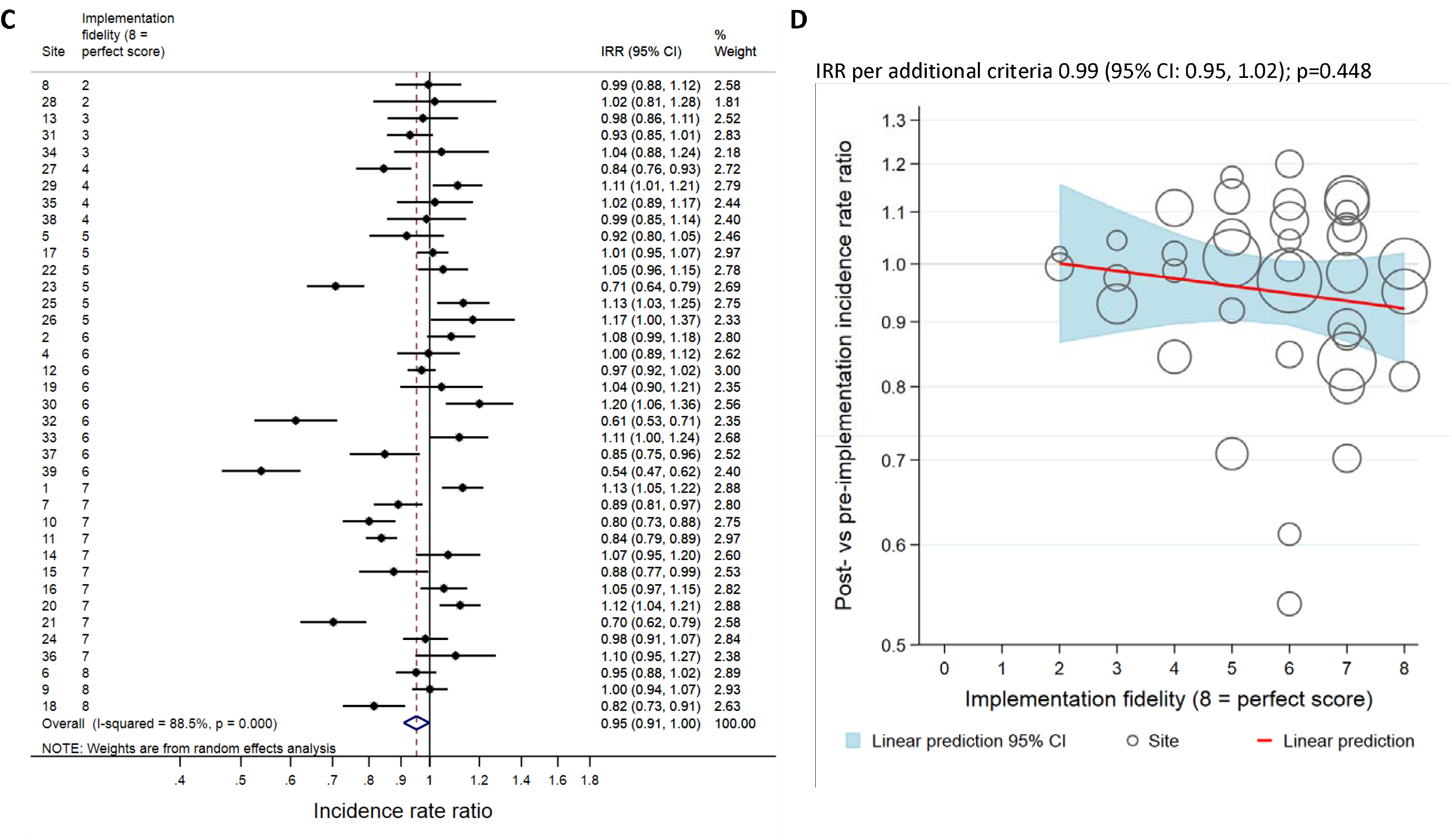
Total antibiotic DDDs/admission (co-primary endpoint) by site; immediate effect at implementation, overall (A) and by implementation fidelity (B), and effect on year-on-year trend post-vs pre-implementation, overall (C) and by implementation fidelity (D). Sites identified numerically by the order in which they were randomised to implement, and ordered by the number of fidelity criteria achieved (Table S1). IRR = incidence rate ratio.

### Secondary antibiotic outcomes

Adjusted models showed no evidence of an immediate impact of the ARK intervention on total antibiotic DDDs/bed-day (−0.4%, 95% CI: -3.2%,+2.5%, p=0.784), with a trend towards reductions in year-on-year trend subsequently (by -4.2%, 95% CI: -8.3%,+0.1%, p=0.056) (**Figures 3, S8**); similar to effects on total DDD/admission (co-primary outcome). At implementation, rates of broad-spectrum and Watch DDDs/admission dropped significantly (−6.3%, 95% CI: -9.5%,-3.0%, p=0.0003, and -9.5%, 95% CI: -13.2%,-5.6%, p<0.0001, respectively) whereas Access DDDs/admission increased slightly (+4.4%, 95% CI: +0.3%,+8.8%, p=0.037) (**Figures 3, S9, S10**). There was no evidence of an effect of the ARK intervention on the other secondary antibiotic outcomes at implementation (**Figure 3**).

However, there were sustained reductions in the year-on-year trend for most antibiotic DDD/admission groups, including for narrow-spectrum (−5.2%, 95% CI: -9.4%,-0.9%, p=0.019) (**Figures 3, S9**), Watch (−11.0%, 95% CI: -17.1%, -4.5%, p=0.001), Access (−5.3%, 95% CI: -10.0%,-0.4%, p=0.033) (**Figure S10**), and oral antibiotics (−6.4%, 95% CI: -11.2%,-1.4%, p=0.012) (**Figure S11**). There was no evidence of longer term effects on broad-spectrum (−2.6%, 95% CI: -8.5%,+3.6%, p=0.395), parenteral (−0.9%, 95% CI: -5.2%,+3.6%, p=0.703), or antibiotics considered Access or Watch depending on indication (+1.0, 95% CI: -5.2%,+7.6%, p=0.762). In contrast, year-on-year trends in DDDs/admissions increased faster post vs pre-implementation for carbapenems (+12.3, 95% CI: +2.3%,+23.2%, p=0.015) (**Figure S12**) with a similar trend for Reserve antibiotics (+7.3%, 95% CI: -1.5%,+17.0%, p=0.107) (**Figure S10**).

### 30-day mortality (co-primary outcome)

Sites contributed a median 23 months (range: 15-37) 30-day mortality data post-implementation (**Figure S1**). Analysis of all-cause 30-day mortality (in/out of hospital) included 7,160,421 admissions in 39 sites (**Table S5**), of which 314,313 (4.4%) died within 30 days of admission (2.6-7.2% across sites, median: 4.6%, IQR: 4.0-5.0). Overall, the ARK intervention was associated with a -2.7% (95% CI: -5.7%,+0.3%, p=0.079) immediate change in the odds of 30-day mortality and a +3.0% (95% CI: - 0.1%,+6.2%, p=0.060) change in year-on-year trends post vs pre-implementation (**Figures 3&5**). Sites with larger immediate mortality reductions tended to have larger increases in year-on-year trends post vs pre-implementation (Spearman rho: -0.28, p=0.082, **Figure S13**). This indicates the weak overall effects could be an artefact from the individual interrupted time series (**Figure S5**), given the substantial heterogeneity in trajectories of 30-day mortality pre and post-intervention (Figure S14), potentially related to the electronic data submitted (**Table S6**). There was no evidence that effects on 30-day mortality were associated with implementation fidelity (immediate effect +1.3% per unit higher, 95% CI: -6.8%,+3.3%, p=0.194; and change in year-on-year trend post vs pre-implementation - 0.1%, 95% CI: -2.1%,+1.9%, p=0.926, respectively) (**Figure 5**). Intervention effects on crude 30-day mortality were relatively similar (**Figure 3**), as were effects excluding all follow-up from March 2020 (**Table S3**).

**Figure 5:**
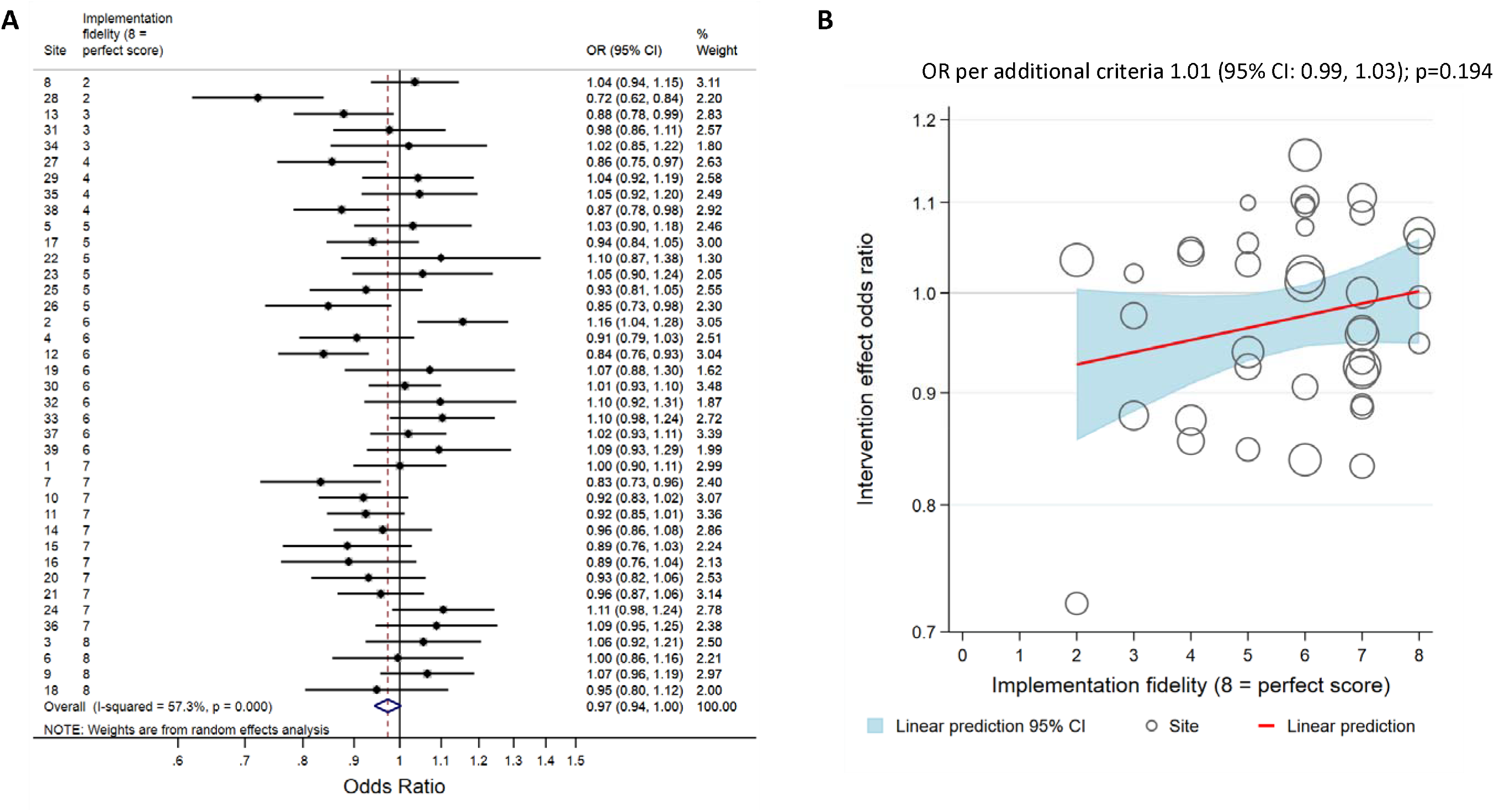
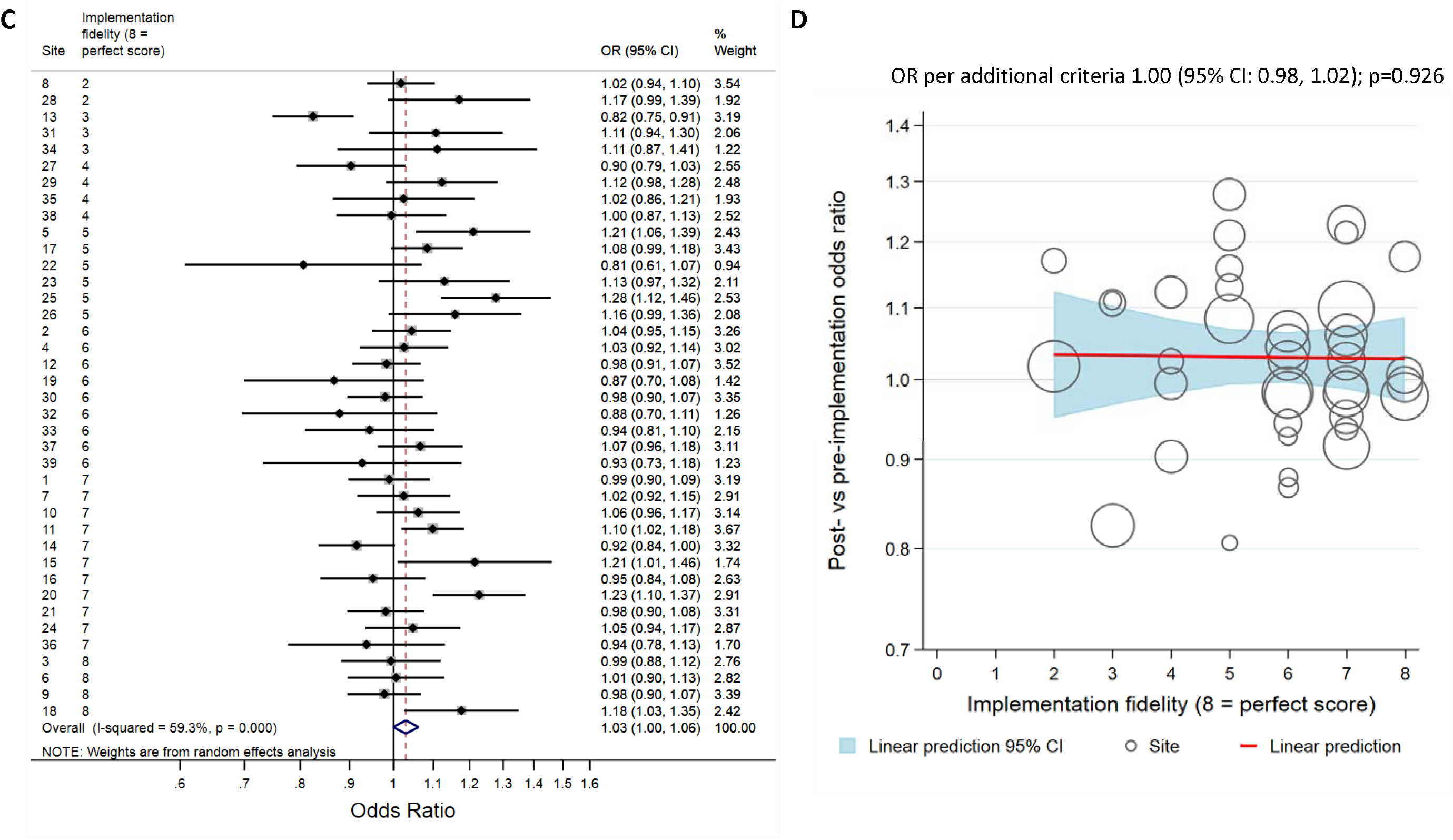
Adjusted 30-day mortality (co-primary endpoint) by site; immediate effect at implementation, overall (A) and by implementation fidelity (B), and effect on year-on-year trend post vs pre implementation, overall (C) and by implementation fidelity (D). Sites identified numerically by the order in which they were randomised to implement, and ordered by the number of fidelity criteria achieved (**Table S1)**. OR = Odds ratio.

Meta-regression results provided weak evidence for greater reductions in the year-on-year trends in 30-day mortality among sites that introduced the ARK categories into the prescribing process by the implementation date (−7.2%, 95% CI: -14.6%,+0.8%, p=0.075), among sites implementing a hard stop versus a soft stop/neither (−7.1%, 95% CI: -12.7%,-1.2%, p=0.020), and among sites implementing in July-September versus January-March (−10.3%, 95% CI: -19.0%,-0.8%, p=0.036) (**Table S4**).

There was no evidence that sites with greater reductions in antibiotic DDDs/admission had larger increases in 30-day mortality for either the immediate effect or the effect on year-on-year trends post vs pre-implementation (**Figure 6**).

**Figure 6:**
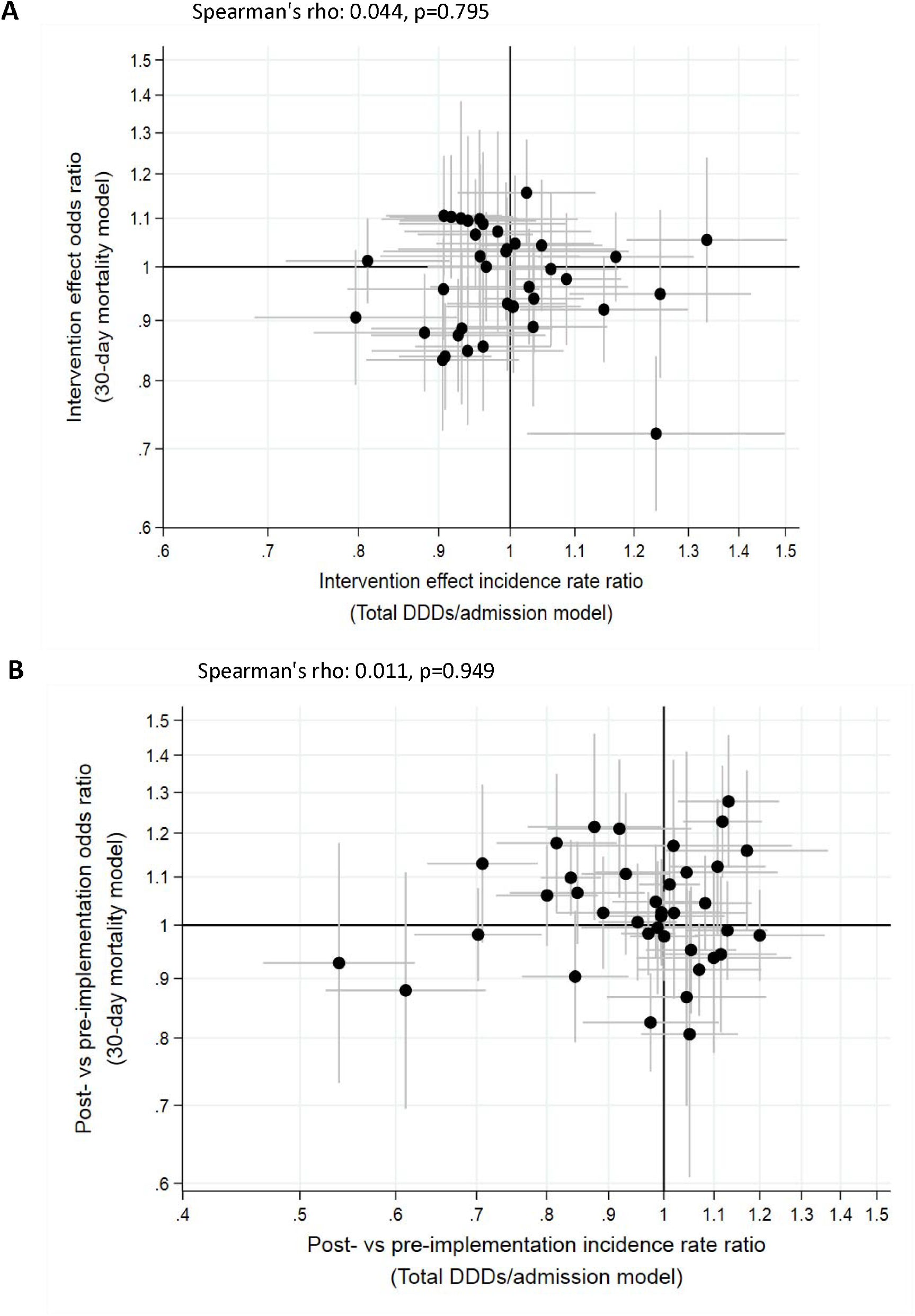
Comparison of intervention effects on 30-day mortality and total antibiotic DDDs/admission, immediate effect at implementation (A) and effect on year-on-year trend post-vs pre-implementation (B)

### Other secondary clinical outcomes

Mortality within 90 days of admission was 8.1% overall (range: 4.6%,12.5% by site). Adjusted models showed the ARK intervention was associated with a -3.1% (95% CI: -6.5%,+0.5%, p=0.092) immediate change in 90-day mortality odds and a +3.9% (95% CI: +0.5%,+7.4%, p=0.023) increase year-on-year post vs pre-implementation (**Figure S15**). However, there was no evidence of associations after excluding admissions from the onset of the COVID pandemic in March 2020 (−3.9%, 95% CI: - 8.3%,+0.7%, p=0.098, and +3.2%, 95% CI: -1.5%,+8.2%, p=0.182, respectively) (**Table S3**).

Admission to critical care was 1.6% overall (range: 0.4%,4.2%). There was no evidence of association with ICU admission at implementation (+2.3%, 95% CI: -1.9%,+6.7%, p=0.284) or in year-on-year trends post vs pre-implementation (−5.9%, 95% CI: -12.8%,+1.6%, p=0.123) (**Figure S16**). Median length-of-stay was 8.5 hours across sites (IQR: 3.1-89.2). There was no evidence of association between length-of-stay and the ARK intervention at implementation (−0.3% relative change in subhazard ratio, 95% CI: -1.0%,+0.5%, p=0.480) or post vs pre-implementation (+0.1%, 95% CI: - 0.8%,1.1%, p=0.766) (**Figure S17**). Emergency readmission to hospital (any speciality) was 13.6% across sites (range: 8.7%,26.4%), with no evidence of association with the ARK intervention at implementation (−0.1%, 95% CI: -2.6%,+2.5%, p=0.957) or post vs pre-implementation (−1.5%, 95% CI: -4.6%,+1.6%, p=0.330) (**Figure S18;** sensitivity analysis using competing risks similar). Detection of *C. difficile* infection within 90 days of admission was low (0.2% overall, range: 0.1%,0.6%), as was *C. difficile* colonisation exclusive of infection (0.5% overall, range: 0.2%,1.1%). The odds of *C. difficile* infection and colonisation (exclusive of infection) within 90 days of admission were not associated with the ARK intervention either at implementation (−4.6%, 95% CI: -16.6%,+9.0%, p=0.486; and - 5.6%, 95% CI; -16.9%,+7.2%, p=0.374, respectively) or post vs pre-implementation (+5.8%, 95% CI: - 8.1%,+21.9%, p=0.432; and -6.9%, 95% CI: -17.5%,5.0%, p=0.243, respectively) (**Figure S19**).

## Discussion

Here, we have evaluated the ARK intervention,^20,21^ which aims to safely reduce antibiotic consumption in adult acute/medical hospital admissions, in a stepped-wedge cluster-randomised trial.

In our final model adjusting for 14 admission factors and COVID-19, we found that the intervention resulted in average reductions in antibiotic use over time of 4.8% per year. That the intervention changed prescribing over time rather than suddenly is expected, given the different components, including training in the use of the novel decision aid, and the importance of audit and feedback re-enforcing learning.^25^ It may also reflect increasing acceptance of stopping antibiotic courses as concerns about not completing courses reduce^26^. Although the trial was powered to detect a 15% immediate reduction associated with the intervention, the impact observed is potentially clinically very significant given that the national Standard Contract for acute trusts in England currently sets out a requirement to reduce antibiotic consumption by 1% per annum. Given the importance of sustainable impact from behaviour change interventions in antibiotic stewardship, it is notable that this reduction was seen over a median of 23 (range: 14-37) months. Of note, consistent reductions were seen in Access and Watch and oral antibiotics but not in broad-spectrum or Reserve classes. Since the intervention was targeted at acute/general medical admissions it is not surprising its impact was seen in narrow-spectrum, Access agents which are typically used as first-line antibiotics or de-escalation choices. The significant increase in carbapenem use post-intervention could suggest a “squeezing the balloon” effect in which reduced use of one set of agents increases use of another. However, it is important to note that the differences measured are relative and carbapenems account for a tiny fraction of all hospital antibiotic consumption. Furthermore, their use is increasing disproportionately across the NHS, and we may simply be observing an increase that the intervention would not be expected to affect.

We found no clear relationship between the overall fidelity of implementation and the intervention’s impact. This may be because there are complex interactions between intervention elements and the implementation setting that are difficult to measure quantitatively in a large-scale trial. The ARK audit tools were designed to support frequent, light touch feedback to prescribers, sometimes called “hand-shake stewardship”^27^ which is dependent on interpersonal factors that we could not analyse here, but will be considered in forthcoming mixed methods process analyses. It is noteworthy that among individual intervention components, implementation of the decision aid into the prescribing process and greater uptake of the online learning were both linked to greater reductions in antibiotic use over time, suggesting that these are key elements in achieving a sustained change.

Hospital stewardship policies and the ARK intervention focus on decisions to continue rather than decisions to start antibiotics because this approach has the potential to reduce overall use without withholding empirical antibiotics from acutely ill patients. Nevertheless, we considered it important to evaluate whether introduction of ARK was associated with excess mortality. The onset of the COVID-19 pandemic in March 2020, when 12/39 sites were still within 12 months of implementation, was associated with substantial increases in mortality among acute hospital admissions throughout the NHS (**Figure S5**). Adjusting for this effect, both in the main models and through sensitivity analysis excluding these 12 sites, we found no clear evidence of an association between the intervention and 30-day or 90-day mortality, with weak trends towards decreased risk of death at intervention implementation and towards increased risk of death over time, likely still reflecting some residual confounding from COVID-19. Notably, implementation of the decision aid with a “hard stop” of antibiotic prescriptions at 72 hours if not revised, was associated with decreased risk of death over time. This is intriguing given that, anecdotally, in the trial clinicians reported anxiety that “hard stops” could compromise clinical outcomes.^28^ It could be explained by clinicians placing a greater emphasis on prescription reviews at sites that introduced “hard stops”, improving patient management more broadly. Furthermore, we found no evidence that sites that achieved greater reductions in antibiotic DDDs/admission had larger increases in mortality (**Figure 6**).

Our study has important limitations. First, there are intrinsic limitations of the cluster randomised design. Although we included one quarter of all acute hospitals in the UK health system, randomising 36 sites cannot reliably exclude imbalance, particularly of time-dependent factors, as highlighted by the onset of the COVID-19 pandemic during the post-implementation period. We cannot exclude the possibility of imbalance in other time-dependent organisational changes (e.g. staffing, practice, case-mix) which may have changed antibiotic consumption at individual sites. Second, although our sites were robustly randomised with respect to the timing of intervention implementation, they may not be a random sample of acute hospitals in the UK. For example, it is likely that only sites with well constituted antimicrobial stewardship teams came forward to participate and we cannot be certain that other sites would experience the same impact, particularly as impact was associated with some aspects of intervention fidelity. Alternatively, the intervention impact could be greater at sites with weaker stewardship teams. Third, the electronic data used to assess study outcomes has limitations. We measured antibiotic consumption indirectly from dispensing data to clinical areas but not all prescribing decisions within a particular area will have been subject to the intervention (e.g. outlying surgical patients). Similarly, as acute/general medical inpatients are not identified in electronic admission data, we had to infer this population from specialty codes which are used in slightly different ways in different organisations; this will have included in the analysis some patients for whom prescribing decisions were not subject to the intervention. Importantly, both these effects would be expected to dilute the observed impact of the intervention on antibiotic use compared to any true effect. Analysing routinely available electronic health record data, we found no consistent evidence of impact on mortality, admission to critical care, length-of-stay or readmission, but we cannot exclude the possibility of other harms related to shorter antibiotic treatment. Conversely, we have not been able to measure potential direct benefits from reduced antibiotic treatment.

Notwithstanding these limitations, the cluster randomised approach we adopted allowed us to capture both the organisation-level impact of the intervention on antibiotic consumption and patient-level impact on clinical outcomes. Our findings are entirely consistent with the three, much smaller, previous trials of hospital stewardship interventions, which demonstrated the importance of intervention co-design with practitioners^29^, practitioner education and clinically relevant audit and feedback to clinicians^30,31^. Our approach to intervention design and evaluation addresses many of the limitations that have prevented the translation of previous research findings into hospital practice.^17,18^ Crucially, the wider ARK-hospital programme has delivered practice-ready materials for implementation which are freely available through BSAC at <u>antibioticreviewkit.org.uk</u>. Acute hospital providers should embed the ARK-hospital toolkit in their staff-training, prescribing processes and stewardship work to reduce antibiotic overuse and protect their patients from antibiotic-related harms.

## Data Availability

All data produced in the present study are available upon reasonable request to the authors

## Acknowledgements

Programme Steering Committee: Alison Holmes (Chair), Philip Gothard, Karla Hemming, Cliff Gorton. Data Monitoring Committee: Neil French (Chair), Tom Rogers, James Lewis.

## The ARK Hospital Team

**SOUTH TEES HOSPITALS NHS FOUNDATION TRUST**: Sath Nag, Richard Bellamy, Debbie Lockwood

**ROYAL CORNWALL HOSPITALS NHS TRUST:** Neil Powell

**LONDON NORTH WEST HEALTHCARE NHS TRUST:** Cassandra Watson, Priya Khanna, Elizabeth Fleet, Kam Cheema, Amit Amin, Alastair McGregor, G Gopal Rao, Mushtaqur Rahman, Sunder Chita

**WYE VALLEY NHS TRUST:** Alison Johnson, Sue Vaughan

**MID YORKSHIRE HOSPITALS NHS TRUST:** Stuart Bond, Jade Lee-Milner

**ROYAL UNITED HOSPITALS BATH NHS FOUNDATION TRUST:** Mandy Slatter, Jane Liu, Belen Espina

**BETSI CADWALADR UNIVERSITY HEALTH BOARD:** Charlotte Makanga

**NORTH MIDDLESEX UNIVERSITY HOSPITAL NHS TRUST:** Mariyam Mirfenderesky, Semra Ali, Salih Hassan, Roxanne Forbes

**AIREDALE NHS FOUNDATION TRUST:** Kevin Frost

**COUNTESS OF CHESTER HOSPITAL NHS FOUNDATION TRUST:** Ruth McEwen, Ildiko Kustos, Joy Nicholls, Alice Rafferty

**SOUTHERN HEALTH (NORTHERN IRELAND):** Geraldine Conlon-Bingham, Sara Hedderwick, Cara McKeating, Peter McKee, Lisa Lennon, Orla McGivern, Jessie McNally, Toni Donnelly, Roger Stewart, Diarmuid McNicholl, Eoghan McCloskey, Wendy Baird, Donna Muckian, Martin Brown

**NEWCASTLE UPON TYNE HOSPITALS NHS FOUNDATION TRUST:** David Price, Kathy Gillespie, Andrew Heed, Matthew Lowery

**EAST KENT HOSPITALS UNIVERSITY NHS FOUNDATION TRUST:** Stephen Glass, Amy Dalton, Doreen Flower, Christopher Parokkaren

**MILTON KEYNES HOSPITAL NHS FOUNDATION TRUST:** John Northfield, Prithwiraj Chakrabarti

**FRIMLEY HEALTH NHS FOUNDATION TRUST:** Veronica Garcia-Arias, Consuelo Amigo-Vaquero, Patrick Doyle, Paul Flattery, Sarah Short

**GREAT WESTERN HOSPITALS NHS FOUNDATION TRUST:** Gloria Kiapi, Laura Gonzalez, Sally Tipping, Mark Hackett

**CHESTERFIELD ROYAL HOSPITAL NHS FOUNDATION TRUST:** Amanda Pegden, Umi Quinn, Caroline Duffin, Sawsan Awad

**POOLE HOSPITAL NHS FOUNDATION TRUST:** Elizabeth Sheridan, Samantha Ruston, Craig Prescott

**SOUTH EASTERN HEALTH&SOCIAL CARE (NORTHERN IRELAND):** Bernie McCullagh, Peter Yew, Frederick McIlwaine

**OXFORD UNIVERSITY HOSPITALS NHS FOUNDATION TRUST:** Nicola Jones, Louise Dunsmure, Mridula Rajwani

**BUCKINGHAMSHIRE HEALTHCARE NHS TRUST**: Jean O’Driscoll, Claire Brandish, Beeyean Ng, Jose Pereira

**ROYAL FREE LONDON NHS FOUNDATION TRUST:** Damian Mack, Stephanie Harris, Marisa Lanzman, Indran Balakrishnan, Rachel Moores, Lucy Lamb, Penny Smith, Christof Stigone

**UNIVERSITY HOSPITALS PLYMOUTH NHS TRUST:** Rosemary Fok, Suzanne Price, Robert Tilley, Austin Hunt, Neha Maasala, Anna Kampa, John Siewruk

**YORK TEACHING HOSPITAL NHS FOUNDATION TRUST:** Damian Mawer, Anita Chalmers, Melissa Cochran, Stephen Waring, Jawwad Azamk, Sachin Thakur, Alexander Brightmore, Melanie Bootland, Tina Leake, Joseph Suich, Fiori Massimo

**NOTTINGHAM UNIVERSITY HOSPITALS NHS TRUST:** Tim Hills, Sam Vickers, Liz Hart, Terence Ong, Carolyne Chee, Pete Eddowes, Adrian Jones, Chamira Rodrigo, Bethan Walsh, Sue Bowler, Annette Clarkson, Shanika Crusz, Robin Allan, Raj Shah, Cheermy Rellosa

**ROYAL DEVON AND EXETER NHS FOUNDATION TRUST:** Robert Porter, Hazel Parker, Hannah Burnett, Sally Tipping, Maximilian Gilbert, Steve Westlake

**NORTHERN DEVON HEALTHCARE NHS TRUST:** Samantha Dymond, Jack Houlton, Duncan Kaye, Rebecca Langley

**NORTH TEES AND HARTLEPOOL NHS FOUNDATION TRUST:** Rebecca Alexander, Kate Armitage, Rashmi Dube, Richard Cowan, Helen Dunn, Ahmed Mohamed, Greg Morris, Naveen Aggarwal, Rachel Hodson, Steve Yeomans, Ben Prudon, Richard Thomas

**EAST SUFFOLK AND NORTH ESSEX HOSPITAL NHS FOUNDATION TRUST:** Immo Weichert, Gillian Urwin, Holly Gissing

**WIRRAL UNIVERSITY TEACHING HOSPITAL NHS FOUNDATION TRUST:** David Harvey, Amy Wilson

**SWANSEA BAY HEALTH BOARD:** Phil Coles, Julie Harris, Charlotte Richards

**THE ROYAL WOLVERHAMPTON NHS TRUST:** Parmjit Kang, Mike cooper, Kate Willmer, Jaspreet Kaur, Phillip Thomas, Atul Gulati, Tariq Aljemmali, Sian Rowley, Jayne Lawrence, Lucy Stelfox, Daniel Rosamond, Alison Walker, Rebecca Brady

**NORTH CUMBRIA INTEGRATED CARE NHS FOUNDATION TRUST**: Clare Hamson, Jan Forlow

**ROYAL BERKSHIRE NHS FOUNDATION TRUST:** Shabnam Iyer, Andy Walden, Wajid Rafai, Varun Nelatur

**SURREY AND SUSSEX HEALTHCARE NHS TRUST:** Bruce Stewart, Amy Lee, Martin Dachsel, Daniel Woosey

**WESTERN HEALTH (NORTHERN IRELAND):** Cairine Gormley, Gerard Glynn, Christopher Armstrong, Emer Teague

**NORTHERN HEALTH (NORTHERN IRELAND):** David Farren, Eileen Dorgan, Salah Elshibly, Naomi Baldwin, Fidelma Magee, Fiona Gilmore, Oonagh McCourt, Andrew Currie, Gareth Lewis, Ronan Donnelly, Eunice Minford, James Gray, Joy Cuthbertson, Karen Darragh, Wilma Williamson, Cynthia Thompson, Laura Davidson, Alastair Thompson

**SOUTHPORT AND ORMSKIRK HOSPITAL NHS TRUST:** Katherine Gray, Rosalyn Ward

**WESTERN GENERAL HOSPITAL EDINBURGH:** Morgan Evans, Alison Cockburn, Carol Philip, Esperanza Palenzuela, Rachel McKinney

## AUTHOR CONTRIBUTIONS

TEAP, ASW, LY, MJL conceived the research. ELAC,, KS, SW, MS, AK, FM, KSH, DC, LV, SH LY, TEAP and ASW conceived and developed the intervention. MLS, RA, SB, PC, GCN, SD, ME, RF, KJF, VGA, SG, CG, KG, CH, DH, TH, SI, AJ, NJ, PK, GK, DM, CM, DM, BMcC, MM, RMcE, SN, AN, JN, JO’D, AP, RP, NP, DP, ES, MS, BS, CW, IW, MD and MJL conducted the trial. EB conducted the statistical analysis. ASW, EB and MJL wrote the first draft. All authors reviewed and approved the final manuscript.

The full protocol is available on http://www.arkstudy.ox.ac.uk/ark-for-healthcare-professionals/.

