## Supplementary Material for "Antibiotic Review Kit for Hospitals (ARK-Hospital): a stepped wedge cluster randomised controlled trial"

[Supplementary Figure S10: Antibiotic DDD/admission classified by WHO’s 2019 AWaRe categories (secondary outcome); immediate impact of implementation Access antibiotics (A), change in trend post vs pre-implementation Access antibiotics (B), immediate impact of implementation Access or Watch antibiotics (C), change in trend post vs pre-implementation Access or Watch antibiotics (D), immediate impact of implementation Watch antibiotics (E), change in trend post vs pre-implementation Watch antibiotics (F), immediate impact of implementation Reserve antibiotics (G), change in trend post vs pre-implementation Reserve antibiotics (H)	55](#_Toc95218712)

### **SUPPLEMENTARY APPENDIX**

**Supplementary Methods**

**Data**

A hospital “spell” begins at admission and ends at the time of discharge, transfer out, or death, and refers to the time spent in a single hospital. An “episode” refers to the time inpatients spend under the care of a consultant (i.e., specialist/senior doctor). Spells may have one or more episodes, and the speciality of the attending consultant is recorded for each episode. Since consultants often have both general and specialist interests, both the specialty in which they are working and contracted are recorded.

Spells data included the following routinely collected data fields: admitting hospital, admission and discharge date and time (hour/minute), demographics (age at admission, sex, ethnicity), admission method and source, discharge destination/method, patient classification (ordinary admission versus day case or regular day attender), intensive care unit (ICU) admission date, time spent in the ICU, and a measure of relative deprivation based on the postal code where the patient resides. Deprivation indices vary for England, Wales, Scotland, and Northern Ireland, so deprivation was modelled as a 0-100 percentile, with zero as the most deprived and 100 as the least deprived in that country. Postal codes and other identifiable data were not collected. Death date (within 90 days of admission) was collected in advance by sites through linkage with the national death registry. Death was considered to be completely ascertained, as the date of administrative censoring in mortality analyses was truncated backwards (**Figure S3**).

Hospital exposure was calculated from the spells data, including overnight admissions in the past year, “complex” overnight admissions in the past year (i.e., spells with >1 consultant episode, excluding episodes in A&E or rehabilitation), and emergency readmissions to hospital within 30 days of discharge. Since pseudonymised patient identifiers were unique within (not across) hospital organisations, hospital exposure estimates were limited to the same hospital organisation.

The episodes data contained primary and secondary diagnosis codes, the main specialty and treatment specialty of the attending consultants, procedure codes and their dates. Admission specialty was categorised by frequency of consultant specialties used to define the study population (**Figure S2**), beginning with the most common overall (acute/general medicine) and followed the next most common specialty, with the least common specialties (cumulatively accounting for 10% of spells) being collapsed into an “other” category. Primary diagnosis (ICD-10) codes were used to derive 142 Clinical Classifications Software (CCS) diagnosis groups based on guidance by NHS Digital.^1^ As previously,^2^ these categories were initially collapsed into 42 subgroups based on clinical feedback. Categories with low 30-day mortality (<1%) were collapsed into a “low risk” group, and the smallest categories (cumulatively accounting for 5% of spells) were combined into two groups—one with 30-day mortality above the overall risk of death in all sites (4.4%), and another where mortality was below this level. Patients with a primary diagnosis code indicating COVID-19 were combined into the CCS category containing influenza and other upper respiratory viruses. Secondary diagnosis codes were used to derive Charlson Comorbidity Index and immunosuppression status (based on the presence of metastatic cancer, severe liver disease, or HIV). All categories are displayed in **Table S5**.

To reduce outlier influence, overnight admissions, Charlson Comorbidity index, and age were truncated at their 99^th^ percentiles. Non-linear covariates were modelled using restricted natural cubic splines, selecting up to 5 knots to minimise the Bayesian information criterion, with 5 Harrell knots for age, 4 knots at the 20^th^, 40^th^, 60^th^, and 80^th^ percentiles from the non-zero distribution for Charlson comorbidity score, and 4 knots at the 10th, 75th, 90th, and 95^th^ percentiles from the non-zero distribution for overnight admissions in the past year (placing knots at evenly spaced intervals was not possible due to limited variation in the truncated distribution). There was no evidence of non-linearity in the effect of deprivation percentile, and modelling overnight complex admissions in the past year as binary (ever versus never) gave the best fit.

The bulk antibiotic data included antibiotic defined daily doses (DDD) by month, antimicrobial agent, and route of administration, on the general medical wards which implemented the ARK intervention. The *C. difficile* data included the faecal sample collection date (and time if available), test type (e.g., glutamase dehydrogenate, toxin enzyme immunoassay, toxin polymerase chain reaction, cell cytotoxicity assay), and the test result (i.e., positive, negative, test failed).

Although sites were asked to optionally provide patient-level antibiotic data (in hospital and at discharge), blood microbiological culture and drug susceptibility results, haematology and biochemistry test results, and imaging test results, very few were able to and so these outcomes were not analysed further.

All patients in the National Health Service can opt out from having their health records used for research or planning purposes, and this remains in effect after death. Sites were asked to exclude patients who were known to have opted out, but were not asked to provide numbers opting out. Rates are expected to be low, for example, being 2.6% in England at October 2020.^3^

Data sharing agreements were signed with each participating site, outlining the required data fields and deadlines for data submissions at the trial end in October 2020. The collection of this data required collaboration from hospital informatics teams, antimicrobial pharmacists, microbiology and IT departments, and sites were financially compensated for the data. Due to the ongoing COVID-19 pandemic, departments varied widely in their capacity to provide the requested data in a timely manner, and some sites needed to provide multiple copies of each dataset to resolve queries. Delays in data submissions meant data from all 39 sites was not obtained until October 2021.

We adhered to the Consolidated Standards of Reporting Trials (CONSORT) checklist for stepped wedge cluster randomised trials (**Table S7**).

*Data cleaning steps (co-primary outcomes)*

A total of 7,657,699 spells meeting the study inclusion criteria were submitted by the 39 pilot and main trial sites, of which 7,160,421 (93.5%) were included in the analysis of 30-day mortality (co-primary outcome) after dropping spells with missing covariates (**Figure S3**), incorrectly calculated previous admissions and other data errors (**Table S6B**) and all spells beginning less than 30 days before the last known date of death in each site. Patient characteristics are outlined in **Table S5**. Missing data by site is presented in **Table S8.**

Only systemic antibiotics were analysed, and anti-tuberculosis medications were excluded. Time trends for individual antibiotics were reviewed for each site. One pilot site was excluded from all analyses of antibiotic use, as they introduced an electronic prescribing system (JAC) 6 months after implementation and this led to an immediate decline (>90%) in reported antibiotic DDDs which prevented valid assessment of post-implementation trends in antibiotic use. Of the remaining 38 sites analysed, 36 provided DDDs for antibiotics prescribed on the general medical wards where the intervention was implemented; however, two sites could only provide hospital-level DDDs due to limitations posed by local pharmacy information systems (sites 22 and 30). Admissions were estimated from the electronic health records submitted by sites. Both the numerator (antibiotic DDDs consumed on general medical wards) and denominator (acute/general medical admissions) were collected to reflect – as accurately as was possible using available routine data reporting systems – the patients cared for by healthcare professionals targeted by the ARK intervention. Time periods for the antibiotic analysis and further cleaning steps are summarised in **Table S6A**.

*Data cleaning steps – secondary outcomes*

*Mortality within 90 days of admission (in/out of hospital)*

Cleaning steps included all those used for 30-day mortality (**Figure S3** and **Table S6B**). However, rather than dropping all spells beginning less than 30 days before the last known date of death in each site, spells beginning within 90 days were dropped instead (n=169,967/7,331,890). Incomplete ascertainment of death was apparent among spells in the final month of three sites (very low death rates) so these months were dropped (n=16,328 spells). The final month in seven sites was dropped as the sample size was low (<500 spells in each month), resulting in imprecise mortality estimates (n=1,914 admissions dropped). Near the start of the study period, six sites clearly undercounted previous admissions, and the earliest months with substantially lower estimates were dropped (i.e., prior to rates stabilising, n=127,430 admissions dropped). In one site, diagnostic coding was incomplete in the final month so the month was dropped (n=1,557 admissions dropped). The main analysis included 7,014,694 spells from 39 sites. A sensitivity analysis dropped admissions from March 2020 onwards (n=596,855), and 12 sites with <12 months post-implementation data as a result (n=2,046,503 spells), leaving 4,371,336 spells from 27 sites in the sensitivity analysis.

*Secondary antibiotic outcomes*

Cleaning steps included all those used for total DDDs/admission (co-primary outcome). However, when analysing carbapenem DDDs/admission, one hospital organisation (site 5) was excluded as it reported no carbapenem use in 55/56 months, leaving 37 sites in the final analysis. The analysis of total DDDs/bed-day estimated bed-days from the electronic health records submitted by sites, calculated as overnight stays +1 (i.e., so if a bed was occupied for part of one day then bed-day was counted as 1, and patients with admissions spanning >1 calendar month contributed bed-days to each month). To avoid undercounting bed-days, the earliest month was dropped in 34/38 sites.

When route of administration was not available, this was inferred from drug formulation by study consultant microbiologists and infectious diseases specialists. Cumulatively, antibiotic formulations with an ambiguous route of administration accounted for a median of 0.1% (IQR: 0.0-4.9%, range: 0-9.4%) of antibiotic DDDs across the sites, and thus misclassification errors are likely to be relatively low and independent of the exposure (non-differential) when parenteral and oral DDDs/admission are modelled as a secondary outcome.

*Emergency readmission*

One study site did not estimate readmissions correctly (based on other submitted data) and was dropped from this analysis (n=272,169/7,331,890 spells). The analysis was restricted to patients discharged alive within 60 days of admission in whom mortality post-discharge could be ascertained (dropped 250,412 spells). To ensure all patients had the potential to be readmitted, spells beginning within 30 days of the last readmission in each site were dropped (n=68,666); this left the final month of 14 sites with very few admissions which were dropped (n=1,430). As described in the analysis of 30-day mortality (**Figure S3**), 2 sites had incomplete diagnosis codes in the final month which were therefore dropped (n=2,674), while 5 of the remaining sites undercounted previous admissions near the start of the dataset and these months were dropped (n=110,997). The analysis included 6,625,542 spells from 38 sites.

*Intensive care unit admission*

One site was excluded as it did not provide information about ICU admissions (n=283,346/7,331,890 spells), while a second site was excluded as ICU information was not provided after October 2019 and this left too little data to estimate post-implementation trends (n=106,160). Of the remaining spells, 99% lasted 45 days or less, so the dataset was truncated 45 days from the final admission in each site (dropped 152,205 spells). In one site, no admissions to ICU were recorded in the final 2 months of the dataset and these months were dropped (n=6,444), and 4 sites had very few remaining spells in the final month so these spells were dropped (n=1,670). As in the analysis of 30-day mortality (**Figure S3**), six of the remaining sites substantially undercounted previous admissions near the start of the study period and these months were dropped (n=127,430), while one site was missing diagnosis codes near the close of the dataset the final month was dropped (n=680). In one site, one of the admission specialty categories had no ICU admissions recorded and was therefore combined with the mode category (i.e., general medicine, n=207 spells). One site showed a sudden (subsequently sustained) doubling of ICU admissions during the pre-implementation phase; excluding data before this step change was not possible as the site was left with too little data to estimate pre-implementation trends, so the site was excluded (n=158,697). The final analysis included 6,499,923 spells from 36 sites.

*Length of hospital stay (time from admission to discharge alive)*

One site was dropped (n=272,169/7,331,890 spells) as the time (hour/minute) of admission and discharge was often identical and this problem increased over time from a low of 1-2% of spells in 2016 to over 20% in 2020, preventing accurate estimates. To ensure each patient had the potential for a long spell, admissions beginning within 47 days (the 99^th^ percentile of length of stay among remaining sites) of the last recorded discharge date were dropped (n=82,258). This led to small numbers of admissions in the final month of six sites so these months were dropped (n=2,182 spells). Admissions were calculated incorrectly near the start of the dataset in 5 of the remaining sites; as in the analysis of 30-day mortality (**Figure S3**), the earliest months with the most severe undercounting were dropped (n=115,229) and at one site diagnosis codes were not complete after June 2020 so these spells were dropped (n=908). In three sites, the frequency of identical admission and discharge times changed over time so the earliest months were dropped to minimise the impact on estimated pre-implementation trends (n=64,460). The final analytic cohort included 6,794,684 spells from 38 sites.

An analysis of length of stay from admission to medically fit for discharge was not performed although in the Statistical Analysis Plan because the date patients were medically fit for discharge was not reported in a median of 97.9% of spells (IQR 59.5-100%) per site.

*C. difficile infection and colonisation within -1/+90 days of admission*

The testing algorithm used in each site was recorded, including whether this changed from year-to-year or by day of the week (e.g., out of hours or on weekends). Submitted test results were compared against the known testing algorithms to flag and resolve queries with sites (e.g., identify missing tests or ambiguous results). One site did not provide toxin tests and was therefore dropped, as it was not possible to reliably identify patients with *C. difficile* infection versus colonisation (n=186,661/7,331,890 spells). The month of the last specimen collection was identified in each site, and spells beginning in this month and the previous 2 months were dropped to enable ascertainment of infection post admission (n=233,881). In six sites, previous admissions were substantially undercounted at the start of the study period; these months were dropped, as in the analysis of 30-day mortality (**Figure S3**) (n=127,430). At one site, diagnosis codes were incomplete after June 2020 and these spells were dropped (n=527). The month of the first specimen collection was identified, and any spell starting before this month was dropped (n=54,954 spells from four sites). The analysis of *C. difficile* infection included 6,728,437 spells from 38 sites.

Colonisation (toxigenic or non-toxigenic) was analysed exclusive of infection. In addition to the cleaning steps carried out to analyse *C. difficile* infection, 4 sites were excluded as negative test results could not be reliably distinguished from results indicating colonisation (n=618,731). One site changed testing algorithms in October 2019 and their results showed no instances of colonization after this date; the proportion of patients tested each month was similar before and after the algorithm change time, so the absence of results indicating colonisation was judged to be a result of changing reporting formats and data from October 2019 onwards was dropped in this site (n=17,416). The analysis of *C. difficile* colonisation included 6,092,290 spells from 34 sites.

*Specific models*

For ICU admission and *C. difficile* infection/colonisation, due to low event rates age was included as linear term rather than a restricted natural cubic spline and patient classification was excluded. Models of emergency readmission to hospital also adjusted for the length of the admission from which discharge was counted, and included a second COVID adjustment to account for brief but pronounced declines in readmission rates in March 2020. *C. difficile* infection/colonisation models excluded admission day of year and the COVID adjustment as there was no evident seasonality or step change in 2020. Outcome trends pre- and post-implementation are summarised in **Table S9**.

*Sample size calculation*

For the main trial, based on simulation studies, the proposed stepped wedge cluster randomised design including data from a minimum of 36 organisations has >85% power to exclude a 5% relative increase in 30-day mortality and to detect a 15% relative reduction in antibiotic use associated with the intervention. The 5% relative non-inferiority margin means that to declare non-inferiority, the entire 95% confidence interval (CI) for the relative change in mortality post vs pre intervention has to lie within [0.95,1.05]. For an overall mortality of 9%, this is analogous to being confident that the post-intervention mortality lies within [8.5%, 9.5%]; that is a non-inferiority margin of +/- 0.5% on the absolute scale or 5% on the relative risk or relative odds scale (which are similar given the low event rates).

This sample size calculation was based on simulation, given the uncertainties regarding parameters underlying the study design. Simulations were run 1000 times for N=10 to 40 participating organisations in increments of 2 organisations and a relative decrease in antibiotic use of R=5% to 20% in increments of 5%. Organisations were assumed to vary in size with an underlying rate of acute/general medicine admissions per year from 5000 to 15000 (uniform distribution), plus one large organisation of 25000 admissions per year (estimates based on acute/general medical admissions in Oxford University Hospitals (OUH) NHS Foundation Trust, and publicly available data on the distribution of annual overnight stays in all English NHS Trusts). Before the intervention is introduced, each organisation is assumed to have an underlying rate of 1000 DDD per acute/general medical admission drawn from a uniform distribution on 1500-3000. Each organisation’s 30-day mortality is assumed to be distributed uniformly between 7-10%, independently of antibiotic prescribing. The intervention was assumed to decrease DDD per 1000 admissions by R% with no effect on mortality: there are no time trends before or after implementation in either outcome. The intervention is implemented in 2 randomly chosen organisations every month for 18 months, followed by a further 12 months post-intervention follow-up. Each month, the number of acute/general medical admissions are simulated using a Poisson distribution based on the underlying rate simulated as above. Each month, DDD are then simulated from a negative binomial distribution based on the simulated number of admissions that month, each organisation’s simulated underlying prescribing rate and whether the month falls before or after the randomly allocated intervention time. The dispersion parameter is fixed at 1.15 times the mean, slightly higher than the estimated dispersion from monthly antibiotic prescribing in OUH acute/general medical inpatients [33]. Each month, the 30-day mortality is then simulated from a binomial distribution based on the simulated number of admissions that month, with a probability drawn from a normal distribution centred on each organisation’s simulated underlying mortality rate with standard deviation 0.1. Analysis follows methods above. No sample size inflation was applied to the two co-primary outcomes because the goal of the non-inferiority comparison is to demonstrate that the 95% CI around the estimated relative risk is within a pre-defined non-inferiority margin, rather than identify a significant difference.

### **Supplementary Tables**

#### Supplementary Table S1: Pre-specified implementation fidelity criteria

| **Criteria** | **Achieved** |
| --- | --- |
| C1: Provision of a list of key essential people – by implementation date | 31/39 (79%) |
| C2: Achieving at least 20 people per 100 acute beds having done the online learning - by end of implementation | 25/39 (64%) |
| C3: Introduction of the ARK categories into the prescribing process - by implementation date | 31/39 (79%) |
| C4: Process in place for making patient leaflet available to acute medical patients - by implementation date | 25/39 (64%) |
| C5: Submission of baseline audit data – by implementation date | 19/39 (49%) |
| C6: Process in place for ongoing audit and feedback – by implementation date | 37/39 (95%) |
| C7: Submission of post implementation audit data – by week 4 | 30/39 (77%) |
| C8: Submission of electronic patient research data – by week 16 following implementation^1^ | 21/36 (58%) |
| Total criteria achieved (median, IQR, range) | 6 (5-7) [2-8] |

^1^ Not required for pilot sites: treated as achieved in analysis.

#### Supplementary Table S2: Antibiotics classified by spectrum of activity and by the World Health Organisation’s 2019 Essential Medicines List AWaRe categories

| **Antibiotic** | **ATC index code 2020** | **Classified as broad-spectrum**^1^ | **AWaRe category (England)** |
| --- | --- | --- | --- |
| Amikacin | J01GB06 | No | Watch |
| Amoxicillin | J01CA04 | No | Access |
| Amoxicillin/Clavulanic Acid | J01CR02 | Yes | Watch |
| Ampicillin | J01CA01 | No | Access |
| Azithromycin | J01FA10 | No | Access or Watch depending on indication |
| Aztreonam | J01DF01 | Yes | Reserve |
| Benzathine Benzylpenicillin | J01CE08 | No | Access |
| Benzylpenicillin | J01CE01 | No | Access |
| Cefaclor ^2^ | J01DC04 | No | Watch |
| Cefadroxil | J01DB05 | No | Watch |
| Cefalexin | J01DB01 | No | Watch |
| Cefazolin | J01DB04 | No | Watch |
| Cefepime | J01DE01 | Yes | Reserve |
| Cefixime | J01DD08 | Yes | Access or Watch depending on indication |
| Cefotaxime | J01DD01 | Yes | Access or Watch depending on indication |
| Cefoxitin | J01DC01 | Yes | Watch |
| Cefradine | J01DB09 | No | Watch |
| Ceftaroline Fosamil | J01DI02 | Yes | Reserve |
| Ceftazidime | J01DD02 | Yes | Watch |
| Ceftazidime/Avibactam | J01DD52 | Yes | Reserve |
| Ceftobiprole Medocaril | J01DI01 | Yes | Reserve |
| Ceftolozane/Tazobactam | J01DI54 | Yes | Reserve |
| Ceftriaxone | J01DD04 | Yes | Access or Watch depending on indication |
| Cefuroxime | J01DC02 | Yes | Watch |
| Chloramphenicol | J01BA01 | No | Watch |
| Cilastatin/Imipenem | J01DH51 | Yes | Reserve |
| Ciprofloxacin | J01MA02 | Yes | Access or Watch depending on indication |
| Clarithromycin | J01FA09 | No | Access or Watch depending on indication |
| Clindamycin | J01FF01 | No | Watch |
| Colistin | J01XB01 | Yes | Reserve |
| Dalbavancin | J01XA04 | No | Reserve |
| Daptomycin | J01XX09 | No | Reserve |
| Doxycycline | J01AA02 | No | Access |
| Ertapenem | J01DH03 | Yes | Reserve |
| Erythromycin | J01FA01 | No | Watch |
| Flucloxacillin | J01CF05 | No | Access |
| Fosfomycin (not administered parenterally) | J01XX01 | No | Access |
| Fosfomycin (parenterally) | J01XX01 | No | Reserve |
| Fusidic Acid ^3^ | J01XC01 | No | Access |
| Gentamicin | J01GB03 | No | Access |
| Levofloxacin | J01MA12 | Yes | Watch |
| Linezolid | J01XX08 | No | Reserve |
| Meropenem | J01DH02 | Yes | Reserve |
| Metronidazole ^4^ | J01XD01 | No | Access |
| Minocycline | J01AA08 | No | Watch |
| Moxifloxacin | J01MA14 | Yes | Watch |
| Nitrofurantoin | J01XE01 | No | Access |
| Ofloxacin | J01MA01 | Yes | Watch |
| Oxytetracycline | J01AA06 | No | Watch |
| Phenoxymethylpenicillin | J01CE02 | No | Access |
| Piperacillin/Tazobactam | J01CR05 | Yes | Access or Watch depending on indication |
| Pivmecillinam | J01CA08 | No | Access |
| Pristinamycin | J01FG01 | No | Watch |
| Procaine Benzylpenicillin | J01CE09 | No | Access |
| Rifampicin | J04AB02 | No | Watch |
| Spectinomycin | J01XX04 | No | Access |
| Sulfadiazine | J01EC02 | No | Access |
| Sulfamethoxazole/Trimethoprim | J01EE01 | No | Access |
| Tedizolid | J01XX11 | No | Reserve |
| Teicoplanin | J01XA02 | No | Watch |
| Temocillin | J01CA17 | No | Watch |
| Tetracycline | J01AA07 | No | Access |
| Tigecycline | J01AA12 | Yes | Reserve |
| Tinidazole | J01XD02 | No | Access |
| Tobramycin | J01GB01 | No | Watch |
| Trimethoprim | J01EA01 | No | Access |
| Vancomycin | J01XA01 | No | Access or Watch depending on indication |

^1^ Broad-spectrum agents were defined as: carbapenems, quinolones, polymyxins, co-amoxiclav, piperacillin/tazobactam, azithromycin, tigecycline, aztreonam, telithromycin, cephalosporin-beta-lactamase inhibitor combinations (e.g., ceftazidime-avibactam, ceftolozane-tazobactam), and second (e.g., cefuroxime), third (e.g., ceftriaxone, ceftazidime), fourth (e.g., cefepime) or fifth (e.g., ceftobiprole) generation cephalosporins.

^2^ All second-generation cephalosporins were categorised as broad-spectrum agents with the exception of Cefaclor as it is administered orally and not well absorbed.

^3^ Excluded when administered topically.

^4^ ATC codes were missing in data from some sites, as a result this category may also include metronidazole (P01AB01).

#### Supplementary Table S3: Summary of impact of the ARK intervention

| Outcome | Model used to derive site-specific estimates | Analysis  type | Random effects meta-analysis | |
| --- | --- | --- | --- | --- |
|  |  |  | Immediate implementation effect: OR, IRR, or SHR (95% CI), p-value^1^ | Change in trend post- vs pre-implementation: OR, IRR, or SHR (95% CI), p-value^1^ |
| Total DDD/admission^2^ | Adjusted negative binomial model, with COVID adjustment, 38 sites (**Figure 4**) | 1 | 0.99 (0.96, 1.02), p=0.540 | 0.95 (0.91, 1.00), p=0.042 |
| Total DDD/admission | Adjusted negative binomial model, without COVID adjustment, 38 sites | 2 | 0.98 (0.95, 1.01), p=0.159 | 1.00 (0.95, 1.04), p=0.864 |
| Total DDD/admission | Adjusted negative binomial model, excluding follow-up after March 2020, 26 sites | 3 | 1.00 (0.96, 1.03), p=0.821 | 0.95 (0.90, 1.00), p=0.048 |
| Total DDD/admission | Unadjusted negative binomial model (i.e., no seasonal effect), with COVID adjustment, 38 sites | 4 | 1.00 (0.96, 1.03), p=0.840 | 0.94 (0.89, 0.98), p=0.008 |
| Total DDD/admission | Adjusted negative binomial model, with COVID adjustment, main trial sites only, 36 sites | Sensi-tivity | 0.99 (0.96, 1.02), p=0.558 | 0.94 (0.90, 0.99), p=0.019 |
| Total DDD/bed-day | Adjusted negative binomial model, with COVID adjustment, 38 sites **(Figure S8)** | 1 | 1.00 (0.97, 1.03), p=0.784 | 0.96 (0.92, 1.00), p=0.056 |
| Broad-spectrum DDD/admission | Adjusted negative binomial model, with COVID adjustment, 38 sites **(Figure S9A&B**) | 1 | 0.94 (0.91, 0.97), p=0.0003 | 0.97 (0.92, 1.04), p=0.395 |
| Narrow-spectrum DDD/ admission | Adjusted negative binomial model, with COVID adjustment, 38 sites **(Figure S9C&D**) | 1 | 1.01 (0.98, 1.05), p=0.488 | 0.95 (0.91, 0.99), p=0.019 |
| Access DDD/admission | Adjusted negative binomial model, with COVID adjustment, 38 sites **(Figure S10A&B)** | 1 | 1.04 (1.00, 1.09), p=0.037 | 0.95 (0.90, 1.00), p=0.033 |
| Access or Watch DDD/ admission^3^ | Adjusted negative binomial model, with COVID adjustment, 38 sites **(Figure S10C&D)** | 1 | 0.97 (0.92, 1.02), p=0.218 | 1.01 (0.95, 1.08), p=0.762 |
| Watch DDD/admission | Adjusted negative binomial model, with COVID adjustment, 38 sites **(Figure S10E&F)** | 1 | 0.91 (0.87,0.94), p<0.0001 | 0.89 (0.83,0.96), p=0.001 |
| Reserve-spectrum DDD/ admission | Adjusted negative binomial model, with COVID adjustment, 38 sites **(Figure S10G&H)** | 1 | 1.03 (0.94, 1.13), p=0.543 | 1.07 (0.98, 1.17), p=0.107 |
| Parenteral DDD/ admission | Adjusted negative binomial model, with COVID adjustment, 38 sites (**Figure S11A&B**) | 1 | 0.99 (0.96, 1.03), p=0.655 | 0.99 (0.95, 1.04), p=0.703 |
| Oral DDD/ admission | Adjusted negative binomial model, with COVID adjustment, 38 sites **(Figure S11C&D)** | 1 | 0.99 (0.95, 1.02), p=0.458 | 0.94 (0.89, 0.99), p=0.012 |
| Carbapenem DDD/ admission | Adjusted negative binomial model, with COVID adjustment, 37 sites **(Figure S12)** | 1 | 1.04 (0.95, 1.14), p=0.376 | 1.12 (1.02, 1.23), p=0.015 |
| 30-day mortality^2^ | Adjusted logit model, with COVID adjustment, 39 sites (**Figure 5**) | 1 | 0.97 (0.94, 1.00), p=0.079 | 1.03 (1.00, 1.06), p=0.060 |
| 30-day mortality | Adjusted logit model, without COVID adjustment, 39 sites | 2 | 0.93 (0.90, 0.96), p<0.0001 | 1.18 (1.14, 1.22), p<0.0001 |
| 30-day mortality | Adjusted logit model, excluding follow-up from March 2020, 27 sites | 3 | 0.98 (0.95, 1.02), p=0.357 | 0.99 (0.96, 1.02), p=0.602 |
| 30-day mortality | Unadjusted logit model, with COVID adjustment and seasonal effect, 39 sites | 4 | 0.94 (0.91, 0.97), p<0.0001 | 1.07 (1.03, 1.11), p=0.0003 |
| 30-day mortality | Adjusted logit model, with COVID adjustment, excluding admissions to cardiology, rheumatology, haematology and neurology, 39 sites^4^ | Sensi-tivity | 0.97 (0.94,1.00), p=0.054 | 1.03 (0.99, 1.06), p=0.136 |
| 30-day mortality | Adjusted logit model, with COVID adjustment, main trial sites only, 36 sites | Sensi-tivity | 0.96 (0.93, 0.99), p=0.021 | 1.03 (1.00, 1.07), p=0.066 |
| 90-day mortality | Adjusted logit model, with COVID adjustment, 39 sites (**Figure S15**) | 1 | 0.97 (0.93, 1.01), p=0.092 | 1.04 (1.01, 1.07), p=0.023 |
| 90-day mortality | Adjusted logit model, excluding follow-up from March 2020, 27 sites | 3 | 0.96 (0.92, 1.01), p=0.098 | 1.03 (0.99, 1.08), p=0.182 |
| ICU admission | Adjusted logit model, with COVID adjustment, 36 sites **(Figure S16)** | 1 | 1.02 (0.98, 1.07), p=0.284 | 0.94 (0.87, 1.02), p=0.123 |
| Length of hospital stay (days) | Adjusted competing risks model, with COVID adjustment, 38 sites **(Figure S17)** | 1 | 1.00 (0.99, 1.00), p=0.480 | 1.00 (0.99, 1.01), p=0.766 |
| 30-day emergency readmission to hospital | Adjusted logit model, with COVID adjustment, 38 sites **(Figure S18)** | 1 | 1.00 (0.97, 1.03), p=0.957 | 0.98 (0.95, 1.02), p=0.330 |
| 30-day emergency readmission to hospital | Adjusted competing risks model, with COVID adjustment, 38 sites | Sensi-tivity | 1.00 (0.98, 1.02), p=0.959 | 0.99 (0.96, 1.01), p=0.325 |
| *C difficile* infection within 90 days of admission | Adjusted logit model, without COVID adjustment 38 sites **(Figure S19A&B)** | 1 | 0.95 (0.83, 1.09), p=0.486 | 1.06 (0.92, 1.22), p=0.432 |
| *C difficile* colonisation within 90 days of admission | Adjusted logit model, without COVID adjustment, 34 sites **(Figure S19C&D)** | 1 | 0.94 (0.83, 1.07), p=0.374 | 0.93 (0.83, 1.05), p=0.243 |

^1^ OR = Odds Ratio (logit models), IRR = Incident Rate Ratio (negative binomial models), SHR = Subdistribution Hazard Ratio (competing risks models)

^2^ See **Figure S5** for fit to individual sites, with and without a COVID adjustment. All adjusted models include a seasonal effect, with the exception of *C difficile* outcomes.

^3^ Antibiotics in this category may be considered either Access or Watch depending on indication. Since indication was unknown, they were analysed separately (**Table S2**).

^4^ As outlined in **Figure S2**, the specialty codes used to identify acute/general medicine inpatients were selected to be highly sensitive, but specificity may vary by site. To improve specificity, this sensitivity analysis excluded admissions to cardiology, rheumatology, haematology, and neurology.

#### Supplementary Table S4 Random-effects meta-regression comparing the association between potential effect modifiers and site-level effect estimates derived from models of total DDD/admission and 30-day mortality (co-primary outcomes)

| **Effect modifiers considered**  **(all models are univariate unless noted otherwise)** | **Category** | **Outcome in meta-regression model (i.e. site-level effect estimates)** | | | |
| --- | --- | --- | --- | --- | --- |
|  |  | **Total DDD/admission (n=38 sites)** | | **30-day mortality (n=39 sites)** | |
|  |  | **Immediate implementation effect**  **IRR (95% CI)** | **Change in trend post- vs pre-implementation IRR (95% CI)** | **Immediate implementation effect**  **OR (95% CI)** | **Change in trend post- vs pre-implementation**  **OR (95% CI)** |
| C1: Provision of a list of key essential people – by implementation date | No | Ref | Ref | Ref | Ref |
|  | Yes | 0.99 (0.91, 1.07)  p=0.735 | 0.95 (0.83, 1.10)  p=0.498 | 1.05 (0.97, 1.14) p=0.239 | 1.04 (0.95, 1.13) p=0.364 |
| C2: Achieving at least 20 people per 100 acute beds having done the online learning - by end of implementation | No | Ref | Ref | Ref | Ref |
|  | Yes | 1.01 (0.94, 1.09) p=0.691 | **0.90 (0.80, 1.01) p=0.075** | 1.04 (0.97, 1.11) p=0.278 | 1.02 (0.95, 1.09) p=0.552 |
| C3: Introduction of the ARK categories into the prescribing process - by implementation date | No | Ref | Ref | Ref | Ref |
|  | Yes | 0.95 (0.87, 1.04) p=0.241 | **0.89 (0.77, 1.02) p=0.083** | 1.02 (0.94, 1.11) p=0.552 | **0.93 (0.85, 1.01) p=0.075** |
| C4: Process in place for making patient leaflet available to acute medical patients - by implementation date | No | Ref | Ref | Ref | Ref |
|  | Yes | 1.02 (0.95, 1.10) p=0.532 | 1.08 (0.96, 1.21) p=0.213 | 1.00 (0.94, 1.08) p=0.905 | 1.04 (0.97, 1.12) p=0.228 |
| C5: Submission of baseline audit data – by implementation date | No | Ref | Ref | Ref | Ref |
|  | Yes | 1.00 (0.93, 1.07) p=0.919 | 1.00 (0.89, 1.12) p=0.996 | 0.99 (0.92, 1.05) p=0.654 | 1.01 (0.94, 1.08) p=0.808 |
| C6: Process in place for ongoing audit and feedback – by implementation date | No | Ref | Ref | Ref | Ref |
|  | Yes | **0.83 (0.71, 0.97) p=0.022** | 1.15 (0.89, 1.48) p=0.286 | 0.93 (0.80, 1.08) p=0.319 | 0.97 (0.84, 1.12) p=0.681 |
| C7: Submission of post implementation audit data – within 4 weeks after implementation | No | Ref | Ref | Ref | Ref |
|  | Yes | **0.92 (0.85, 0.99) p=0.027** | 0.98 (0.85, 1.12) p=0.728 | 1.01 (0.94, 1.10) p=0.732 | 0.98 (0.91, 1.06) p=0.633 |
| Multivariate model containing implementation criteria C6 and C7 (above) | C6 No | Ref | Ref | N/A | N/A |
|  | C6 Yes | 0.87 (0.74, 1.03) p=0.109 | 1.21 (0.91, 1.61) p=0.181 | N/A | N/A |
|  | C7 No | Ref | Ref | N/A | N/A |
|  | C7 Yes | 0.94 (0.87, 1.02) p=0.139 | 0.93 (0.80, 1.09) p=0.370 | N/A | N/A |
| C8: Submission of electronic patient research data – within 16 weeks after implementation (not required for pilot sites) | No | Ref | Ref | Ref | Ref |
|  | Yes | 1.06 (0.99, 1.14) p=0.105 | 0.97 (0.86, 1.10) p=0.662 | **1.07 (1.00, 1.14) p=0.047** | 0.97 (0.90, 1.04) p=0.363 |
| Total number of implementation fidelity criteria achieved (out of 8) | Per additional criteria | 0.99 (0.97, 1.02) p=0.640 | 0.99 (0.95, 1.02) p=0.448 | 1.01 (0.99, 1.03) p=0.194 | 1.00 (0.98, 1.02) p=0.926 |
| Randomisation block | Block 1 (site 4-8) | Ref | Ref | Ref | Ref |
|  | Pilot sites | 1.05 (0.87, 1.27) p=0.619 | 1.16 (0.85, 1.59) p=0.330 | 1.11 (0.97, 1.28) p=0.130 | 0.96 (0.83, 1.12) p=0.612 |
|  | Block 2 (site 9-12) | 1.05 (0.90, 1.22) p=0.560 | 0.95 (0.74, 1.22) p=0.667 | 0.97 (0.85, 1.10) p=0.645 | 0.98 (0.86, 1.12) p=0.777 |
|  | Block 3 (site 13-18) | 1.08 (0.93, 1.25) p=0.285 | 1.02 (0.81, 1.28) p=0.894 | 0.96 (0.85, 1.08) p=0.456 | 0.95 (0.84, 1.08) p=0.461 |
|  | Block 4 (site 19-21) | 1.02 (0.85, 1.21) p=0.851 | 0.99 (0.75, 1.31) p=0.932 | 1.01 (0.87, 1.17) p=0.873 | 0.99 (0.85, 1.17) p=0.931 |
|  | Block 5 (site 22-27) | 1.05 (0.91, 1.21), p=0.456 | 1.02 (0.81, 1.28) p=0.882 | 1.01 (0.89, 1.14) p=0.932 | 1.02 (0.89, 1.16) p=0.811 |
|  | Block 6 (site 28-32) | 1.07 (0.92, 1.24) p=0.397 | 1.00 (0.79, 1.28) p=0.976 | 1 (0.88, 1.14) p=0.941 | 1 (0.87, 1.16) p=0.957 |
|  | Block 7 (site ≥33) | 1.03 (0.90, 1.19) p=0.647 | 0.98 (0.78, 1.23) p=0.838 | 1.07 (0.95, 1.20) p=0.256 | 0.95 (0.84, 1.09) p=0.485 |
| Calendar period of randomised implementation date (reflecting different NHS pressures) | Jan-Mar | Ref | Ref | Ref | Ref |
|  | Apr-Jun | 1.02 (0.93, 1.11) p=0.742 | 1.00 (0.86, 1.17) p=0.967 | 1.04 (0.95, 1.13) p=0.375 | 0.97 (0.89, 1.06) p=0.487 |
|  | Jul Sep | 0.96 (0.86, 1.08) p=0.488 | 0.91 (0.76, 1.11) p=0.344 | 0.98 (0.88, 1.09) p=0.683 | **0.90 (0.81, 0.99) p=0.036** |
|  | Oct Dec | 1.03 (0.93, 1.13) p=0.620 | 1.01 (0.86, 1.20) p=0.882 | 1.04 (0.94, 1.15) p=0.441 | 0.98 (0.9, 1.08) p=0.737 |
| Site size (categorised into terciles based on beds available) | Small (≤550 beds) | Ref | Ref | Ref | Ref |
|  | Medium (551-850 beds) | **0.93 (0.85, 1.00) p=0.064** | **1.15 (1.00, 1.31) p=0.049** | 0.98 (0.90, 1.07) p=0.646 | 0.98 (0.90, 1.06) p=0.583 |
|  | Large (>850 beds) | 0.98 (0.90, 1.07) p=0.676 | 1.06 (0.92, 1.22) p=0.374 | 1.00 (0.91, 1.08) p=0.912 | 1.00 (0.92, 1.09) p=0.973 |
| UK region | South of England | Ref | Ref | Ref | Ref |
|  | North of England | 0.99 (0.90, 1.09) p=0.867 | 1.00 (0.86, 1.15) p=0.957 | 0.98 (0.90, 1.07) p=0.673 | 1.02 (0.94, 1.12) p=0.572 |
|  | Midlands / East of England | 1.00 (0.89, 1.11) p=0.939 | **0.85 (0.72, 1.01) p=0.064** | 0.96 (0.87, 1.06) p=0.427 | 1.01 (0.91, 1.12) p=0.909 |
|  | London | 0.99 (0.86, 1.15) p=0.907 | 0.89 (0.72, 1.11) p=0.295 | 1.03 (0.90, 1.17) p=0.656 | 1.00 (0.87, 1.14) p=0.966 |
|  | Scotland | 0.92 (0.75, 1.14) p=0.438 | 1.09 (0.77, 1.53) p=0.628 | 1.12 (0.83, 1.50) p=0.445 | 0.79 (0.56, 1.11) p=0.167 |
|  | Northern Ireland | 0.95 (0.83, 1.08) p=0.397 | 1.08 (0.89, 1.32) p=0.414 | 0.92 (0.81, 1.04) p=0.183 | 1.06 (0.94, 1.20) p=0.338 |
|  | Wales | 0.87 (0.74, 1.04) p=0.116 | 1.19 (0.91, 1.55) p=0.188 | 1.06 (0.91, 1.23) p=0.432 | 0.94 (0.80, 1.11) p=0.466 |
| Functional role of the local champion (study lead) | Microbiology / Infectious diseases | Ref | Ref | Ref | Ref |
|  | Acute medicine | 0.97 (0.89, 1.07) p=0.523 | 0.96 (0.82, 1.13) p=0.631 | 1.02 (0.92, 1.12) p=0.734 | 0.96 (0.87, 1.06) p=0.382 |
|  | Pharmacist | 0.98 (0.90, 1.06) p=0.601 | 1.01 (0.88, 1.16) p=0.865 | 0.98 (0.90, 1.06) p=0.574 | 1.03 (0.96, 1.12) p=0.394 |
| Paper versus electronic prescribing systems | Paper | Ref | Ref | Ref | Ref |
|  | Electronic prescribing system | 0.99 (0.92, 1.06) p=0.697 | 0.97 (0.86, 1.09) p=0.585 | 1.00 (0.93, 1.07) p=0.996 | 0.97 (0.91, 1.04) p=0.399 |
| Implemented a hard stop versus soft stop/neither | Soft stop/neither | Ref | Ref | Ref | Ref |
|  | Hard stop | 0.96 (0.89, 1.02) p=0.186 | 1.03 (0.92, 1.16) p=0.594 | 1.01 (0.94, 1.08) p=0.844 | **0.93 (0.87, 0.99) p=0.020** |
| Percentage of essential people completing training within 12 weeks of implementation | Per 10% increase | 1.00 (0.99, 1.02) p=0.836 | 0.99 (0.97, 1.01) p=0.462 | 1.01 (0.99, 1.02) p=0.306 | 1.01 (0.99, 1.02) p=0.392 |
| Total number of healthcare workers completing training by 12 weeks per 100 acute beds | Per increase of 10 people | 1.00 (0.98, 1.01) p=0.921 | 1.00 (0.97, 1.02) p=0.812 | 1.01 (1.00, 1.03) p=0.123 | 1.01 (0.99, 1.02) p=0.360 |
| Years between implementation and March 2020 | Per 1 year later | 0.99 (0.92, 1.06) p=0.763 | 1.05 (0.93, 1.18) p=0.449 | 1.00 (0.94, 1.07) p=0.968 | 0.99 (0.93, 1.06), p=0.868 |
| Years between start of follow-up and implementation | Per 1 year later | 0.99 (0.93, 1.06) p=0.874 | 0.96 (0.86, 1.07) p=0.425 | 1.01 (0.96, 1.07) p=0.724 | 0.98 (0.92, 1.03) p=0.405 |

Note: heterogeneity p<0.1 shown in bold underline.

#### Supplementary Table S5: Characteristics of 7,160,421 Acute/General Medicine Admissions in 39 sites in 30-day mortality analysis (co-primary endpoint)

| **Characteristic** |  |  | **Died within 0-30 days of admission** | |
| --- | --- | --- | --- | --- |
|  |  | **Admissions**  **N=7,160,421 (col %)** | **No**  **N=6,846,108 (col %)** | **Yes**  **N=314,313 (col %)** |
| **Sex** | Female | 3,615,319 (50.5%) | 3,466,835 (50.6%) | 148,484 (47.2%) |
|  | Male | 3,545,102 (49.5%) | 3,379,273 (49.4%) | 165,829 (52.8%) |
| **Age at admission^1^** | Median (IQR) | 67 (53-79) | 67 (52-78) | 81 (71-88) |
| **Ethnic category** | White | 5,368,034 (75.0%) | 5,124,816 (74.9%) | 243,218 (77.4%) |
|  | Asian | 294,376 (4.1%) | 286,556 (4.2%) | 7,820 (2.5%) |
|  | Black | 126,127 (1.8%) | 123,216 (1.8%) | 2,911 (0.9%) |
|  | Other | 115,175 (1.6%) | 112,146 (1.6%) | 3,029 (1.0%) |
|  | Unknown | 1,256,709 (17.6%) | 1,199,374 (17.5%) | 57,335 (18.2%) |
| **Immunosuppressed^2^** | No | 6,906,379 (96.5%) | 6,650,523 (97.1%) | 255,856 (81.4%) |
|  | Yes | 254,042 (3.5%) | 195,585 (2.9%) | 58,457 (18.6%) |
| **Charlson co-morbidity index^1^** | Median (IQR) | 0 (0-9) | 0 (0-8) | 14 (4-21) |
| **Deprivation percentile 0-100 (0 is most deprived)^3^** | Median (IQR) | 53.9 (29.3-77.3) | 53.9 (29.3-77.3) | 54.0 (29.6-77.0) |
| **Admission method** | A&E | 2,818,261 (39.4%) | 2,583,878 (37.7%) | 234,383 (74.6%) |
|  | Elective & Other non-emergency | 3,417,545 (47.7%) | 3,385,663 (49.5%) | 31,882 (10.1%) |
|  | Emergency via GP or other source | 924,615 (12.9%) | 876,567 (12.8%) | 48,048 (15.3%) |
| **Admission source** | Usual/other place of residence | 6,923,343 (96.7%) | 6,629,998 (96.8%) | 293,345 (93.3%) |
|  | NHS general ward/other care provider | 237,078 (3.3%) | 216,110 (3.2%) | 20,968 (6.7%) |
| **Admission specialty** | General medicine | 3,596,428 (50.2%) | 3,378,153 (49.3%) | 218,275 (69.4%) |
|  | Gastroenterology | 1,107,772 (15.5%) | 1,100,145 (16.1%) | 7,627 (2.4%) |
|  | Clinical Haematology | 901,955 (12.6%) | 886,741 (13.0%) | 15,214 (4.8%) |
|  | Cardiology | 473,464 (6.6%) | 461,379 (6.7%) | 12,085 (3.8%) |
|  | Other^4^ | 1,080,802 (15.1%) | 1,019,690 (14.9%) | 61,112 (19.4%) |
| **Patient classification** | Ordinary admission | 4,039,592 (56.4%) | 3,744,368 (54.7%) | 295,224 (93.9%) |
|  | Day case admission | 2,771,252 (38.7%) | 2,755,525 (40.2%) | 15,727 (5.0%) |
|  | Regular day attender | 349,577 (4.9%) | 346,215 (5.1%) | 3,362 (1.1%) |
| **Admission date of week** | Weekday | 6,060,172 (84.6%) | 5,821,095 (85.0%) | 239,077 (76.1%) |
|  | Weekend | 1,100,249 (15.4%) | 1,025,013 (15.0%) | 75,236 (23.9%) |
| **Admission time of day** | 00:00-03:59 | 510,647 (7.1%) | 469,369 (6.9%) | 41,278 (13.1%) |
| **(hour/minute)** | 04:00-07:59 | 391,119 (5.5%) | 368,323 (5.4%) | 22,796 (7.3%) |
|  | 08:00-11:59 | 2,484,033 (34.7%) | 2,436,604 (35.6%) | 47,429 (15.1%) |
|  | 12:00-15:59 | 2,015,571 (28.1%) | 1,946,036 (28.4%) | 69,535 (22.1%) |
|  | 16:00-19:59 | 1,031,005 (14.4%) | 957,048 (14.0%) | 73,957 (23.5%) |
|  | 20:00-23:59 | 728,046 (10.2%) | 668,728 (9.8%) | 59,318 (18.9%) |
| **Admission day of year** | January to March | 1,822,398 (25.5%) | 1,735,714 (25.4%) | 86,684 (27.6%) |
|  | April to June | 1,795,397 (25.1%) | 1,716,262 (25.1%) | 79,135 (25.2%) |
|  | July to September | 1,877,032 (26.2%) | 1,804,124 (26.4%) | 72,908 (23.2%) |
|  | October to December | 1,665,594 (23.3%) | 1,590,008 (23.2%) | 75,586 (24.0%) |
| **Overnight admissions** | Median (IQR; range) | 0 (0-1) | 0 (0-1) | 1 (0-2) |
| **in past year^1^** | 0 | 4,362,166 (60.9%) | 4,236,063 (61.9%) | 126,103 (40.1%) |
|  | 1 | 1,365,517 (19.1%) | 1,287,052 (18.8%) | 78,465 (25.0%) |
|  | 2 | 619,178 (8.6%) | 572,454 (8.4%) | 46,724 (14.9%) |
|  | 3 | 317,706 (4.4%) | 291,717 (4.3%) | 25,989 (8.3%) |
|  | >3 | 495,854 (6.9%) | 458,822 (6.7%) | 37,032 (11.8%) |
| **Any complex overnight admission in past year (>1 consultant episode, excluding episodes in A&E or rehabilitation)** | No | 5,317,335 (74.3%) | 5,148,803 (75.2%) | 168,532 (53.6%) |
|  | Yes | 1,843,086 (25.7%) | 1,697,305 (24.8%) | 145,781 (46.4%) |
| **NHS acute hospital site**  **(cluster), numbered in order of randomised implementation date** | 17 | 357,432 (5.0%) | 348,112 (5.1%) | 9,320 (3.0%) |
|  | 11 | 356,399 (5.0%) | 341,582 (5.0%) | 14,817 (4.7%) |
|  | 37 | 358,080 (5.0%) | 340,167 (5.0%) | 17,913 (5.7%) |
|  | 2 | 298,967 (4.2%) | 287,501 (4.2%) | 11,466 (3.6%) |
|  | 31 | 289,994 (4.0%) | 281,948 (4.1%) | 8,046 (2.6%) |
|  | 30 | 283,346 (4.0%) | 266,072 (3.9%) | 17,274 (5.5%) |
|  | 8 | 275,615 (3.8%) | 265,970 (3.9%) | 9,645 (3.1%) |
|  | 1 | 271,737 (3.8%) | 260,697 (3.8%) | 11,040 (3.5%) |
|  | 13 | 255,827 (3.6%) | 245,873 (3.6%) | 9,954 (3.2%) |
|  | 21 | 242,365 (3.4%) | 229,951 (3.4%) | 12,414 (3.9%) |
|  | 38 | 231,388 (3.2%) | 220,491 (3.2%) | 10,897 (3.5%) |
|  | 10 | 227,647 (3.2%) | 216,728 (3.2%) | 10,919 (3.5%) |
|  | 33 | 224,165 (3.1%) | 215,500 (3.1%) | 8,665 (2.8%) |
|  | 12 | 210,299 (2.9%) | 200,108 (2.9%) | 10,191 (3.2%) |
|  | 24 | 186,661 (2.6%) | 178,400 (2.6%) | 8,261 (2.6%) |
|  | 9 | 178,006 (2.5%) | 169,303 (2.5%) | 8,703 (2.8%) |
|  | 27 | 172,362 (2.4%) | 164,094 (2.4%) | 8,268 (2.6%) |
|  | 14 | 171,804 (2.4%) | 163,958 (2.4%) | 7,846 (2.5%) |
|  | 29 | 168,462 (2.4%) | 161,024 (2.4%) | 7,438 (2.4%) |
|  | 25 | 166,648 (2.3%) | 159,181 (2.3%) | 7,467 (2.4%) |
|  | 3 | 162,744 (2.3%) | 155,784 (2.3%) | 6,960 (2.2%) |
|  | 4 | 160,224 (2.2%) | 153,663 (2.2%) | 6,561 (2.1%) |
|  | 5 | 159,739 (2.2%) | 152,422 (2.2%) | 7,317 (2.3%) |
|  | 26 | 145,183 (2.0%) | 138,990 (2.0%) | 6,193 (2.0%) |
|  | 28 | 128,655 (1.8%) | 123,850 (1.8%) | 4,805 (1.5%) |
|  | 32 | 123,502 (1.7%) | 118,923 (1.7%) | 4,579 (1.5%) |
|  | 35 | 125,582 (1.8%) | 118,277 (1.7%) | 7,305 (2.3%) |
|  | 7 | 123,802 (1.7%) | 118,157 (1.7%) | 5,645 (1.8%) |
|  | 36 | 123,939 (1.7%) | 116,753 (1.7%) | 7,186 (2.3%) |
|  | 39 | 114,770 (1.6%) | 109,331 (1.6%) | 5,439 (1.7%) |
|  | 34 | 111,380 (1.6%) | 106,896 (1.6%) | 4,484 (1.4%) |
|  | 22 | 104,788 (1.5%) | 101,138 (1.5%) | 3,650 (1.2%) |
|  | 19 | 103,413 (1.4%) | 100,489 (1.5%) | 2,924 (0.9%) |
|  | 20 | 103,905 (1.5%) | 97,746 (1.4%) | 6,159 (2.0%) |
|  | 16 | 102,101 (1.4%) | 97,037 (1.4%) | 5,064 (1.6%) |
|  | 15 | 101,875 (1.4%) | 96,397 (1.4%) | 5,478 (1.7%) |
|  | 6 | 98,087 (1.4%) | 92,398 (1.3%) | 5,689 (1.8%) |
|  | 23 | 87,650 (1.2%) | 83,057 (1.2%) | 4,593 (1.5%) |
|  | 18 | 51,878 (0.7%) | 48,140 (0.7%) | 3,738 (1.2%) |
| **Clinical Classifications Software (CCS) diagnosis group^3^** | Combination of CCS categories with a low risk of death (<1%) within 30 days of admission | 1,370,815 (19.1%) | 1,360,228 (19.9%) | 10,587 (3.4%) |
|  | Cancer of bladder; Cancer of bone and connective tissue, Cancer of thyroid, Malignant neoplasm without specification of site; Cancer of brain and nervous system; Cancer of breast; Cancer of bronchus, lung; Cancer of cervix, Cancer of other female genital organs; Cancer of colon; Cancer of head and neck; Cancer of kidney and renal pelvis, Cancer of other urinary organs; Cancer of liver and intrahepatic bile duct; Cancer of oesophagus; Cancer of other GI organs, peritoneum; Cancer of ovary; Cancer of pancreas; Cancer of prostate, Cancer of testis, Cancer of other male genital organs; Cancer of rectum and anus; Cancer of stomach; Cancer of uterus; Cancer, other and unspecified primary, Maintenance chemotherapy; radiotherapy; Cancer, other respiratory and intrathoracic; Hodgkin's disease, Non-Hodgkin's lymphoma; Leukemias; Melanomas of skin, Other non-epithelial cancer of skin; Multiple myeloma; Neoplasms of unspecified nature or uncertain behavior, Nonmalignant breast conditions; Pathological fracture; Secondary malignancies | 786,440 (11.0%) | 741,437 (10.8%) | 45,003 (14.3%) |
|  | Nonspecific chest pain; Cardiac dysrhythmias; Coronary atherosclerosis and other heart disease; Pulmonary heart disease; Essential hypertension, Hypertension with complications and secondary hypertension; Peri-; endo-; and myocarditis; cardiomyopathy (except that caused by tuberculosis or sexually transmitted disease); Conduction disorders; Heart valve disorders; Cardiac arrest and ventricular fibrillation; Other and ill-defined heart disease; Other circulatory disease | 754,804 (10.5%) | 740,929 (10.8%) | 13,875 (4.4%) |
|  | Headache; including migraine, Cataract, Retinal detachments; defects; vascular occlusion; and retinopathy, Glaucoma, Blindness and vision defects, Inflammation; infection of eye (except that caused by tuberculosis or sexually transmitted disease), Other eye disorders, Otitis media and related conditions, Conditions associated with dizziness or vertigo, Other ear and sense organ disorders; Syncope; Epilepsy; convulsions; Other nervous system disorders; Malaise and fatigue; Multiple sclerosis, Other hereditary and degenerative nervous system conditions; Parkinson's disease; Paralysis, Late effects of cerebrovascular disease | 449,642 (6.3%) | 442,759 (6.5%) | 6,883 (2.2%) |
|  | Deficiency and other anemia, Acute posthemorrhagic anemia; Nutritional deficiencies, Disorders of lipid metabolism, Other nutritional; endocrine; and metabolic disorders; Diseases of white blood cells | 426,298 (6.0%) | 418,888 (6.1%) | 7,410 (2.4%) |
|  | Other connective tissue disease; Gout and other crystal arthropathies, Rheumatoid arthritis and related disease, Osteoarthritis, Acquired foot deformities, Other acquired deformities, Systemic lupus erythematosus and connective tissue disorders, Other bone disease and musculoskeletal deformities; Other non-traumatic joint disorders | 338,840 (4.7%) | 333,813 (4.9%) | 5,027 (1.6%) |
|  | Other, 30-day mortality risk is < overall risk (4.39%) | 317,911 (4.4%) | 308,617 (4.5%) | 9,294 (3.0%) |
|  | Esophageal disorders; Gastrointestinal hemorrhage | 267,221 (3.7%) | 262,222 (3.8%) | 4,999 (1.6%) |
|  | Pneumonia (except that caused by tuberculosis or sexually transmitted disease); Aspiration pneumonitis; food/vomitus | 310,904 (4.3%) | 254,193 (3.7%) | 56,711 (18.0%) |
|  | Chronic obstructive pulmonary disease and bronchiectasis; Cystic fibrosis, Other lower respiratory disease; Lung disease due to external agents | 222,646 (3.1%) | 209,607 (3.1%) | 13,039 (4.1%) |
|  | Acute bronchitis; Asthma; | 143,653 (2.0%) | 138,371 (2.0%) | 5,282 (1.7%) |
|  | Mental retardation, Senility and organic mental disorders; Alcohol-related mental disorders, Substance-related mental disorders, Affective disorders, Anxiety; somatoform; dissociative; and personality disorders; Other psychoses; Schizophrenia and related disorders, Preadult disorders, Other mental conditions, Personal history of mental disorder | 143,059 (2.0%) | 135,523 (2.0%) | 7,536 (2.4%) |
|  | Urinary tract infections; Genitourinary symptoms and ill-defined conditions | 137,471 (1.9%) | 130,216 (1.9%) | 7,255 (2.3%) |
|  | Other liver diseases; Biliary tract disease; Liver disease; alcohol-related; Pancreatic disorders (not diabetes) | 131,419 (1.8%) | 123,524 (1.8%) | 7,895 (2.5%) |
|  | Superficial injury; contusion; Open wounds of head; neck; and trunk; Crushing injury or internal injury; Fracture of upper limb; Intracranial injury; Other injuries & conditions due to external causes; Open wounds of extremities; Fracture of lower limb; Fracture of neck of femur (hip); Burns | 130,196 (1.8%) | 122,558 (1.8%) | 7,638 (2.4%) |
|  | Other, 30-day mortality risk is ≥ overall risk (4.39%) | 129,101 (1.8%) | 114,600 (1.7%) | 14,501 (4.6%) |
|  | Skin and subcutaneous tissue infections; Other inflammatory condition of skin, Chronic ulcer of skin, Other skin disorders | 117,667 (1.6%) | 114,570 (1.7%) | 3,097 (1.0%) |
|  | Acute cerebrovascular disease; Occlusion or stenosis of precerebral arteries, Other and ill-defined cerebrovascular disease, Transient cerebral ischemia; Coma; stupor; and brain damage | 130,620 (1.8%) | 113,804 (1.7%) | 16,816 (5.4%) |
|  | Covid-19, Influenza, Acute and chronic tonsillitis, Other upper respiratory infections, Other upper respiratory disease, Disorders of teeth and jaw, Diseases of mouth; excluding dental | 123,132 (1.7%) | 113,484 (1.7%) | 9,648 (3.1%) |
|  | Septicaemia (except in labour), Shock | 137,316 (1.9%) | 109,463 (1.6%) | 27,853 (8.9%) |
|  | Spondylosis; intervertebral disc disorders; other back problems, Osteoporosis; Joint disorders and dislocations; trauma-related, Spinal cord injury, Skull and face fractures, Other fractures, Sprains and strains | 105,597 (1.5%) | 103,277 (1.5%) | 2,320 (0.7%) |
|  | Acute myocardial infarction | 107,280 (1.5%) | 99,834 (1.5%) | 7,446 (2.4%) |
|  | Intestinal infection | 99,909 (1.4%) | 96,738 (1.4%) | 3,171 (1.0%) |
|  | Congestive heart failure; nonhypertensive | 101,752 (1.4%) | 89,326 (1.3%) | 12,426 (4.0%) |
|  | Phlebitis; thrombophlebitis and thromboembolism, Varicose veins of lower extremity, Hemorrhoids, Other disease of veins and lymphatics; Lymphadenitis, Gangrene; Peripheral and visceral atherosclerosis; Aortic and peripheral arterial embolism or thrombosis; Aortic; peripheral; and visceral artery aneurysms | 86,976 (1.2%) | 84,161 (1.2%) | 2,815 (0.9%) |
|  | Nausea and vomiting; Fluid and electrolyte disorders | 89,752 (1.3%) | 83,966 (1.2%) | 5,786 (1.8%) |

^1^ Truncated at the 99th percentile.

^2^ Defined as HIV, severe liver disease, or metastatic cancer in primary or secondary diagnosis codes

^3^ Deprivation indices were converted to a 0-100 scale, where 0 was the most deprived, and 100 was the least deprived.

^4^ The ‘other’ consultant specialties include geriatric medicine, endocrinology, diabetic medicine, acute internal medicine, respiratory/thoracic medicine, infectious diseases, neurology, and rheumatology (the rationale for their inclusion is outlined in **Figure S1**).

#### Supplementary Table S6: Time periods included for analyses of primary endpoints (A) antibiotic DDDs per site, and (B) 30-day mortality

A) Antibiotic DDDs per site

| **Site** | **Antibiotic data start date** | **Antibiotic data end date** | **Analysis start date** | **Reason for change** | **Analysis end date** | **Reason for change** |
| --- | --- | --- | --- | --- | --- | --- |
| 1 | Feb-16 | Sep-20 | Feb-16 |  | Sep-20 |  |
| 2 | Feb-16 | Oct-20 | Feb-16 |  | Oct-20 |  |
| 3 | Feb-16 | Oct-20 | N/A |  | N/A | Excluded, as the introduction of an electronic prescribing system in May 2018 led to implausible declines (>90%) in reported antibiotic DDDs, leaving too little post-implementation data (6 months) to estimate post-implementation trends. |
| 4 | Feb-16 | Aug-20 | Feb-16 |  | Aug-20 |  |
| 5 | Feb-16 | Sep-20 | Feb-16 |  | Sep-20 |  |
| 6 | Feb-16 | Oct-20 | Feb-16 |  | Sep-20 | October 2020 was excluded as admissions were not available for the full month. |
| 7 | Feb-16 | Oct-20 | Feb-16 |  | Jul-20 | No admissions data in October 2020. Aug-Sept 2020 were excluded as there were large declines (>60-90%) in admissions compared to all previous months. |
| 8 | Feb-16 | Oct-20 | Feb-16 |  | Oct-20 |  |
| 9 | Feb-16 | Oct-20 | Feb-16 |  | Sep-20 | October 2020 was excluded as admissions were not available for the full month. |
| 10 | Feb-16 | Oct-20 | Feb-16 |  | Sep-20 | October 2020 was excluded as admissions were not available for the full month. |
| 11 | Feb-16 | Oct-20 | Feb-16 |  | Sep-20 | October 2020 was excluded as there was a large decline (>70%) in admissions compared to all previous months. |
| 12 | Feb-16 | Oct-20 | Feb-16 |  | Oct-20 |  |
| 13 | Feb-16 | Sep-20 | Feb-16 |  | Sep-20 |  |
| 14 | Feb-16 | Sep-20 | Feb-16 |  | Sep-20 |  |
| 15 | Feb-16 | Oct-20 | Feb-17 | Admissions not available before Feb-2017 | Oct-20 |  |
| 16 | Feb-16 | Oct-20 | Feb-16 |  | Oct-20 |  |
| 17 | Feb-16 | Oct-20 | Feb-16 |  | Oct-20 |  |
| 18 | Feb-16 | Sep-20 | Feb-16 |  | Sep-20 |  |
| 19 | Feb-16 | Oct-20 | Feb-16 |  | Jun-20 | No admissions data in October 2020. Jul-Sept 2020 were excluded as there was a large decline (>70%) in admissions compared to all previous months. |
| 20 | Feb-16 | Oct-20 | Feb-16 |  | Sep-20 | No admissions data in October 2020. |
| 21 | Feb-16 | Oct-20 | Feb-16 |  | Sep-20 | October 2020 was excluded as admissions were not available for the full month. |
| 22^1^ | Feb-16 | Jul-20 | Feb-16 |  | Jul-20 |  |
| 23 | Apr-16 | Oct-20 | Apr-16 |  | Oct-20 |  |
| 24 | Feb-16 | Oct-20 | Feb-16 |  | Sep-20 | October 2020 was excluded as admissions were not available for the full month. |
| 25 | Feb-16 | Oct-20 | Feb-16 |  | Sep-20 | October 2020 was excluded as admissions were not available for the full month. |
| 26 | Feb-16 | Oct-20 | Feb-16 |  | Aug-20 | No admissions data in October 2020. September 2020 was excluded as there a large decline (>50%) in admissions compared to all previous months. |
| 27 | Feb-16 | Sep-20 | Feb-16 |  | Sep-20 |  |
| 28 | Jul-17 | Oct-20 | Jul-17 |  | Sep-20 | October 2020 was excluded as admissions were not available for the full month. |
| 29 | Feb-16 | Oct-20 | Feb-16 |  | Oct-20 |  |
| 30^1^ | Apr-16 | Oct-20 | Apr-16 |  | Oct-20 |  |
| 31 | Feb-16 | Oct-20 | Feb-16 |  | Oct-20 |  |
| 32 | Feb-16 | Oct-20 | Feb-16 |  | Oct-20 |  |
| 33 | Feb-16 | Oct-20 | Feb-16 |  | Sep-20 | October 2020 was excluded as admissions were not available for the full month. |
| 34 | Feb-16 | Oct-20 | Feb-16 |  | Sep-20 | No admissions data in October 2020. |
| 35 | Feb-16 | Oct-20 | Feb-16 |  | Sep-20 | October 2020 was excluded as admissions were not available for the full month. |
| 36 | Aug-16 | Sep-20 | Aug-16 |  | Sep-20 |  |
| 37 | Feb-16 | Oct-20 | Feb-16 |  | Oct-20 |  |
| 38 | Feb-16 | Oct-20 | Feb-16 |  | Oct-20 |  |
| 39 | Feb-16 | Sep-20 | Feb-16 |  | Aug-20 | Sept 2020 was excluded as admissions were not available for the full month. |

^1^ The site could only provide hospital-level antibiotic data, rather than ward-level data for acute/general medicine.

B) 30-day mortality

| **Site** | **Complete cases data start date** | **Complete cases data end date** | **30-day mortality analysis start date** | **Reason for change** | **30-day mortality analysis end date** | **Reason for change** | **Other covariate patterns** |
| --- | --- | --- | --- | --- | --- | --- | --- |
| 1 | 01-Feb-16 | 23-Oct-20 | 01-Feb-16 |  | 28-Sep-20 | See footnote^1^ |  |
| 2 | 01-Feb-16 | 31-Oct-20 | 01-Feb-16 |  | 30-Sep-20 | See footnote^1^ |  |
| 3 | 01-Feb-16 | 30-Sep-20 | 01-Feb-16 |  | 30-Sep-20 |  | Charlson comorbidity score increases gradually through 2016-2017. |
| 4 | 01-Feb-16 | 27-Oct-20 | 01-Feb-16 |  | 29-Sep-20 | See footnote^1^ |  |
| 5 | 01-Feb-16 | 31-Oct-20 | 01-Sep-16 | Patient admissions in the past year was calculated incorrectly in 2016; rates stabilise from September 2016 onwards. | 31-Oct-20 |  |  |
| 6 | 01-Feb-16 | 26-Oct-20 | 01-Feb-16 |  | 27-Sep-20 | See footnote^1^ |  |
| 7 | 01-Feb-16 | 30-Sep-20 | 01-Feb-16 |  | 31-Aug-20 | Incomplete ascertainment of death in September 2020. |  |
| 8 | 01-Feb-16 | 31-Oct-20 | 01-Feb-16 |  | 30-Sep-20 | Incomplete ascertainment of death in October 2020. | Charlson comorbidity score increases gradually through 2016-2017. |
| 9 | 01-Feb-16 | 20-Oct-20 | 01-Feb-16 |  | 22-Sep-20 | See footnote^1^ |  |
| 10 | 01-Feb-16 | 28-Oct-20 | 01-Oct-16 | Previous admissions rise by >10% in 2016; rates stabilise from October 2016 onwards. | 30-Sep-20 | See footnote^1^ |  |
| 11 | 01-Feb-16 | 30-Oct-20 | 01-Feb-16 |  | 30-Sep-20 | See footnote^1^ |  |
| 12 | 01-Feb-16 | 31-Oct-20 | 01-Feb-16 |  | 31-Oct-20 |  | Charlson comorbidity score increases gradually through 2016-2017. |
| 13 | 01-Feb-16 | 09-Oct-20 | 01-Apr-16 | Patient admissions in the past year changed >10% between February-April 2016; rates stabilise from April 2016 onwards. | 09-Sep-20 | See footnote^1^ | There appear to be changes in case-mix over time, as the proportion of day-case spells declines from 50-20% in 2016-19. |
| 14 | 01-Feb-16 | 31-Oct-20 | 01-Feb-16 |  | 31-Oct-20 |  |  |
| 15 | 01-Feb-17 | 31-Oct-20 | 01-Jul-17 | Patient admissions in the past year was calculated incorrectly in 2017; rates stabilise from July 2017 onwards. | 31-Oct-20 |  |  |
| 16 | 01-Feb-16 | 31-Oct-20 | 01-Feb-16 |  | 31-Oct-20 |  |  |
| 17 | 01-Feb-16 | 31-Oct-20 | 01-Feb-16 |  | 31-Oct-20 |  |  |
| 18 | 01-Feb-16 | 31-Oct-20 | 01-Feb-16 |  | 31-Oct-20 |  | In June 2017 the proportion of patients with a non-emergency admission increases from 10%-25% and this rate is sustained through October 2020. |
| 19 | 01-Feb-16 | 30-Sep-20 | 01-Feb-16 |  | 30-Jun-20 | Incomplete diagnostic coding after June 2020; also incomplete ascertainment of death after July 2020 |  |
| 20 | 01-Feb-16 | 30-Sep-20 | 01-Feb-16 |  | 28-Sep-20 | See footnote^1^ |  |
| 21 | 01-Feb-16 | 16-Oct-20 | 01-Feb-16 |  | 18-Sep-20 | See footnote^1^ |  |
| 22 | 01-Feb-16 | 01-Aug-20 | 01-Feb-16 |  | 30-Jun-20 | Incomplete ascertainment of death in July and August 2020. |  |
| 23 | 01-Feb-16 | 30-Oct-20 | 01-Feb-16 |  | 30-Sep-20 | See footnote^1^ |  |
| 24 | 01-Feb-16 | 20-Oct-20 | 01-Feb-16 |  | 20-Oct-20 |  |  |
| 25 | 01-Feb-16 | 15-Oct-20 | 01-Oct-16 | Patient admissions in the past year was calculated incorrectly in 2016; rates stabilise from October 2016 onwards. | 29-Sep-20 | See footnote^1^ | Charlson comorbidity score increases gradually through 2016-2019. |
| 26 | 01-Feb-16 | 30-Sep-20 | 01-Feb-16 |  | 30-Sep-20 |  | Charlson comorbidity score increases gradually through 2016-20, as do complex admissions in the past year. |
| 27 | 01-Feb-16 | 01-Oct-20 | 01-Feb-16 |  | 30-Sep-20 | See footnote^1^ | Charlson comorbidity score increases gradually through 2016-2020. |
| 28 | 01-Feb-16 | 29-Oct-20 | 01-Feb-16 |  | 30-Sep-20 | Incomplete diagnostic coding in October 2020. |  |
| 29 | 01-Feb-16 | 31-Oct-20 | 01-Feb-16 |  | 31-Oct-20 |  |  |
| 30 | 01-Feb-16 | 31-Oct-20 | 01-Feb-16 |  | 31-Oct-20 |  |  |
| 31 | 01-Feb-16 | 31-Oct-20 | 01-Feb-16 |  | 09-Oct-20 | See footnote^1^ |  |
| 32 | 01-Feb-16 | 31-Oct-20 | 01-Feb-16 |  | 16-Oct-20 | See footnote^1^ |  |
| 33 | 01-Feb-16 | 27-Oct-20 | 01-Feb-16 |  | 29-Sep-20 | See footnote^1^ |  |
| 34 | 01-Feb-16 | 30-Sep-20 | 01-Feb-16 |  | 30-Sep-20 |  |  |
| 35 | 01-Feb-16 | 24-Oct-20 | 01-Sep-16 | Patient admissions in the past year was calculated incorrectly in 2016; rates stabilise from September 2016 onwards. | 25-Sep-20 | See footnote^1^ |  |
| 36 | 01-Feb-16 | 18-Oct-20 | 01-Feb-16 |  | 18-Sep-20 | See footnote^1^ |  |
| 37 | 01-Feb-16 | 31-Oct-20 | 01-Feb-16 |  | 31-Oct-20 |  | Charlson comorbidity score increases gradually through 2016-2017. |
| 38 | 01-Feb-16 | 31-Oct-20 | 01-Feb-16 |  | 31-Oct-20 |  |  |
| 39 | 01-Feb-16 | 28-Sep-20 | 01-Feb-16 |  | 11-Sep-20 | See footnote^1^ |  |

^1^ Admissions beginning within 30 days of the last known death date were dropped to ensure each spell had the potential to be linked with the death registry. If the final month had admissions on just 1 day then these spells were also dropped as mortality estimates lacked precision.

#### Supplementary Table S7 CONSORT checklist items for reporting a stepped wedge cluster randomised trial

| **Topic** | **Item** | **Checklist item** | **Page no** |
| --- | --- | --- | --- |
| **Title and abstract** | | | |
|  | 1a | Identification as a stepped wedge cluster randomised trial in the title. | 1 |
|  | 1b | Structured summary of trial design, methods, results, and conclusions (see separate SW-CRT checklist for abstracts). | 3 |
| **Introduction** | | | |
| Background and objectives | 2a | Scientific background. Rationale for using a cluster design and rationale for using a stepped wedge design. | 5 |
|  | 2b | Specific objectives or hypotheses. | 6 |
| **Methods** | | | |
| Trial design | 3a | Description and diagram of trial design including definition of cluster, number of sequences, number of clusters randomised to each sequence, number of periods, duration of time between each step, and whether the participants assessed in different periods are the same people, different people, or a mixture. | S44 |
|  | 3b | Important changes to methods after trial commencement (such as eligibility criteria), with reasons. | N/A |
| Participants | 4a | Eligibility criteria for clusters and participants. | 6 |
|  | 4b | Settings and locations where the data were collected. | 6 |
| Interventions | 5 | The intervention and control conditions with sufficient details to allow replication, including whether the intervention was maintained or repeated, and whether it was delivered at the cluster level, the individual participant level, or both. | 6 |
| Outcomes | 6a | Completely defined prespecified primary and secondary outcome measures, including how and when they were assessed. | 6-7 |
|  | 6b | Any changes to trial outcomes after the trial commenced, with reasons. | N/A |
| Sample size | 7a | How sample size was determined. Method of calculation and relevant parameters with sufficient detail so the calculation can be replicated. Assumptions made about correlations between outcomes of participants from the same cluster. (see separate checklist for SW-CRT sample size items). | 7 and S7 |
|  | 7b | When applicable, explanation of any interim analyses and stopping guidelines. | N/A |
| **Randomisation** | | | |
| Sequence generation | 8a | Method used to generate the random allocation to the sequences of treatments. | 6 |
|  | 8b | Type of randomisation; details of any constrained randomisation or stratification, if used. | 6 |
| Allocation concealment mechanism | 9 | Specification that allocation was based on clusters; description of any methods used to conceal the allocation from the clusters until after recruitment. | 6 |
| Implementation | 10a | Who generated the randomisation schedule, who enrolled clusters, and who assigned clusters to sequences. | 6 |
|  | 10b | Mechanism by which individual participants were included in clusters for the purposes of the trial (such as complete enumeration, random sampling; continuous recruitment or ascertainment; or recruitment at a fixed point in time), including who recruited or identified participants. | 6 |
|  | 10c | Whether, from whom and when consent was sought and for what; whether this differed between treatment conditions. | 6 |
| Blinding | 11a | If done, who was blinded after assignment to sequences (eg, cluster level participants, individual level participants, those assessing outcomes) and how. | N/A |
|  | 11b | If relevant, description of the similarity of treatments. | N/A |
| Statistical methods | 12a | Statistical methods used to compare treatment conditions for primary and secondary outcomes including how time effects, clustering and repeated measures were taken into account. | 7 |
|  | 12b | Methods for additional analyses, such as subgroup analyses, sensitivity analyses, and adjusted analyses. | S3-7 |
| **Results** | | | |
| Participant flow  (diagram strongly recommended) | 13a | For each treatment condition or allocated sequence, the numbers of clusters and participants who were assessed for eligibility, were randomly assigned, received intended treatments, and were analysed for the primary outcome (see separate SW-CRT flow chart). | 7, S43 |
|  | 13b | For each treatment condition or allocated sequence, losses and exclusions for both clusters and participants with reasons. | 7 S43 |
| Recruitment | 14a | Dates defining the steps, initiation of intervention, and deviations from planned dates. Dates defining recruitment and follow-up for participants. | S43 |
|  | 14b | Why the trial ended or was stopped. | N/A |
| Baseline data | 15 | Baseline characteristics for the individual and cluster levels as applicable for each treatment condition or allocated sequence. | 8 |
| Numbers analysed | 16 | The number of observations and clusters included in each analysis for each treatment condition and whether the analysis was according to the allocated schedule. | 8-9 |
| Outcomes and estimation | 17a | For each primary and secondary outcome, results for each treatment condition, and the estimated effect size and its precision (such as 95% confidence interval); any correlations (or covariances) and time effects estimated in the analysis. | 8-10 |
|  | 17b | For binary outcomes, presentation of both absolute and relative effect sizes is recommended. | Yes 8-10 |
| Ancillary analyses | 18 | Results of any other analyses performed, including subgroup analyses and adjusted analyses, distinguishing prespecified from exploratory. | N/A |
| Harms | 19 | Important harms or unintended effects in each treatment condition (for specific guidance see CONSORT for harms). | Included in primary and secondary outcomes |
| **Discussion** | | | |
| Limitations | 20 | Trial limitations, addressing sources of potential bias, imprecision, and, if relevant, multiplicity of analyses. | 12 |
| Generalisability | 21 | Generalisability (external validity, applicability) of the trial findings. Generalisability to clusters or individual participants, or both (as relevant). | 12 |
| Interpretation | 22 | Interpretation consistent with results, balancing benefits and harms, and considering other relevant evidence. | 4 and 12 |
| **Other information** | | | |
| Registration | 23 | Registration number and name of trial registry. | 3 |
| Protocol | 24 | Where the full trial protocol can be accessed, if available. | 17 |
| Funding | 25 | Sources of funding and other support (such as supply of drugs), and the role of funders. | 3 |
| Research ethics review | 26 | Whether the study was approved by a research ethics committee, with identification of the review committee(s). Justification for any waiver or modification of informed consent requirements. | 6 |

#### Supplementary Table S8: Data cleaning steps used to derive complete cases for the analysis of 30-day mortality (co-primary endpoint), by site

| **NHS site** | **Admissions in raw data** | **Spells dropped that were unfinished at the close of the dataset** | | **Spells dropped that were missing age at admission** | | **Spells dropped that were missing Charlson score** | | **Spells dropped that were missing deprivation percentile** | | **Complete cases,**  **i.e., no missing data among factors included in 30-day mortality models**  **(co-primary endpoint)** | |
| --- | --- | --- | --- | --- | --- | --- | --- | --- | --- | --- | --- |
|  |  | **N** | **%** | **N** | **%** | **N** | **%** | **N** | **%** | **N** | **% of raw data dropped** |
| 1 | 275,770 | 0 | 0.0% | 21 | 0.0% | 848 | 0.3% | 454 | 0.2% | 274,447 | 0.5% |
| 2 | 337,981 | 0 | 0.0% | 127 | 0.0% | 10,765 | 3.2% | 23,908 | 7.3% | 303,181 | 10.3% |
| 3 | 168,534 | 0 | 0.0% | 13 | 0.0% | 112 | 0.1% | 5,665 | 3.4% | 162,744 | 3.4% |
| 4 | 182,005 | 390 | 0.2% | 1 | 0.0% | 16,589 | 9.1% | 3,917 | 2.4% | 161,108 | 11.5% |
| 5 | 192,814 | 9 | 0.0% | 26 | 0.0% | 398 | 0.2% | 11,196 | 5.8% | 181,185 | 6.0% |
| 6 | 100,085 | 291 | 0.3% | 1 | 0.0% | 267 | 0.3% | 332 | 0.3% | 99,194 | 0.9% |
| 7 | 128,296 | 0 | 0.0% | 4 | 0.0% | 4,010 | 3.1% | 370 | 0.3% | 123,912 | 3.4% |
| 8 | 288,364 | 19 | 0.0% | 6 | 0.0% | 2,842 | 1.0% | 4,758 | 1.7% | 280,739 | 2.6% |
| 9 | 208,546 | 0 | 0.0% | 40 | 0.0% | 15,814 | 7.6% | 12,340 | 6.4% | 180,352 | 13.5% |
| 10 | 305,391 | 24 | 0.0% | 3,880 | 1.3% | 1,205 | 0.4% | 31,966 | 10.6% | 268,316 | 12.1% |
| 11 | 366,201 | 711 | 0.2% | 4 | 0.0% | 5,038 | 1.4% | 2,038 | 0.6% | 358,410 | 2.1% |
| 12 | 210,983 | 0 | 0.0% | 0 | 0.0% | 71 | 0.0% | 613 | 0.3% | 210,299 | 0.3% |
| 13 | 272,791 | 30 | 0.0% | 0 | 0.0% | 8 | 0.0% | 584 | 0.2% | 272,169 | 0.2% |
| 14 | 174,050 | 16 | 0.0% | 2 | 0.0% | 115 | 0.1% | 2,113 | 1.2% | 171,804 | 1.3% |
| 15 | 116,969 | 5 | 0.0% | 1 | 0.0% | 1 | 0.0% | 2,393 | 2.0% | 114,569 | 2.1% |
| 16 | 122,629 | 0 | 0.0% | 0 | 0.0% | 4 | 0.0% | 20,524 | 16.7% | 102,101 | 16.7% |
| 17 | 359,703 | 0 | 0.0% | 47 | 0.0% | 225 | 0.1% | 1,999 | 0.6% | 357,432 | 0.6% |
| 18 | 65,345 | 0 | 0.0% | 1 | 0.0% | 0 | 0.0% | 13,466 | 20.6% | 51,878 | 20.6% |
| 19 | 110,551 | 0 | 0.0% | 2 | 0.0% | 4,253 | 3.8% | 1,570 | 1.5% | 104,726 | 5.3% |
| 20 | 104,142 | 0 | 0.0% | 0 | 0.0% | 15 | 0.0% | 122 | 0.1% | 104,005 | 0.1% |
| 21 | 257,639 | 202 | 0.1% | 2 | 0.0% | 4,152 | 1.6% | 8,365 | 3.3% | 244,918 | 4.9% |
| 22 | 106,705 | 0 | 0.0% | 0 | 0.0% | 0 | 0.0% | 545 | 0.5% | 106,160 | 0.5% |
| 23 | 89,382 | 32 | 0.0% | 0 | 0.0% | 336 | 0.4% | 220 | 0.2% | 88,794 | 0.7% |
| 24 | 188,432 | 3 | 0.0% | 1 | 0.0% | 955 | 0.5% | 812 | 0.4% | 186,661 | 0.9% |
| 25 | 222,189 | 174 | 0.1% | 0 | 0.0% | 1,758 | 0.8% | 25,722 | 11.7% | 194,535 | 12.4% |
| 26 | 147,997 | 0 | 0.0% | 0 | 0.0% | 1,968 | 1.3% | 846 | 0.6% | 145,183 | 1.9% |
| 27 | 172,966 | 0 | 0.0% | 0 | 0.0% | 51 | 0.0% | 447 | 0.3% | 172,468 | 0.3% |
| 28 | 134,346 | 3 | 0.0% | 2 | 0.0% | 131 | 0.1% | 3,791 | 2.8% | 130,419 | 2.9% |
| 29 | 180,449 | 0 | 0.0% | 4 | 0.0% | 10 | 0.0% | 11,973 | 6.6% | 168,462 | 6.6% |
| 30 | 291,008 | 10 | 0.0% | 10 | 0.0% | 0 | 0.0% | 7,642 | 2.6% | 283,346 | 2.6% |
| 31 | 313,459 | 311 | 0.1% | 18 | 0.0% | 1,753 | 0.6% | 19,149 | 6.1% | 292,228 | 6.8% |
| 32 | 129,503 | 5 | 0.0% | 17 | 0.0% | 255 | 0.2% | 4,741 | 3.7% | 124,485 | 3.9% |
| 33 | 229,616 | 0 | 0.0% | 2 | 0.0% | 1,456 | 0.6% | 1,645 | 0.7% | 226,513 | 1.4% |
| 34 | 113,528 | 19 | 0.0% | 97 | 0.1% | 30 | 0.0% | 2,002 | 1.8% | 111,380 | 1.9% |
| 35 | 150,668 | 318 | 0.2% | 4 | 0.0% | 1,040 | 0.7% | 5,684 | 3.8% | 143,622 | 4.7% |
| 36 | 127,855 | 512 | 0.4% | 5 | 0.0% | 1,323 | 1.0% | 605 | 0.5% | 125,410 | 1.9% |
| 37 | 360,930 | 0 | 0.0% | 54 | 0.0% | 655 | 0.2% | 2,141 | 0.6% | 358,080 | 0.8% |
| 38 | 232,434 | 1 | 0.0% | 3 | 0.0% | 45 | 0.0% | 997 | 0.4% | 231,388 | 0.5% |
| 39 | 117,443 | 0 | 0.0% | 0 | 0.0% | 1,578 | 1.3% | 598 | 0.5% | 115,267 | 1.9% |
| **Total** | **7,657,699** | **3,085** | **0.0%** | **4,395** | **0.1%** | **80,076** | **1.0%** | **238,253** | **3.1%** | **7,331,890** | **4.3%** |

#### **Supplementary Table S9: Summary of trends in outcomes pre- and post-implementation of the ARK intervention**

| Outcome | Model used to derive site-specific estimates | Analysis  type | Random effects meta-analysis | |
| --- | --- | --- | --- | --- |
|  |  |  | Pre-implementation trend: OR, IRR, or SHR (95% CI), p-value^1^ | Post-implementation trend: OR, IRR, or SHR (95% CI), p-value^1^ |
| Total DDD/admission | Adjusted negative binomial model, without COVID adjustment, 38 sites **(Figure S7)** | 1 | 0.99 (0.97, 1.01), p=0.205 | 0.94 (0.90,0.97), p=0.001 |
| Total DDD/admission | Adjusted negative binomial model, without COVID adjustment, 38 sites | 2 | 0.99 (0.97, 1.01), p=0.233 | 0.98 (0.94, 1.02), p=0.345 |
| Total DDD/admission | Adjusted negative binomial model, excluding all follow-up after March 2020, 26 sites | 3 | 0.98 (0.96, 1.01), p=0.138 | 0.93 (0.89, 0.98), p=0.004 |
| Total DDD/admission | Unadjusted negative binomial model, with COVID adjustment, 38 sites | 4 | 0.99 (0.97, 1.01), p=0.261 | 0.92 (0.89, 0.96), p<0.0001 |
| Total DDD/admission | Adjusted negative binomial model, with COVID adjustment, main trial sites only, 36 sites | Sensi-tivity | 0.99 (0.97, 1.01), p=0.344 | 0.93 (0.90, 0.97), p=0.001 |
| Total DDD/bed-day | Adjusted negative binomial model, with COVID adjustment, 38 sites | 1 | 1.00 (0.98, 1.02), p=0.914 | 0.96 (0.92, 0.99), p=0.013 |
| Broad-spectrum DDD/ admission | Adjusted negative binomial model, with COVID adjustment, 38 sites | 1 | 1.00 (0.97, 1.03), p=0.978 | 0.97 (0.93, 1.01), p=0.188 |
| Narrow-spectrum DDD/ admission | Adjusted negative binomial model, with COVID adjustment, 38 sites | 1 | 0.98 (0.95, 1.00), p=0.090 | 0.93 (0.89, 0.96), p=0.0002 |
| Access DDD/admission | Adjusted negative binomial model, with COVID adjustment, 38 sites | 1 | 0.99 (0.96, 1.02), p=0.530 | 0.94 (0.90, 0.98), p=0.002 |
| Access or Watch DDD/ admission^2^ | Adjusted negative binomial model, with COVID adjustment, 38 sites | 1 | 0.92 (0.89, 0.95), p<0.0001 | 0.92 (0.88, 0.98), p=0.005 |
| Watch DDD/admission | Adjusted negative binomial model, with COVID adjustment, 38 sites | 1 | 1.07 (1.03, 1.11), p=0.001 | 0.95 (0.91, 1.00), p=0.036 |
| Reserve-spectrum DDD/ admission | Adjusted negative binomial model, with COVID adjustment, 38 sites | 1 | 0.92 (0.87, 0.97), p=0.001 | 0.99 (0.92, 1.05), p=0.681 |
| Parenteral DDD/admission | Adjusted negative binomial model, with COVID adjustment, 38 sites | 1 | 0.98 (0.96, 1.00), p=0.113 | 0.97 (0.94, 1.01), p=0.127 |
| Oral DDD/admission | Adjusted negative binomial model, with COVID adjustment, 38 sites | 1 | 0.99 (0.97, 1.01), p=0.375 | 0.92 (0.89, 0.96), p=0.0002 |
| Carbapenem DDD/admission | Adjusted negative binomial model, with COVID adjustment, 37 sites | 1 | 0.86 (0.81, 0.90), p<0.0001 | 0.96 (0.89, 1.03), p=0.236 |
| 30-day mortality | Adjusted logit model, with COVID adjustment, 39 sites **(Figure S14)** | 1 | 0.96 (0.95, 0.98), p<0.0001 | 0.99 (0.97, 1.01), p=0.260 |
| 30-day mortality | Adjusted logit model, without COVID adjustment, 39 sites | 2 | 0.97 (0.95,0.98), p<0.0001 | 1.13 (1.10,1.17), p<0.0001 |
| 30-day mortality | Adjusted logit model, excluding all follow-up from March 2020, 27 sites | 3 | 0.95 (0.93, 0.97) p<0.0001 | 0.95 (0.93, 0.97) p<0.0001 |
| 30-day mortality | Unadjusted logit model, with COVID adjustment and seasonal effect, 39 sites | 4 | 0.98 (0.97, 1.00), p=0.042 | 1.04 (1.01, 1.07), p=0.003 |
| 30-day mortality | Adjusted logit model, with COVID adjustment, excluding admissions to cardiology, rheumatology, haematology and neurology, 39 sites | Sensi-tivity | 0.96 (0.95, 0.98), p<0.0001 | 0.98 (0.96,1.01), p=0.163 |
| 30-day mortality | Adjusted logit model, with COVID adjustment, main trial sites only, 36 sites | Sensi-tivity | 0.96 (0.95, 0.98), p<0.0001 | 0.99 (0.97, 1.02), p=0.547 |
| 90-day mortality | Adjusted logit model, with COVID adjustment, 39 sites | 1 | 0.97 (0.95, 0.99), p=0.001 | 1.00 (0.97, 1.03), p=0.855 |
| 90-day mortality | Adjusted logit model, excluding follow-up from March 2020, 27 sites | 3 | 0.96 (0.93, 0.98), p=0.0001 | 0.99 (0.95, 1.03), p=0.615 |
| ICU admission | Adjusted logit model, with COVID adjustment, 36 sites | 1 | 0.96 (0.92, 1.00), p=0.037 | 0.92 (0.87, 0.96), p=0.001 |
| 30-day emergency readmission to hospital | Adjusted logit model, with COVID adjustment, 38 sites | 1 | 1.01 (1.00, 1.03), p=0.142 | 1.00 (0.97, 1.02), p=0.768 |
| 30-day emergency readmission to hospital | Adjusted competing risks model, with COVID adjustment, 38 sites | Sensi-tivity | 1.01 (1.00, 1.03), p=0.156 | 1.00 (0.98, 1.02), p=0.695 |
| *C difficile* infection within 90 days of admission | Adjusted logit model, without COVID adjustment, 38 sites | 1 | 0.98 (0.92, 1.04), p=0.464 | 1.08 (0.98, 1.20), p=0.137 |
| *C difficile* colonisation within 90 days of admission | Adjusted logit model, without COVID adjustment, 34 sites | 1 | 0.93 (0.89, 0.98), p=0.003 | 0.91 (0.82, 1.00), p=0.049 |
| Length of hospital stay (days) | Adjusted competing risks model, with COVID adjustment, 38 sites | 1 | 1.01 (1.00, 1.02), p=0.006 | 1.01 (1.00, 1.02), p=0.001 |

^1^ OR = Odds Ratio (logistic regression models), IRR = Incident Rate Ratio (negative binomial models), SHR = Subdistribution Hazard Ratio (competing risks models).

^2^ Antibiotics in this category may be considered either Access or Watch depending on indication. Since indication was unknown, they were analysed separately (**Table S2**).

See **Table S3** for intervention effects.

### **Supplementary Figures**

#### Supplementary Figure S1: Data included in the co-primary endpoint analysis in each site compared with randomised and actual implementation; 30-day mortality (A) and total antibiotic DDD/admission (B)

| **A** | 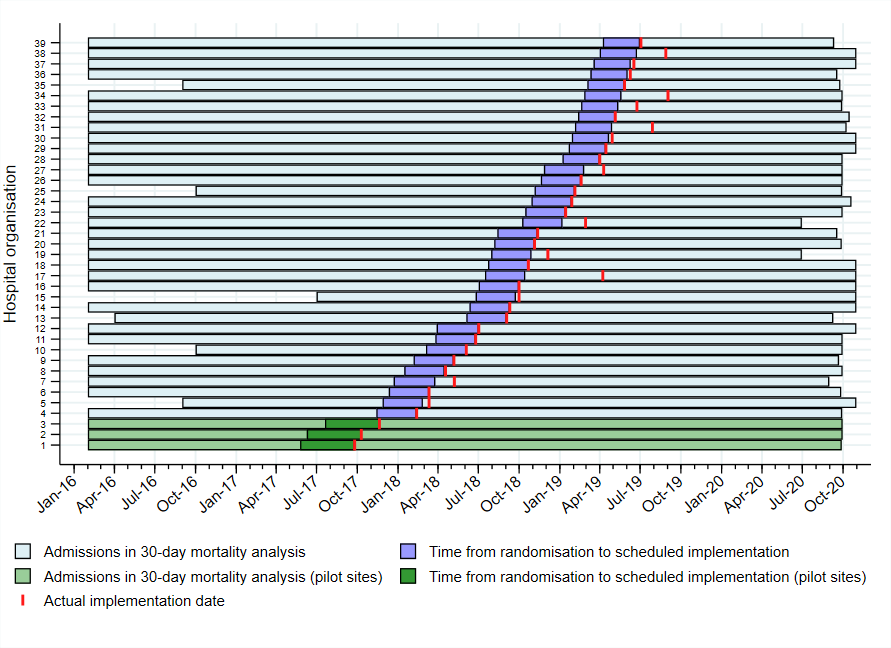 |
| --- | --- |
| **B** | 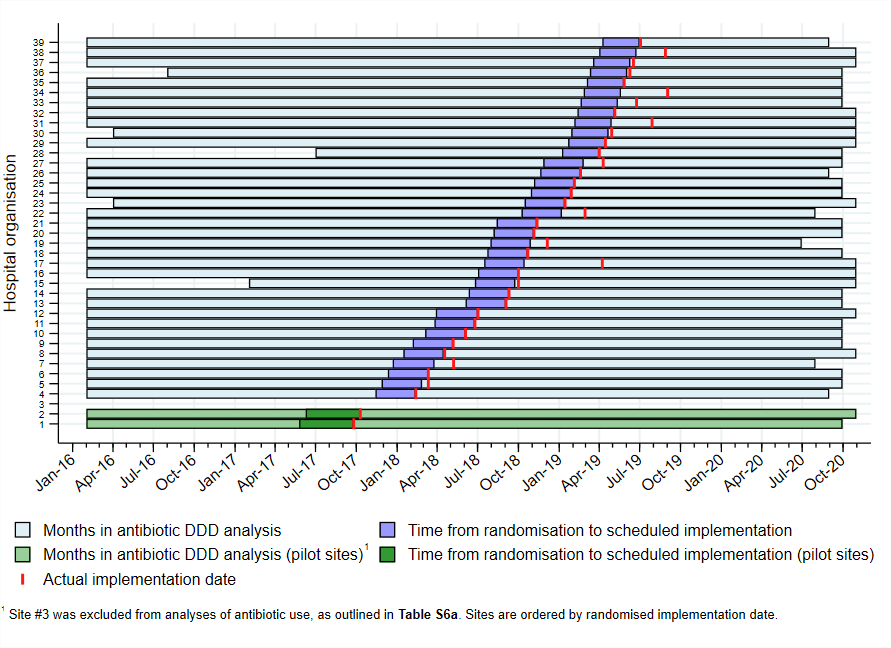 |

#### Supplementary Figure S2: Study population

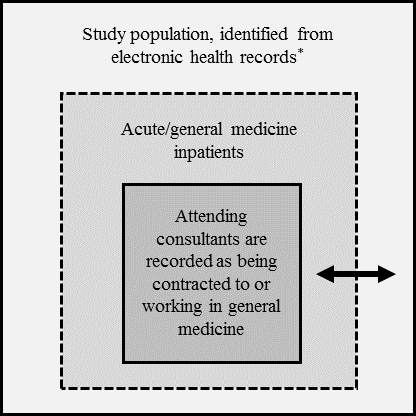

Note: The targeted study population included acute/general medical admissions managed by hospital care providers targeted by the ARK intervention (middle square). The study aimed to identify this population from electronic health records which do not record this directly, and therefore used the specialty codes of consultants attending to admitted patients, using records from the first and second episode of each spell. Patients cared for by consultants contracted to or working in general medicine (inner square) are likely to be a subset of the desired study population, as there is wide variation in the configuration of acute medical services across hospital organisations, and care pathways often differ within and across hospital sites depending on the local practice or time of day.^4^ To account for variation in coding practices across hospital organisations, a broad list of specialty codes was selected to reflect the specialties that are most often used to admit adult general medicine inpatients, including: acute/general medicine, gastroenterology, endocrinology, haematology, diabetic medicine, cardiology, acute internal medicine, respiratory/thoracic medicine, infectious diseases, neurology, rheumatology, and geriatric medicine (outer square). This definition was selected to be highly sensitive (i.e., it should include all patients cared for by healthcare professionals exposed to the ARK intervention). However, the specificity of this definition will vary by site (indicated by the arrow), depending on local coding practices and care pathways.

#### Supplementary Figure S3: Data cleaning steps used to derive analytic cohort for the analysis of 30-day mortality (co-primary endpoint)

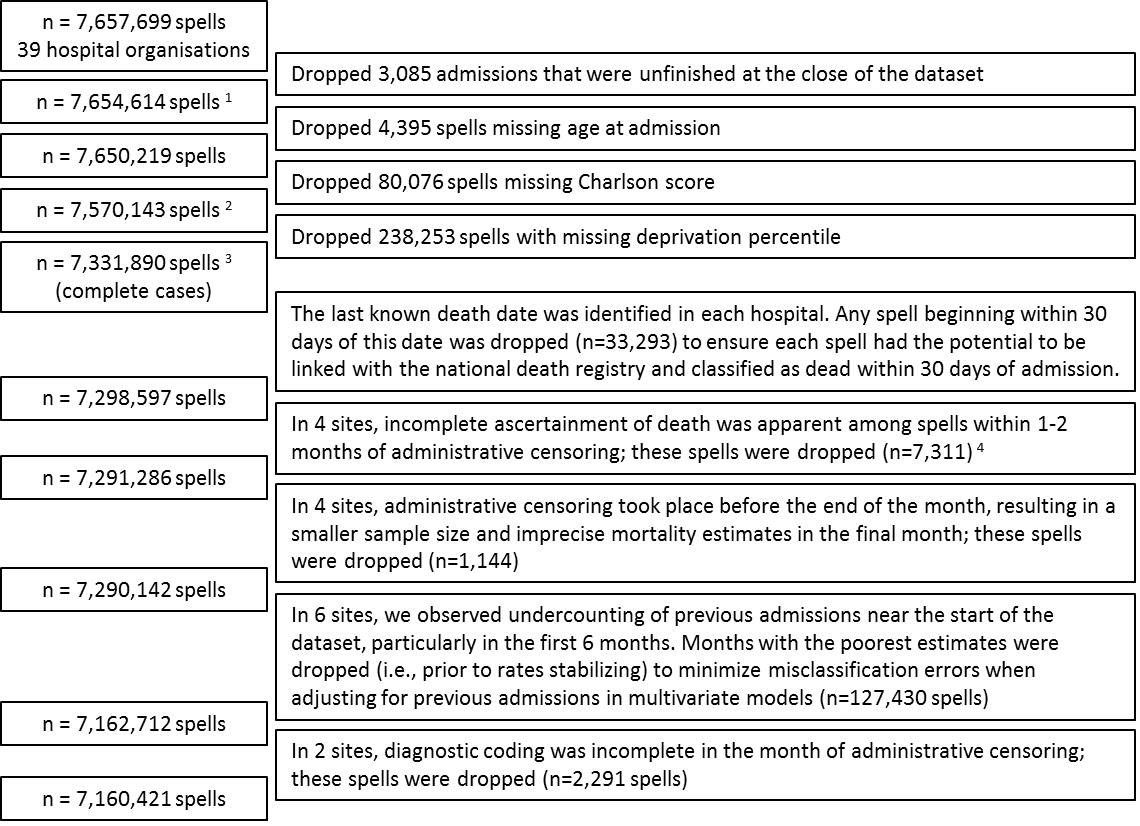

^1^ Spells reported to be in a patient >114 years of age were set to missing (n=138). When possible, missing age was imputed from the previous spell (n=25) or subsequent spell (n=17).

^2^ When possible, missing deprivation percentile (n=266,583; 3.5%) was imputed using the value found in the nearest spell (n=29,376), and missing ethnicity (n=1,333,180; 17.6%) was imputed from previous or subsequent spells (n=23,039).

^3^ Spells with missing sex (n=931) were set to the mode (female), as were spells with missing admission method (n=124), admission source (n=7,298), and patient classification (n=3,652). The remaining spells had no missing data among the factors included in 30-day mortality models (co-primary endpoint).

^4^ Linkage with the national death registry was judged to be incomplete as these spells showed 30-day mortality near 0% or below the lower 95% confidence bound of all months in the previous 4 years.

#### Supplementary Figure S4: Baseline antibiotic use in the 12 months before implementation; top 10 most commonly used antibiotics as a share of total DDDs (A), **WHO’s 2019 AWaRe categories as a share of total DDDs (B), monthly variation in DDDs/admission by antibiotic category (C) (D)**

| **A** | | 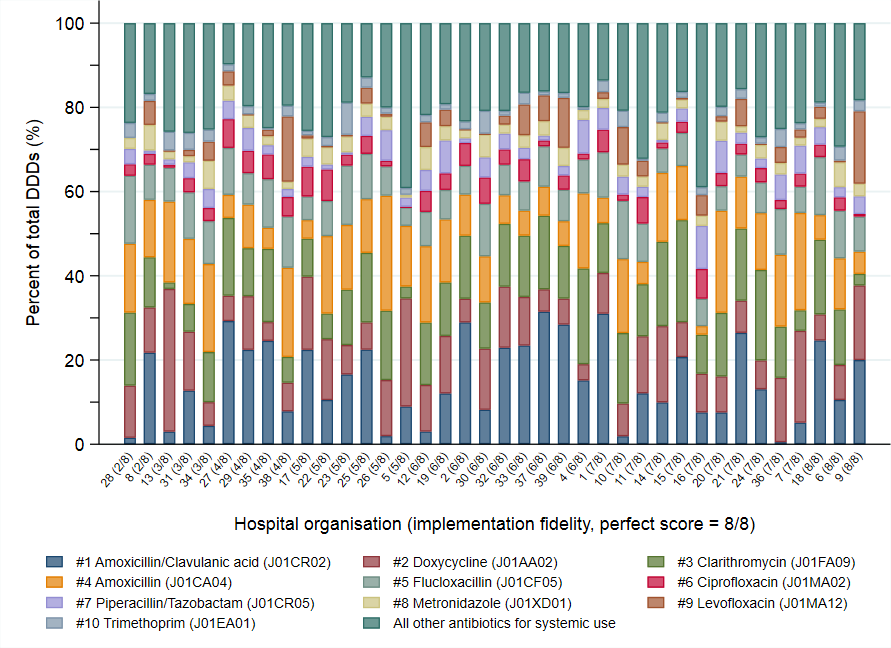 |
| --- | --- | --- |
| **B** | 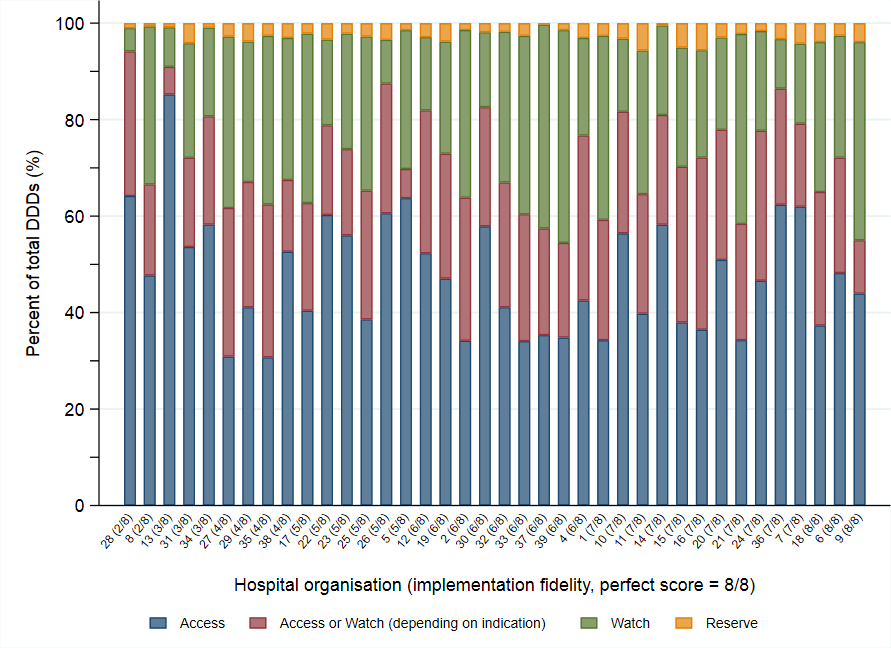 | |

| **C** | **Baseline antibiotic use (38 sites)**  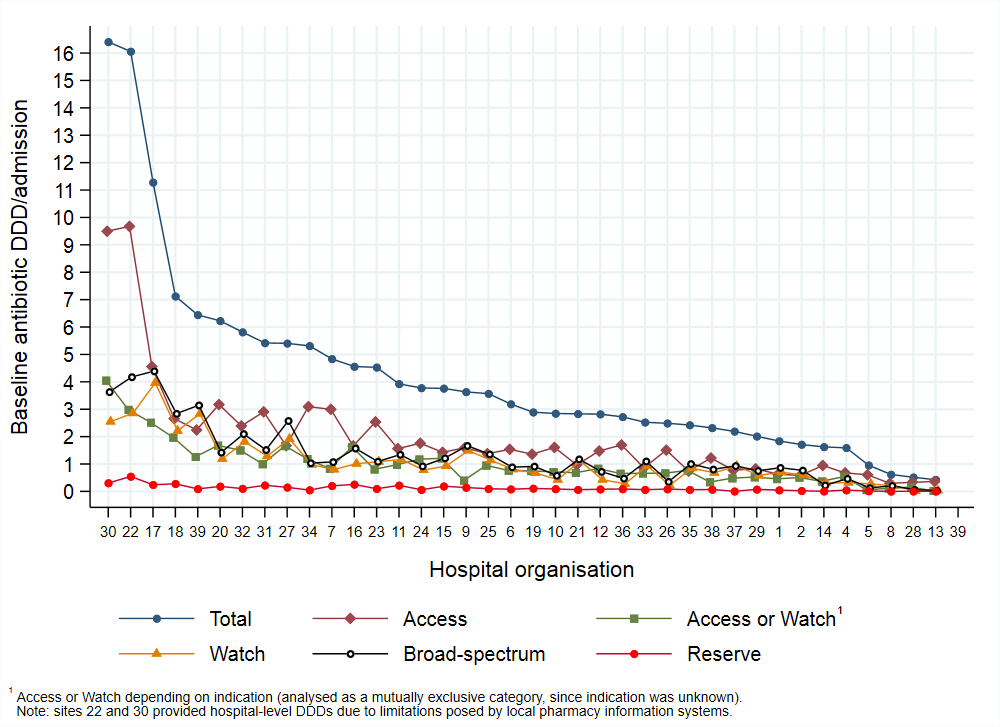 |
| --- | --- |
| **D** | **Baseline antibiotic use (38 sites)** 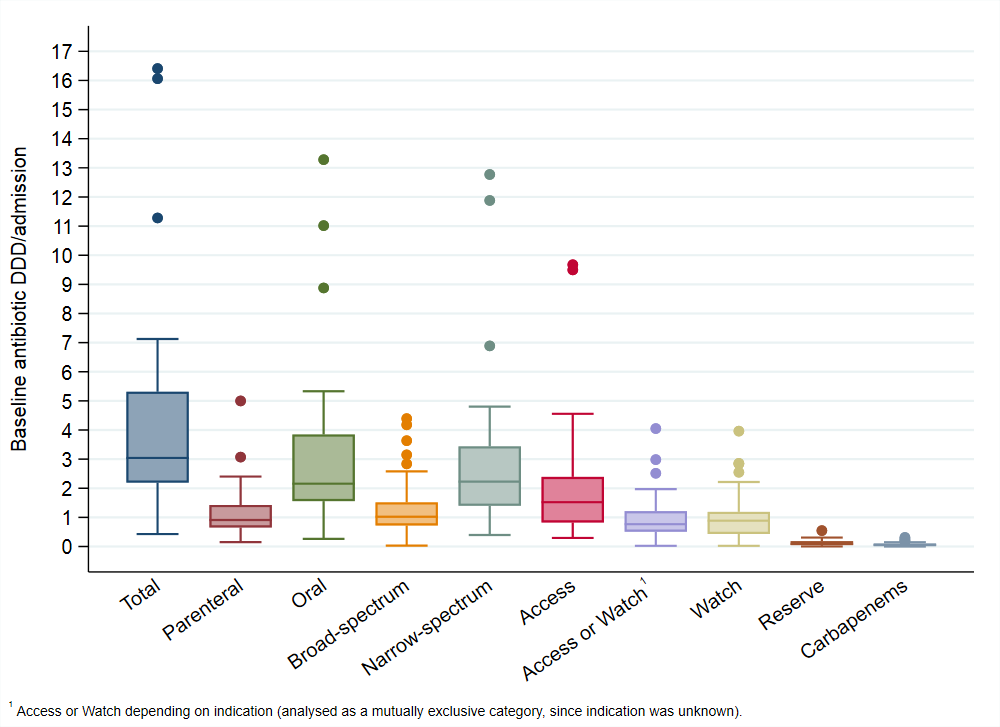 |

#### Supplementary Figure S5: overall antibiotic DDDs/admission and 30-day mortality over time by site

See separate file

#### Supplementary Figure S6: Intervention effects on total antibiotic DDDs/admission

Spearman's rho: -0.088, p=0.599
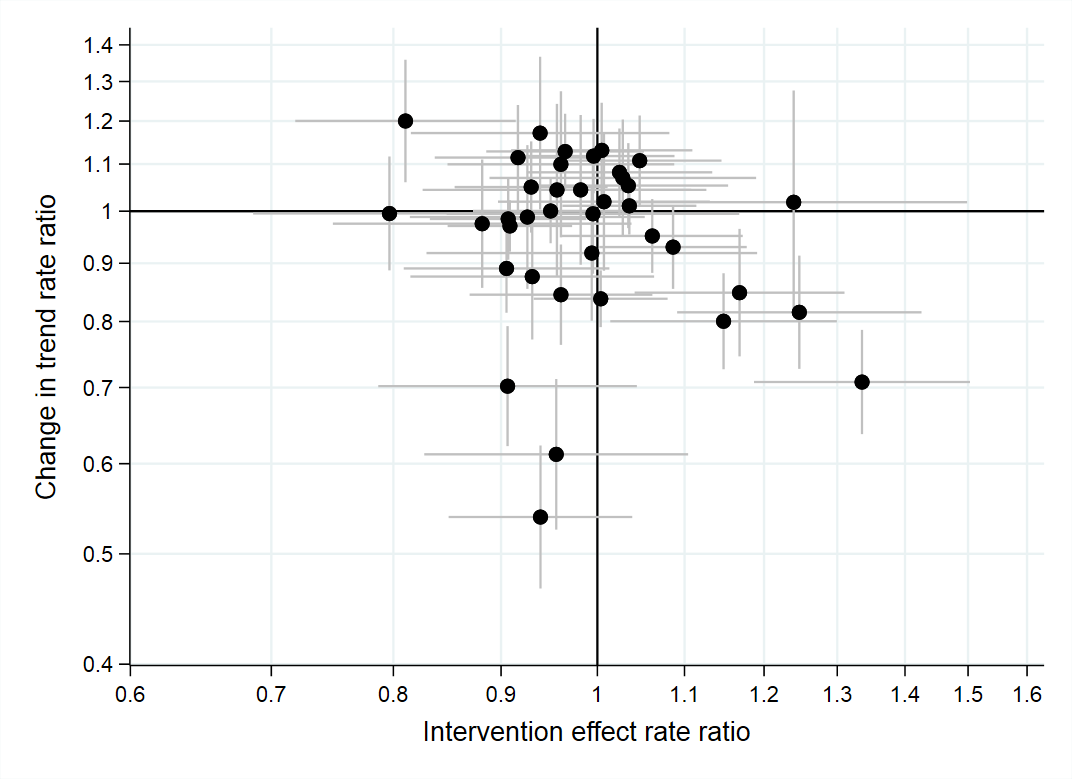

#### Supplementary Figure S7: Total antibiotic DDDs/admission (co-primary endpoint); change over time pre- (A) and post- implementation (B)

| **A** | **Pre-implementation trend (per year)**  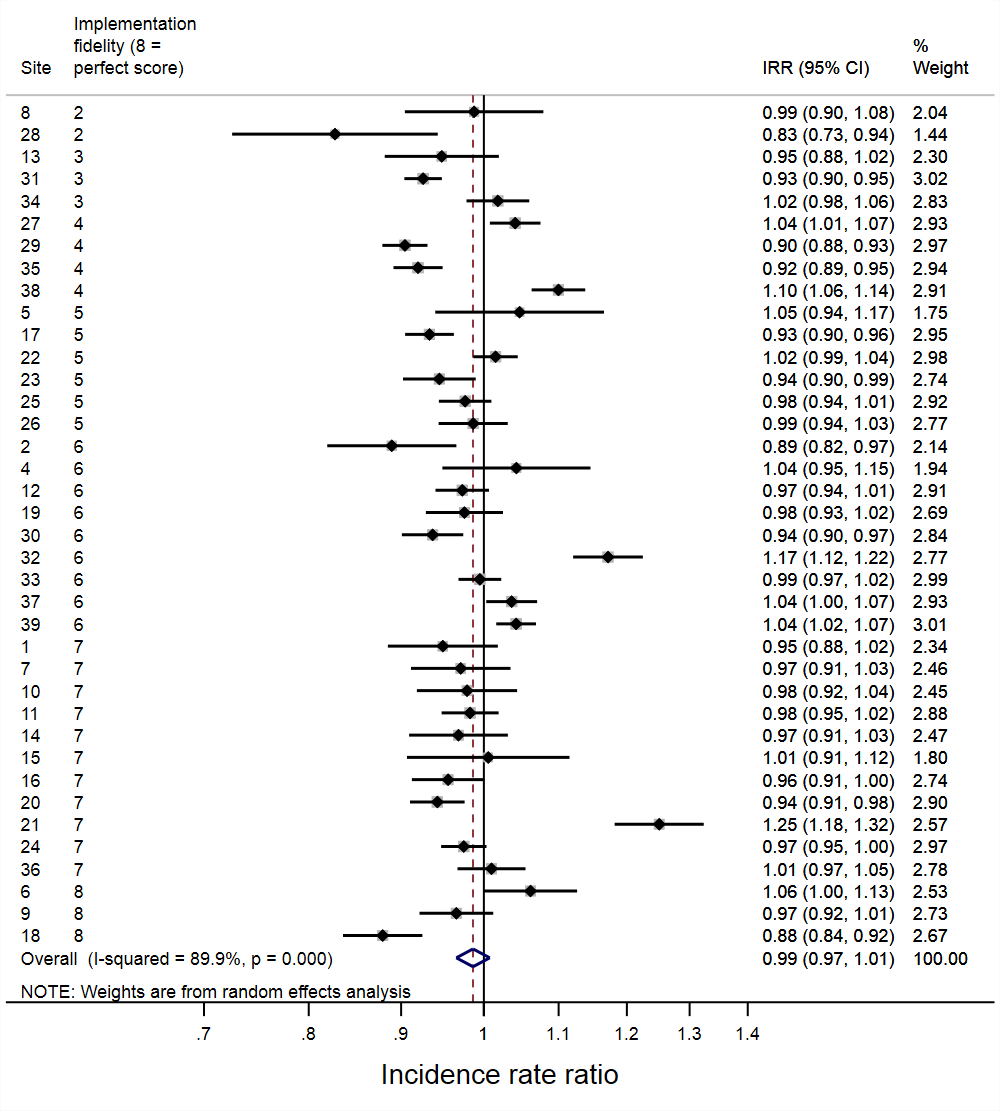 | **B** | **Post-implementation trend (per year)**  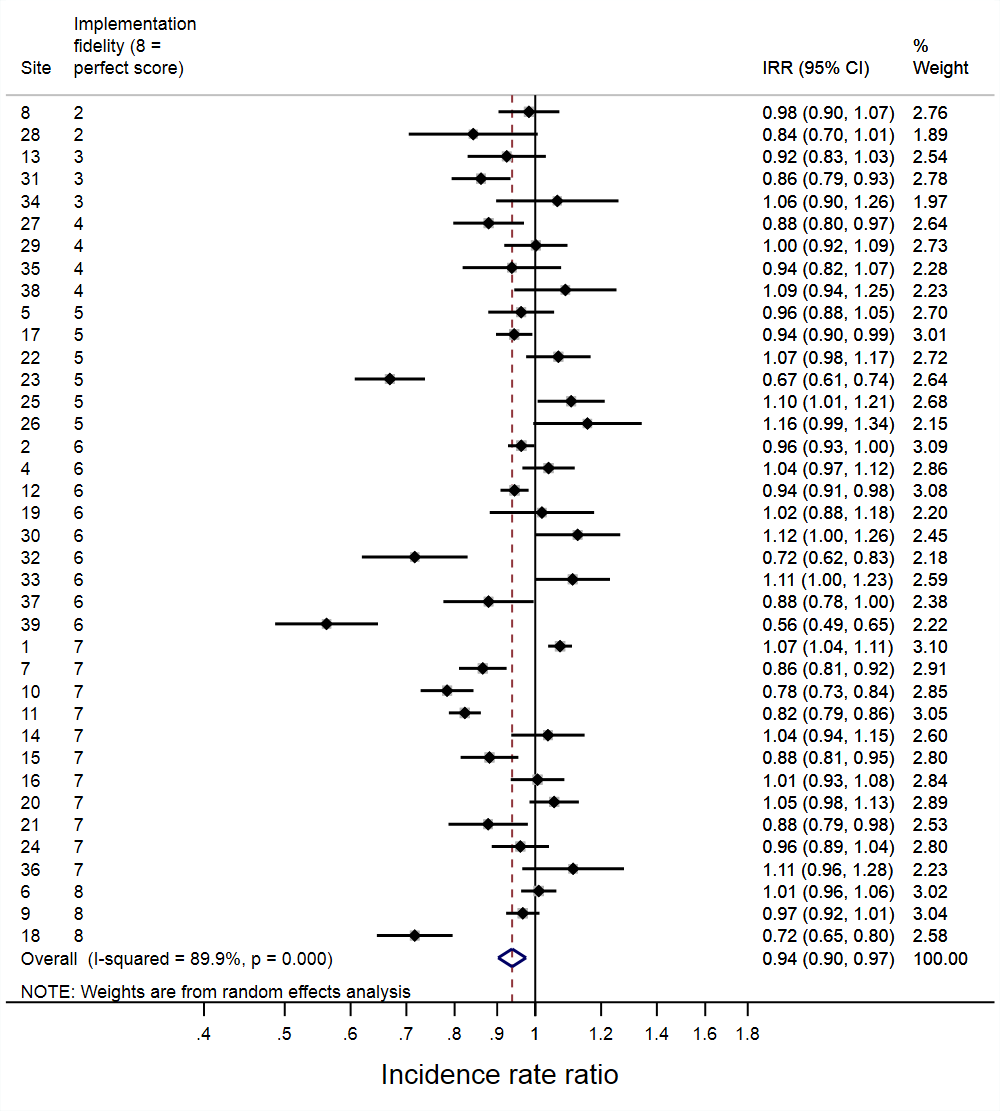 |
| --- | --- | --- | --- |

#### Supplementary Figure S8: Total antibiotic DDDs/bed-day (secondary outcome); **immediate impact of implementation (A), change in trend post vs pre-implementation (B)**

| **A** | **Immediate implementation effect**  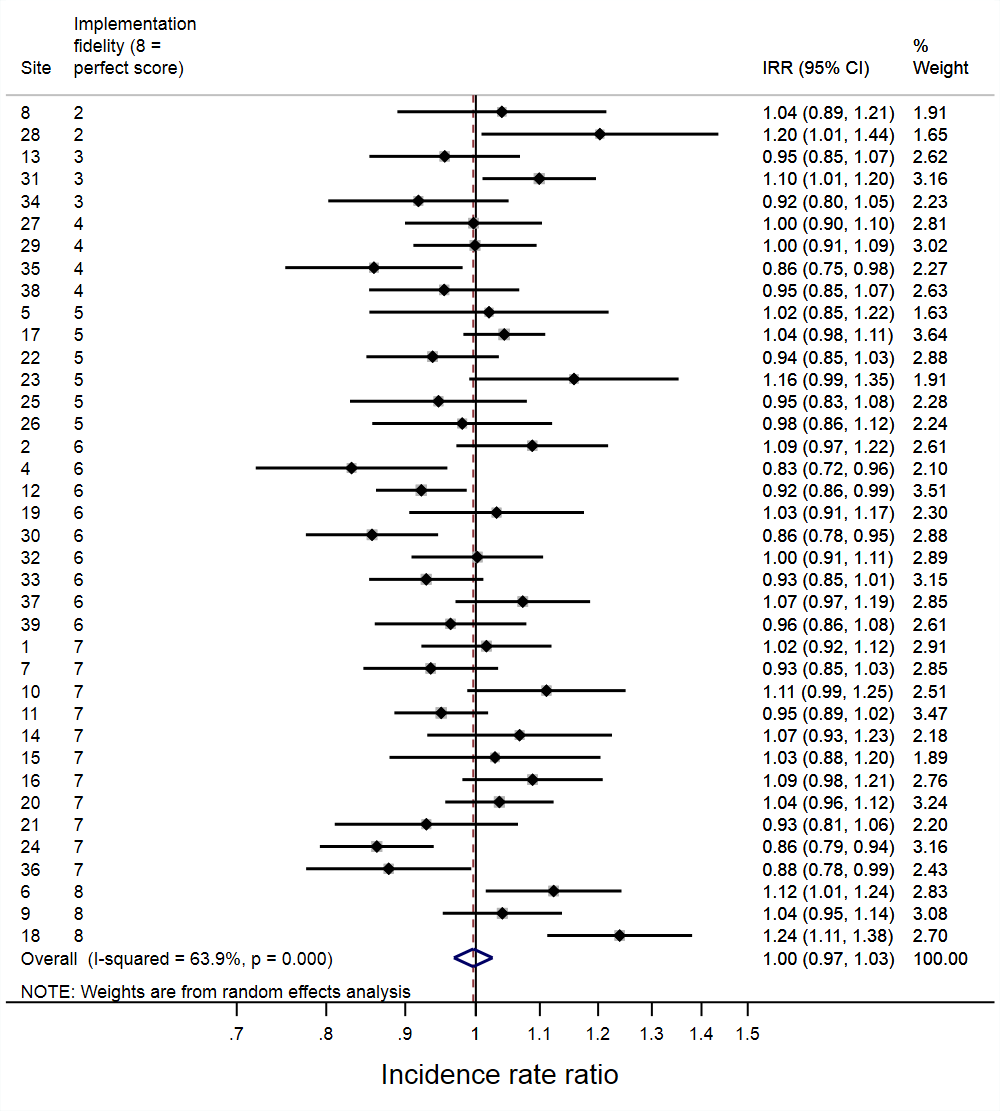 | **B** | **Change in trend post vs pre-implementation**  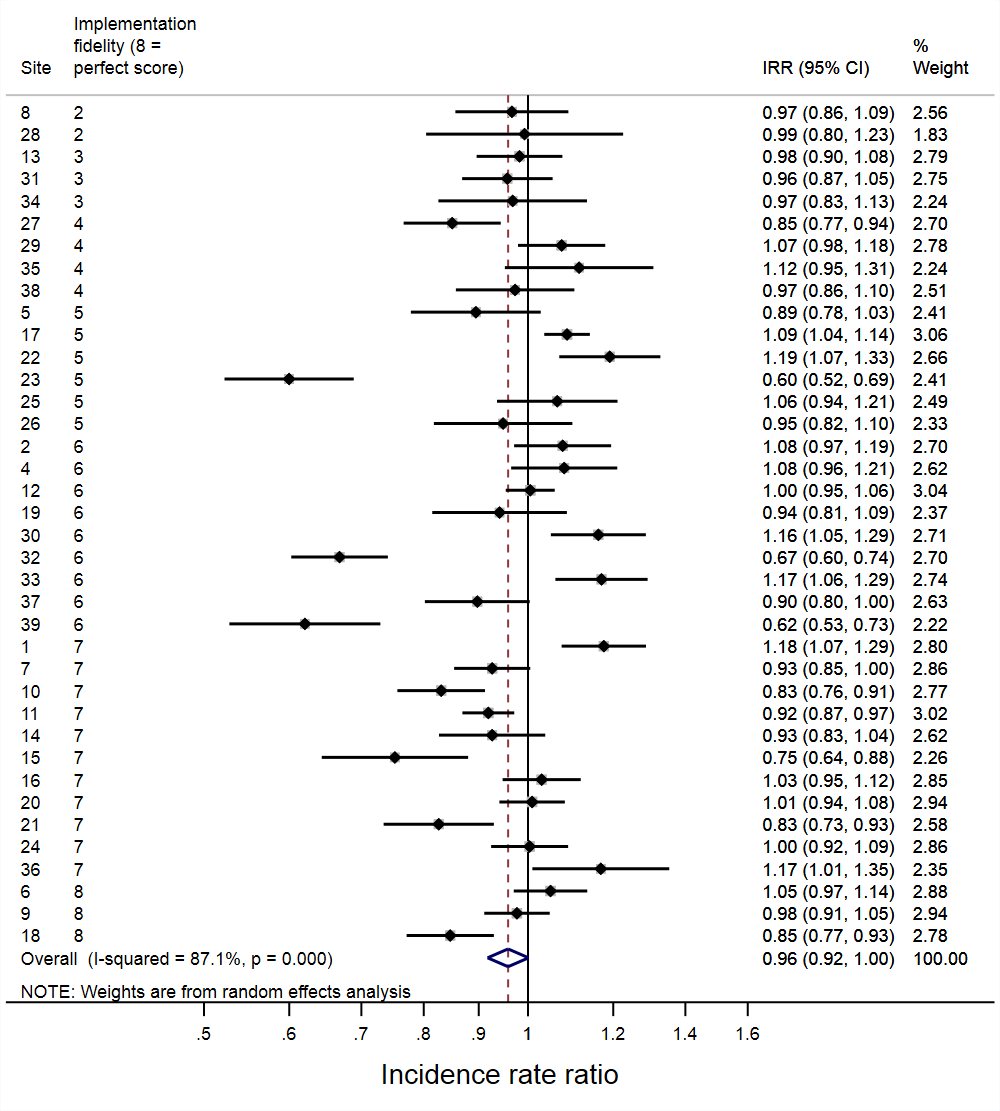 |
| --- | --- | --- | --- |

#### Supplementary Figure S9: Antibiotic DDD/admission classified by spectrum of activity (secondary outcome); **immediate impact of implementation broad spectrum antibiotics (A), change in trend post vs pre-implementation broad spectrum antibiotics (B), immediate impact of implementation narrow spectrum antibiotics (C), change in trend post vs pre-implementation narrow spectrum antibiotics (D)**

| **A** | **Broad spectrum DDD/admission**  **Immediate implementation effect**  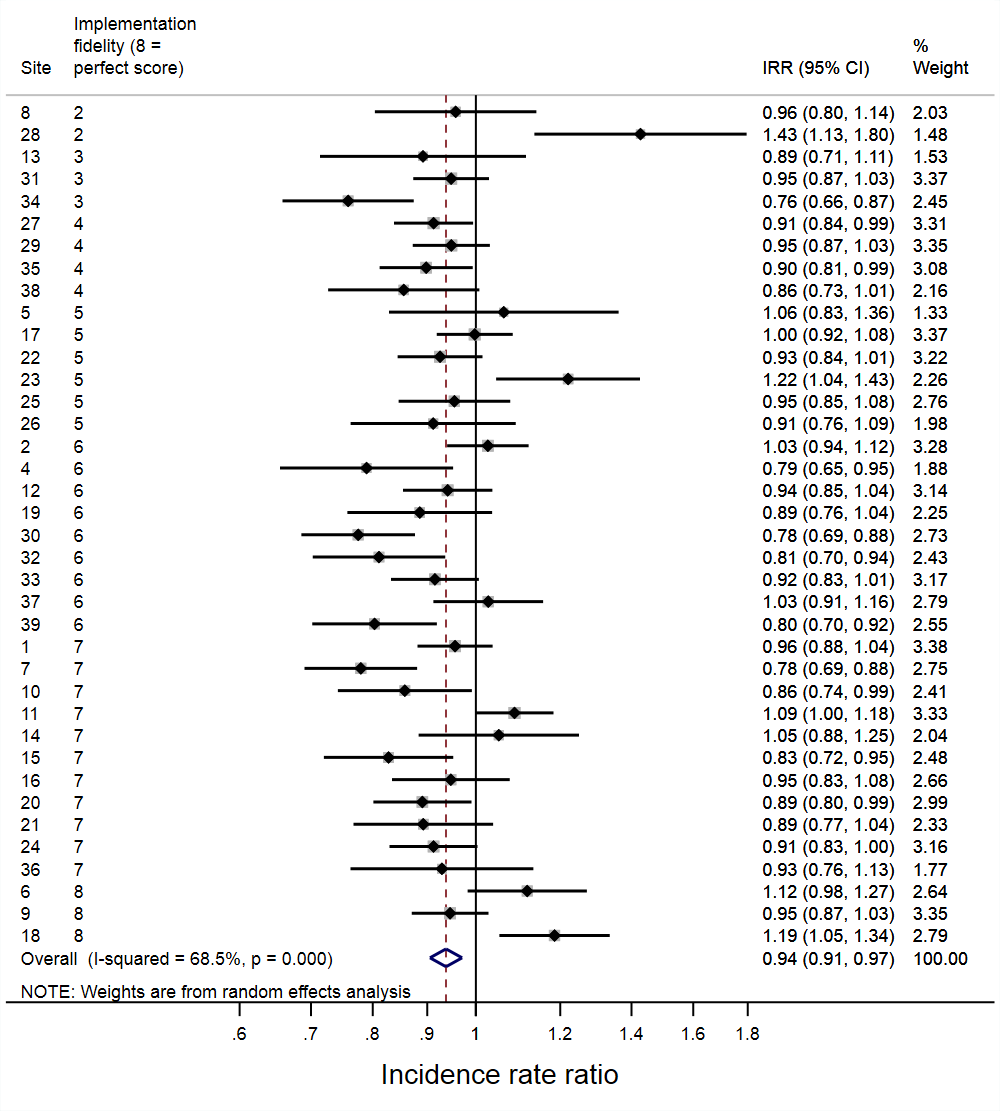 | **B** | **Broad spectrum DDD/admission**  **Change in trend post vs pre-implementation**  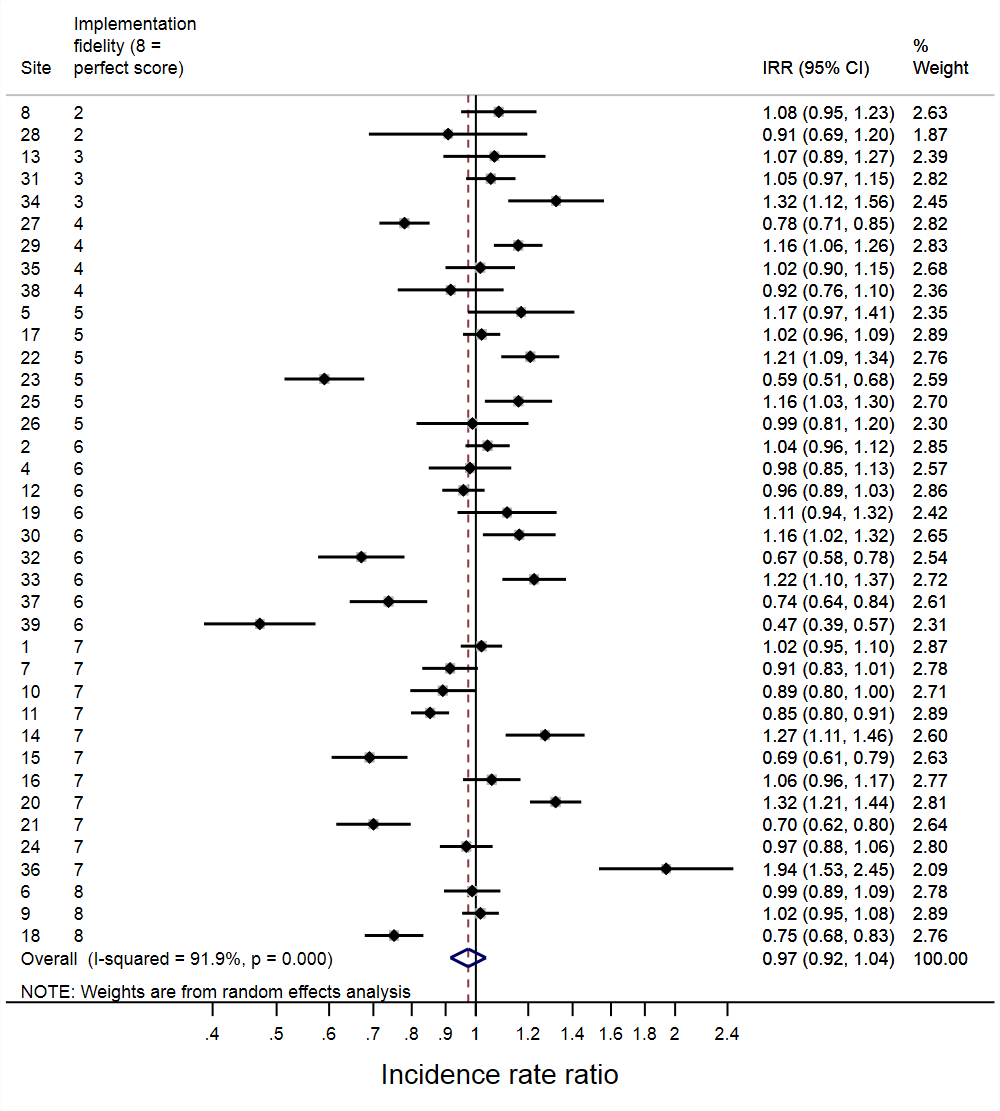 |
| --- | --- | --- | --- |
| **C** | **Narrow spectrum DDD/admission**  **Immediate implementation effect**  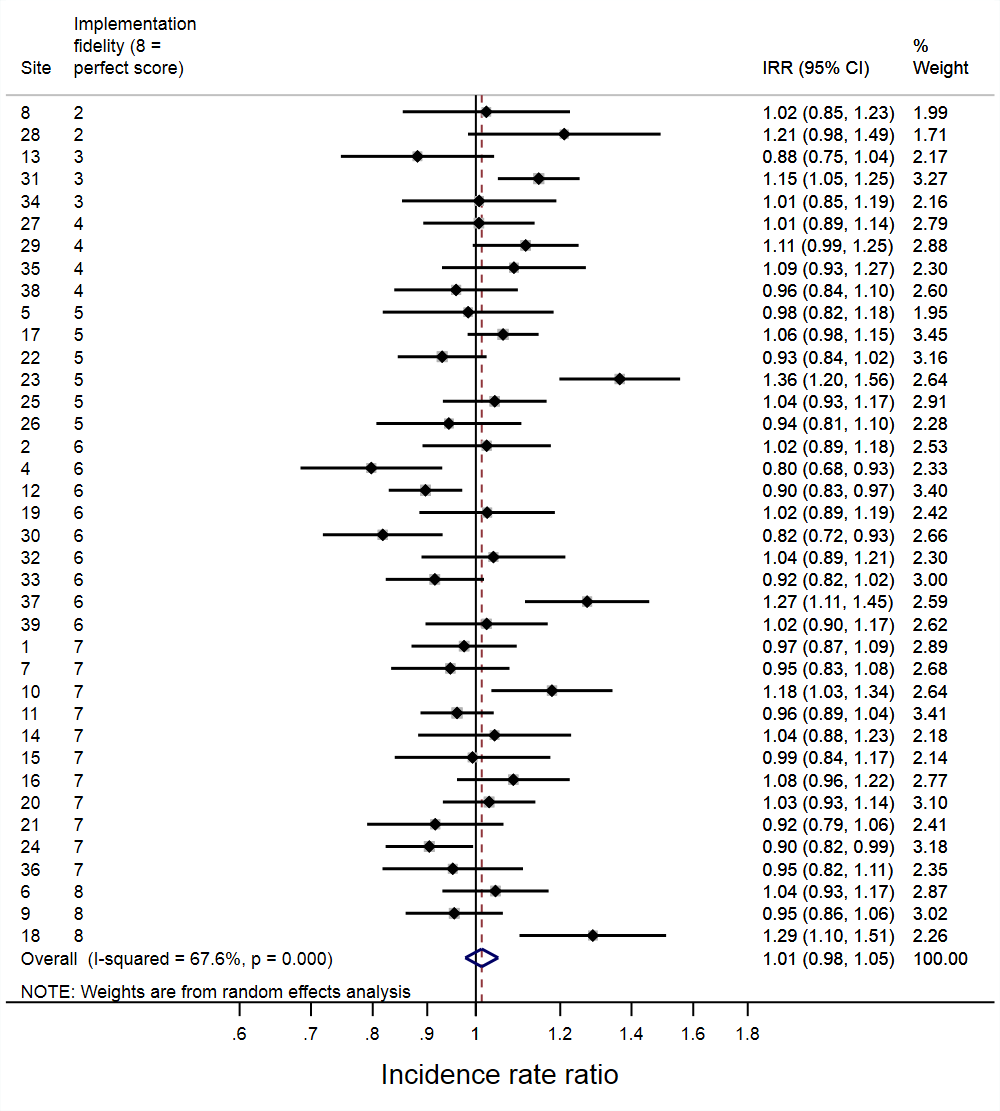 | **D** | **Narrow spectrum DDD/admission**  **Change in trend post vs pre-implementation**  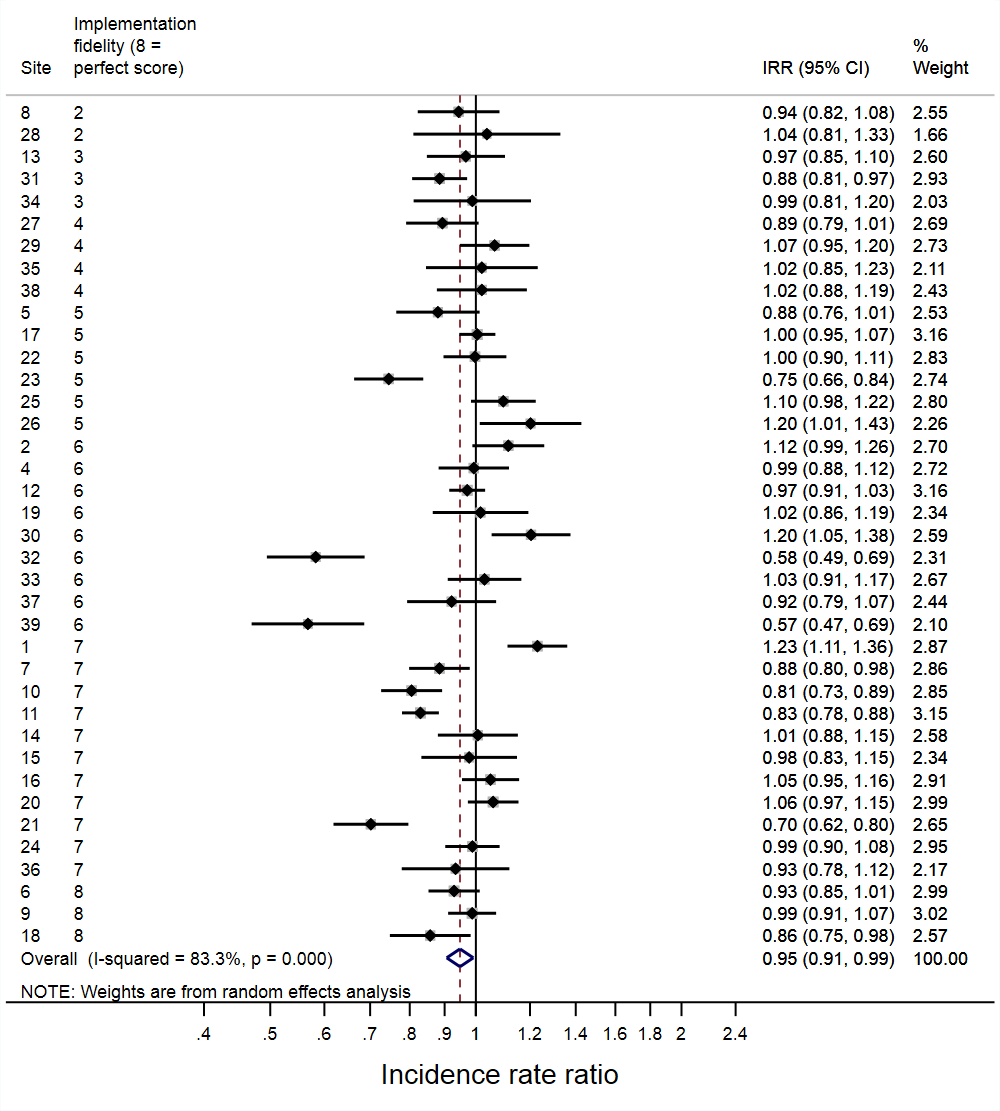 |

Supplementary Figure S10: Antibiotic DDD/admission classified by WHO’s 2019 AWaRe categories (secondary outcome); immediate impact of implementation Access antibiotics (A), change in trend post vs pre-implementation Access antibiotics (B), immediate impact of implementation Access or Watch antibiotics (C), change in trend post vs pre-implementation Access or Watch antibiotics (D), immediate impact of implementation Watch antibiotics (E), change in trend post vs pre-implementation Watch antibiotics (F), immediate impact of implementation Reserve antibiotics (G), change in trend post vs pre-implementation Reserve antibiotics (H)

| **A** | **Access antibiotic DDD/admission**  **Immediate implementation effect**  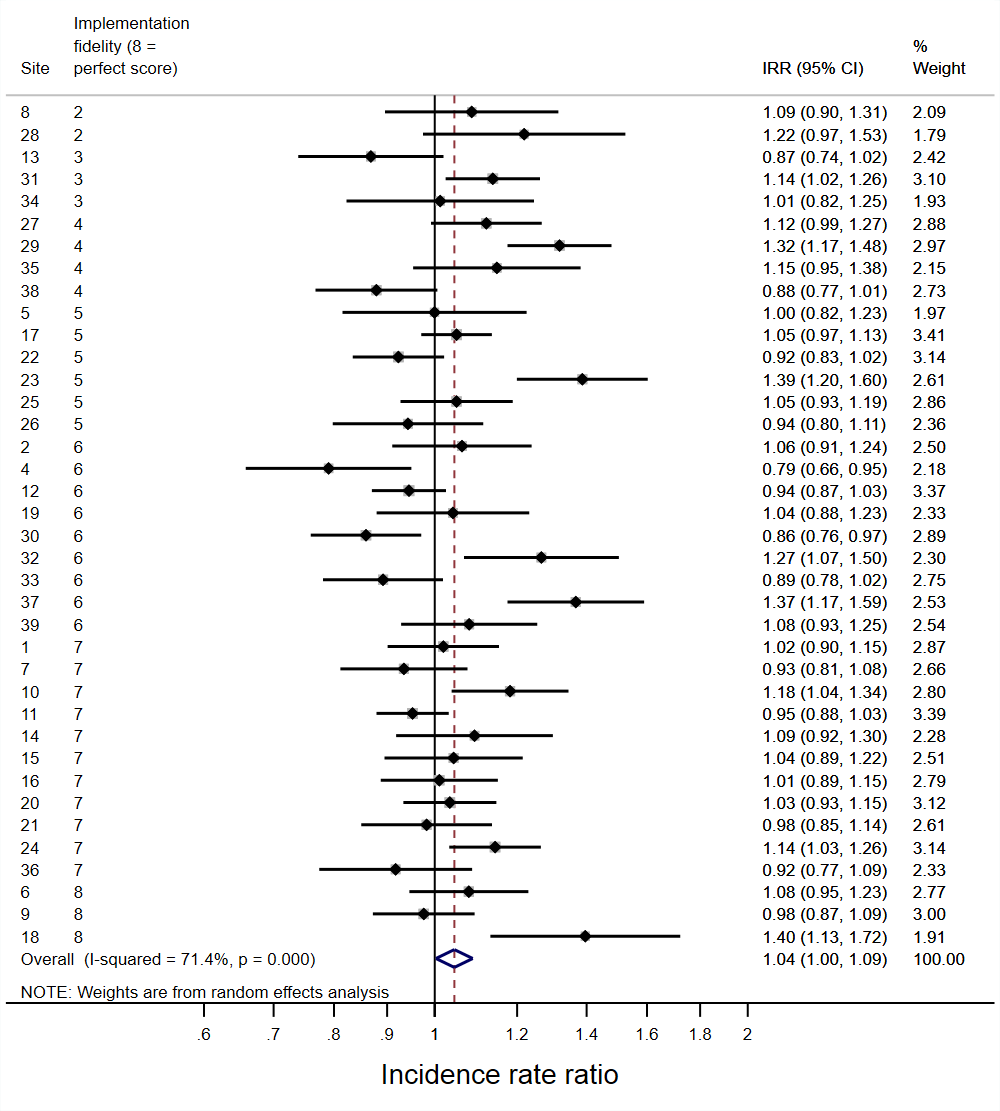 | **B** | **Access antibiotic DDD/admission**  **Change in trend post vs pre-implementation**  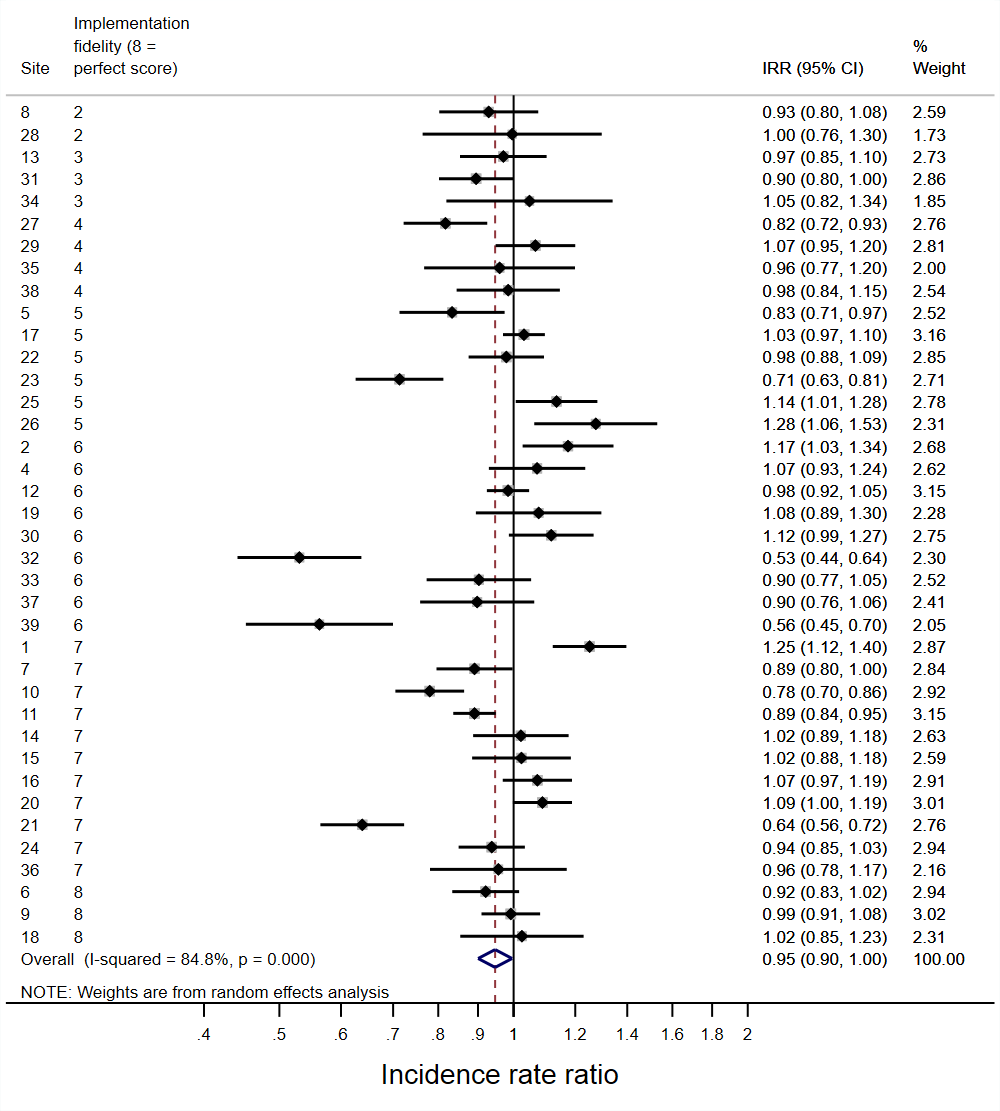 |
| --- | --- | --- | --- |
| **C** | **Access or Watch antibiotic DDD/admission***  **Immediate implementation effect**  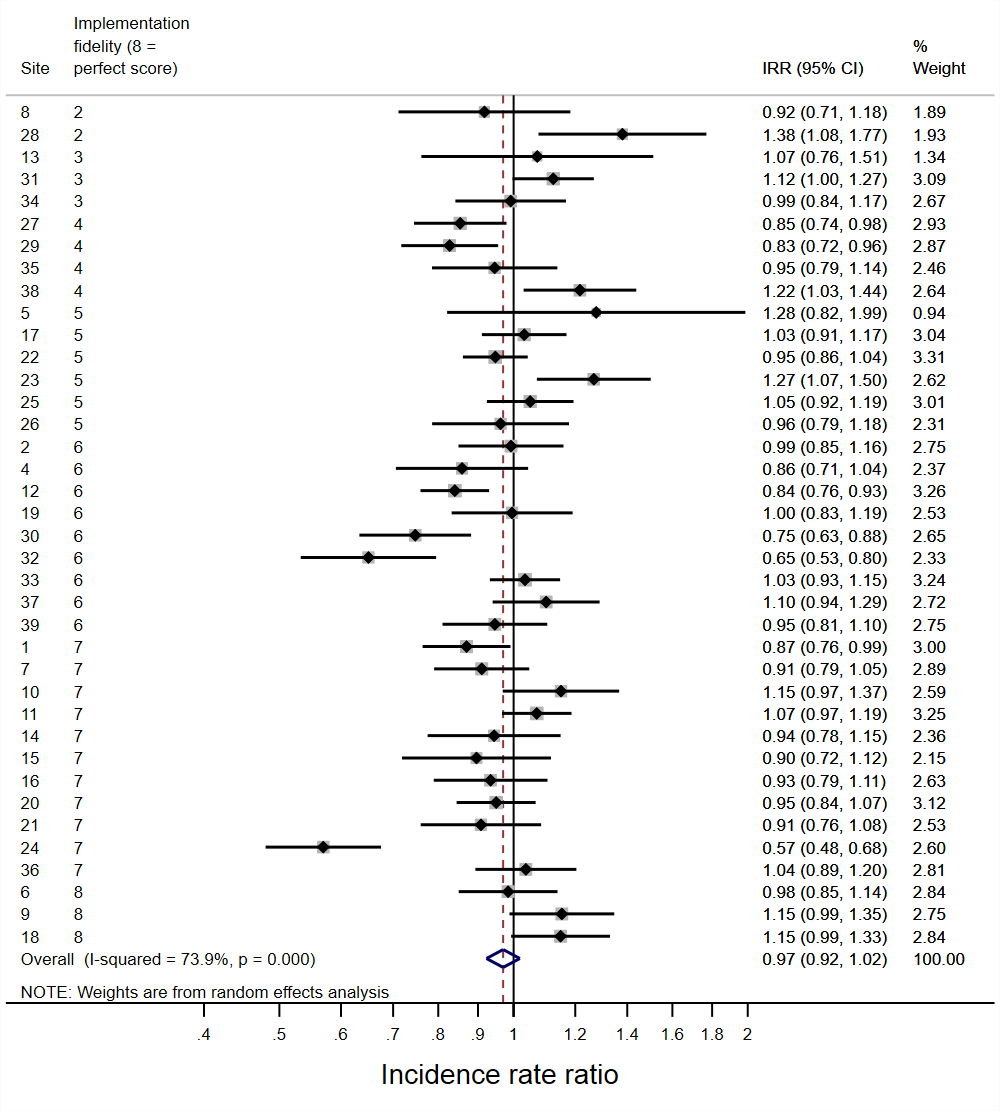 | **D** | **Access or Watch antibiotic DDD/admission***  **Change in trend post vs pre-implementation**  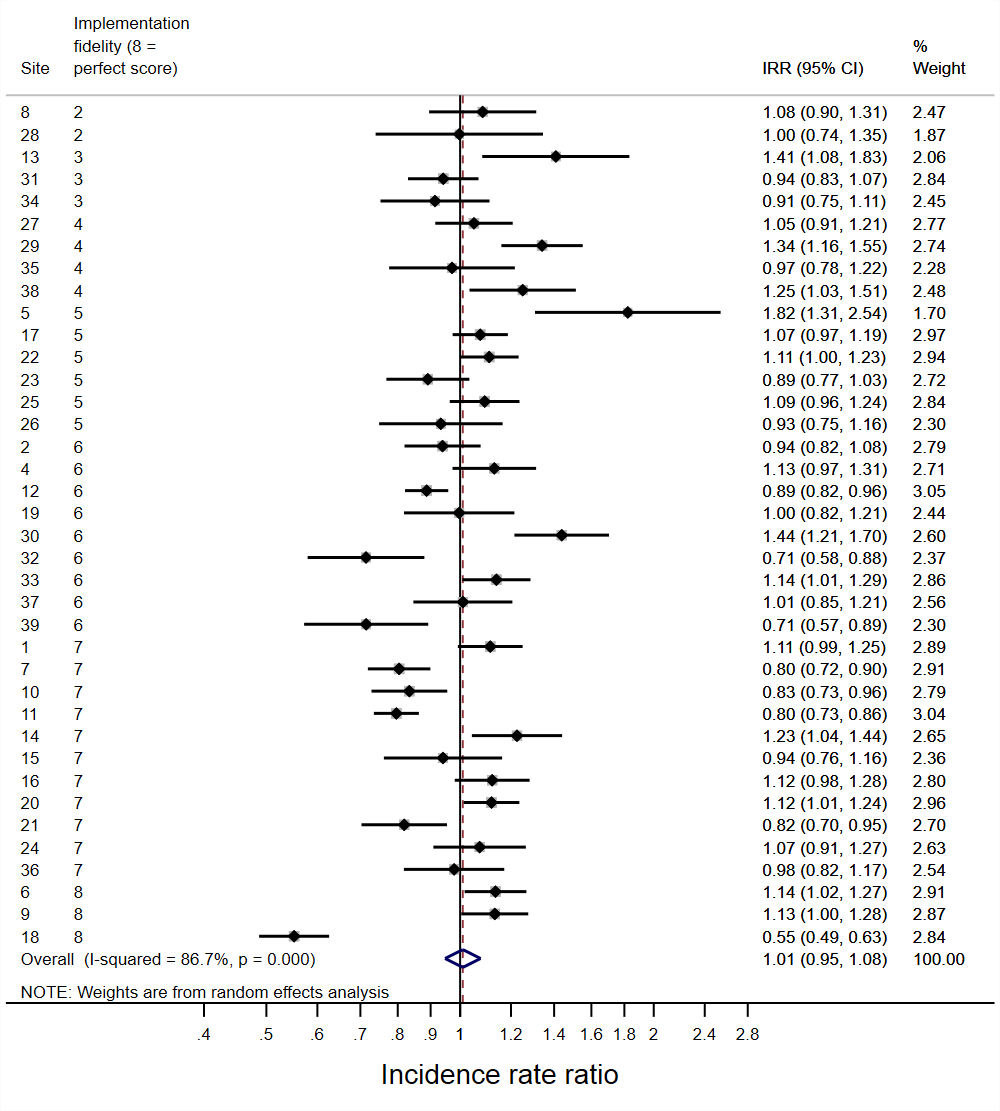 |

* Classified as either Access or Watch depending on indication

| **E** | **Watch antibiotic DDD/admission**  **Immediate implementation effect**  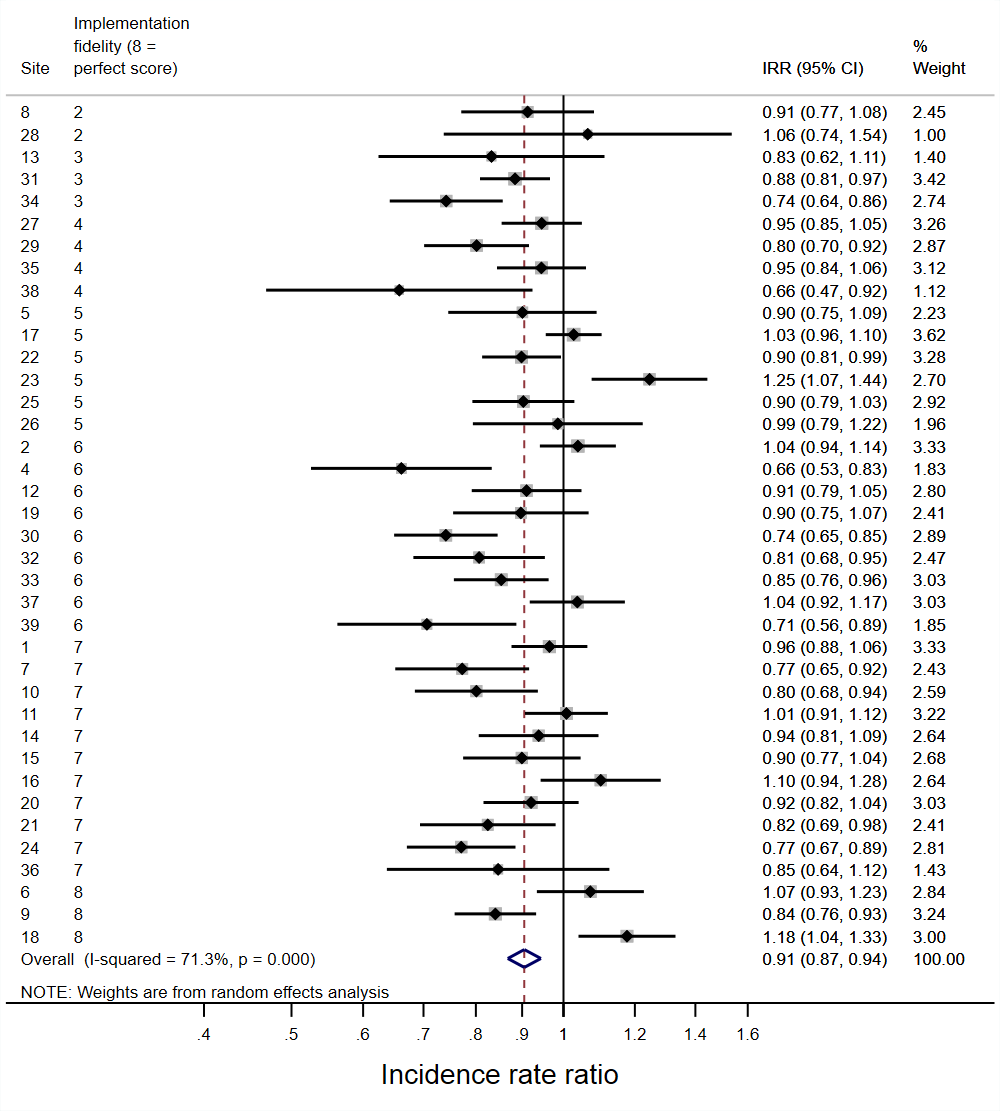 | **F** | **Watch antibiotic DDD/admission**  **Change in trend post vs pre-implementation**  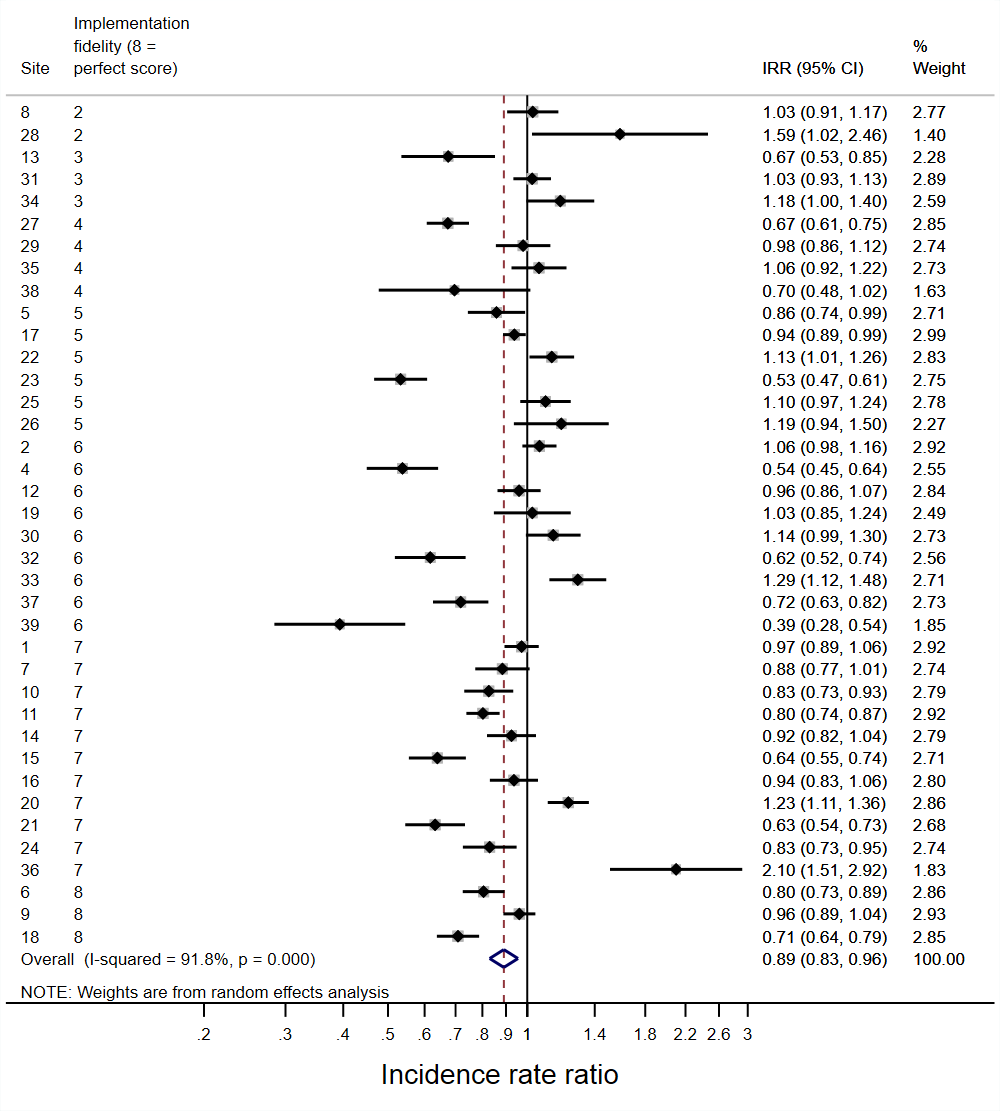 |
| --- | --- | --- | --- |
| **G** | **Reserve antibiotic DDD/admission**  **Immediate implementation effect** 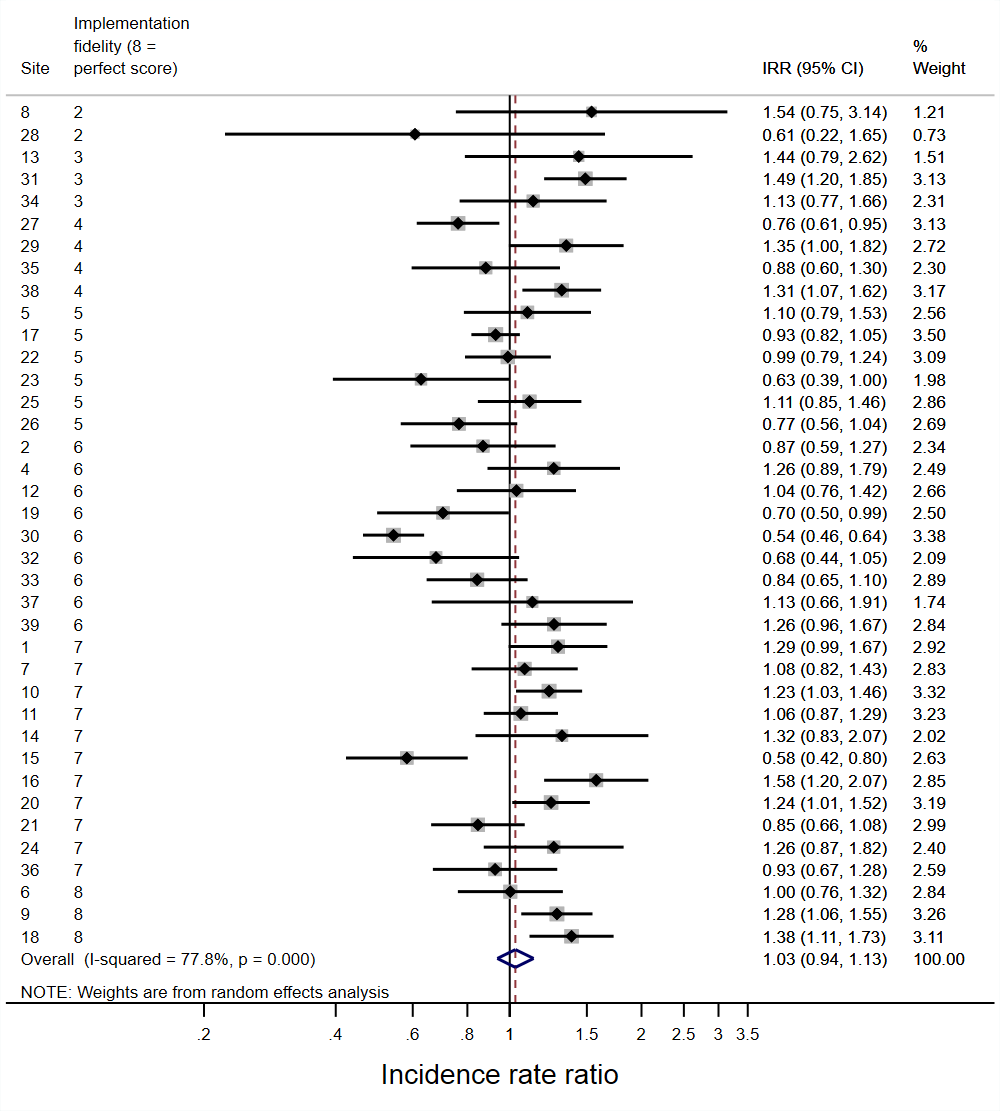 | **H** | **Reserve antibiotic DDD/admission**  **Change in trend post vs pre-implementation**  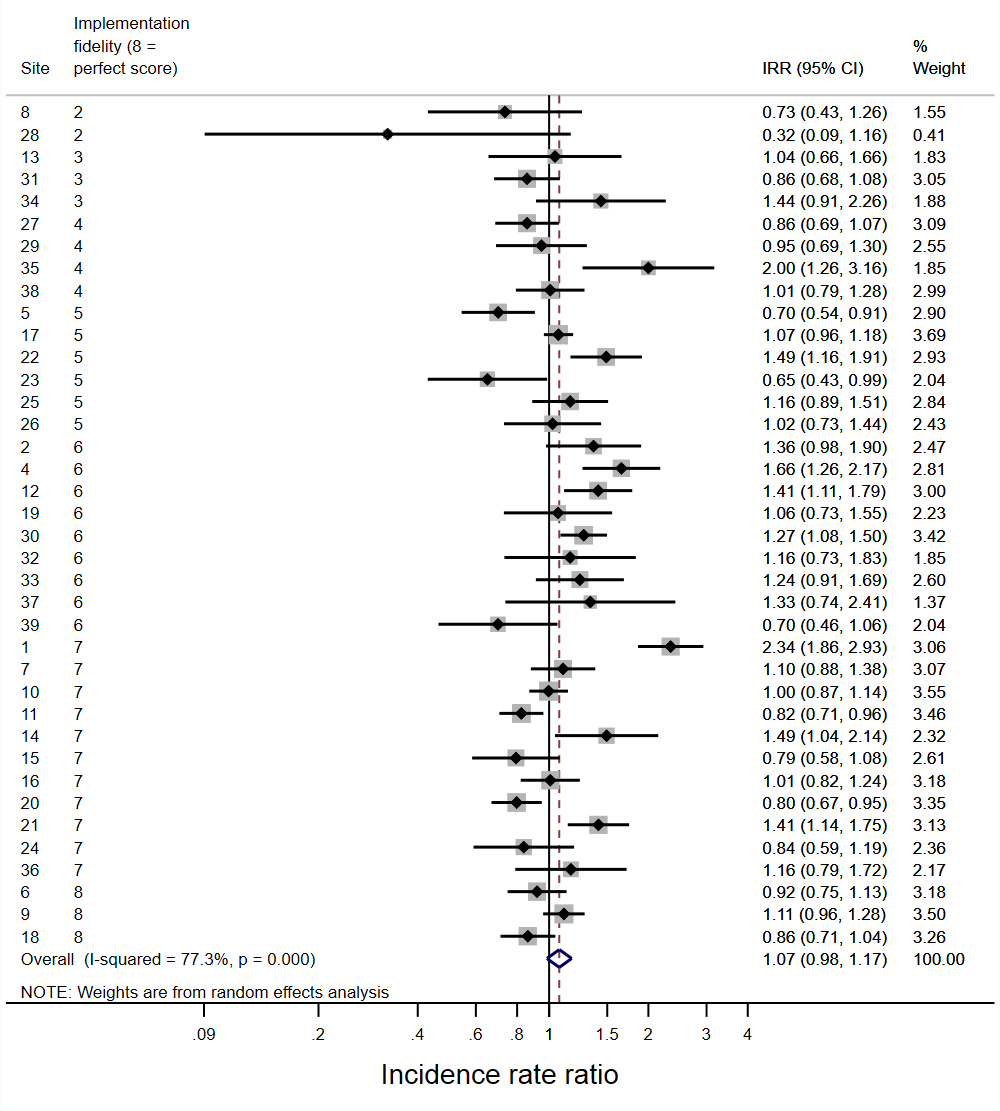 |

#### Supplementary Figure S11: Antibiotic DDD/admission classified by route of administration (secondary outcome); **immediate impact of implementation parenteral antibiotics (A), change in trend post vs pre-implementation parenteral antibiotics (B), immediate impact of implementation oral antibiotics (C), change in trend post vs pre-implementation oral antibiotics (D)**

| **A** | **Parenteral DDD/admission**  **Immediate implementation effect**  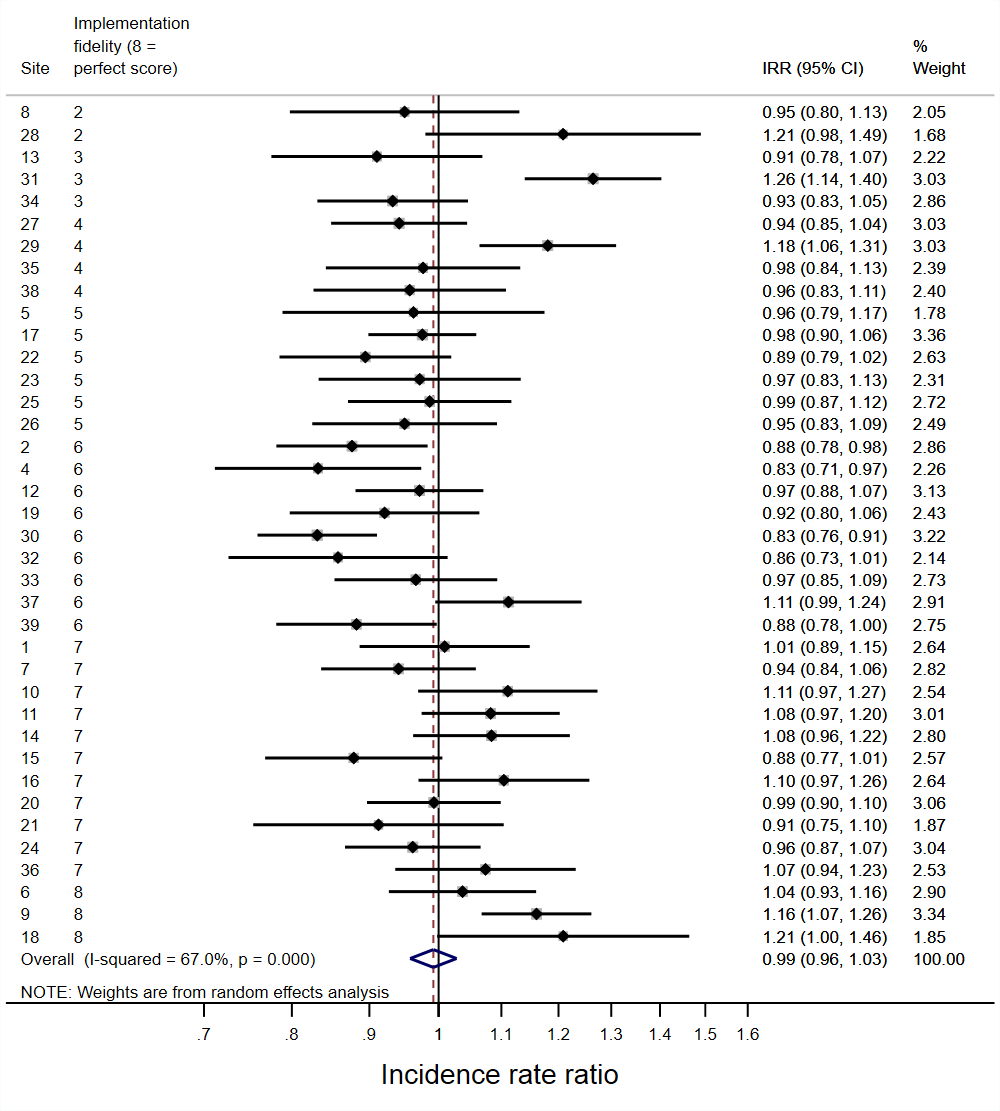 | **B** | **Parenteral DDD/admission**  **Change in trend post vs pre-implementation**  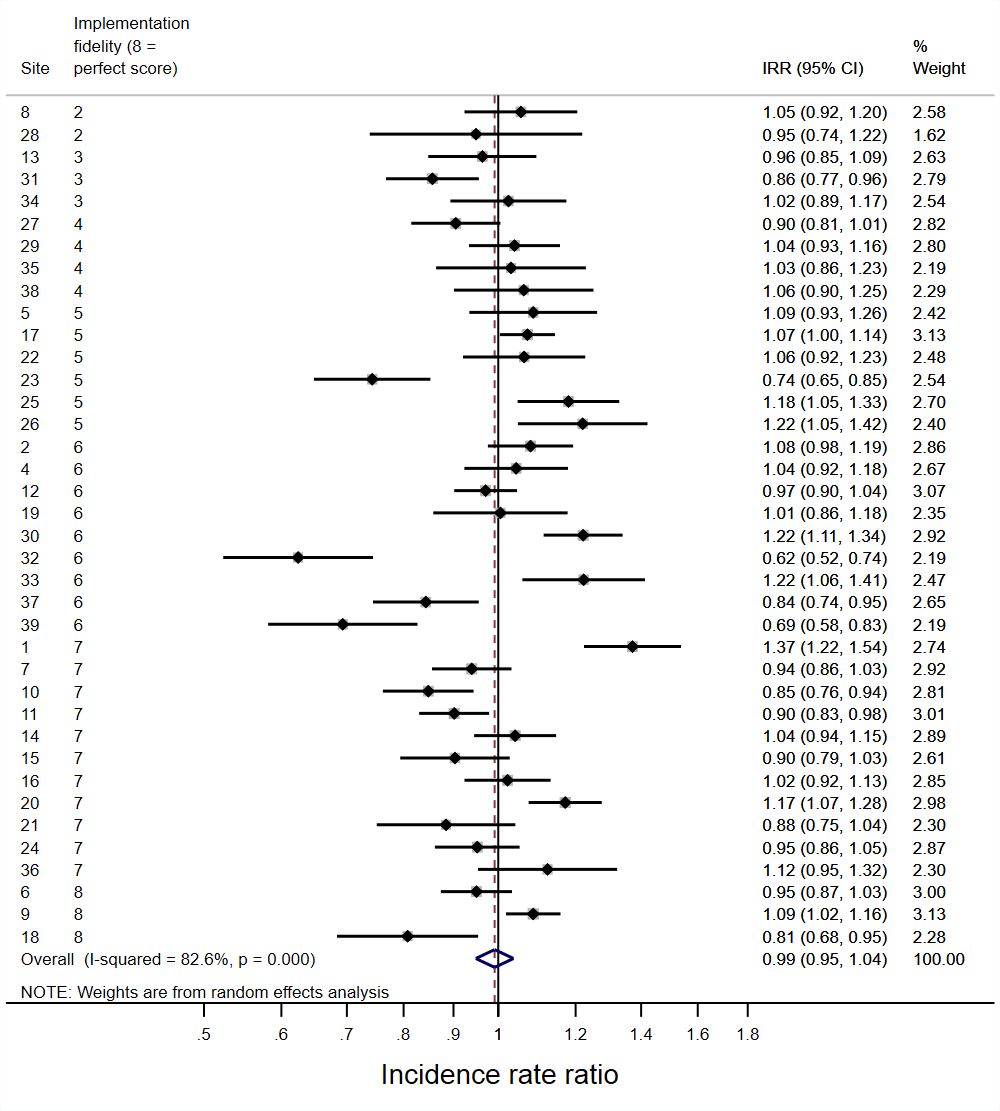 |
| --- | --- | --- | --- |
| **C** | **Oral DDD/admission**  **Immediate implementation effect**  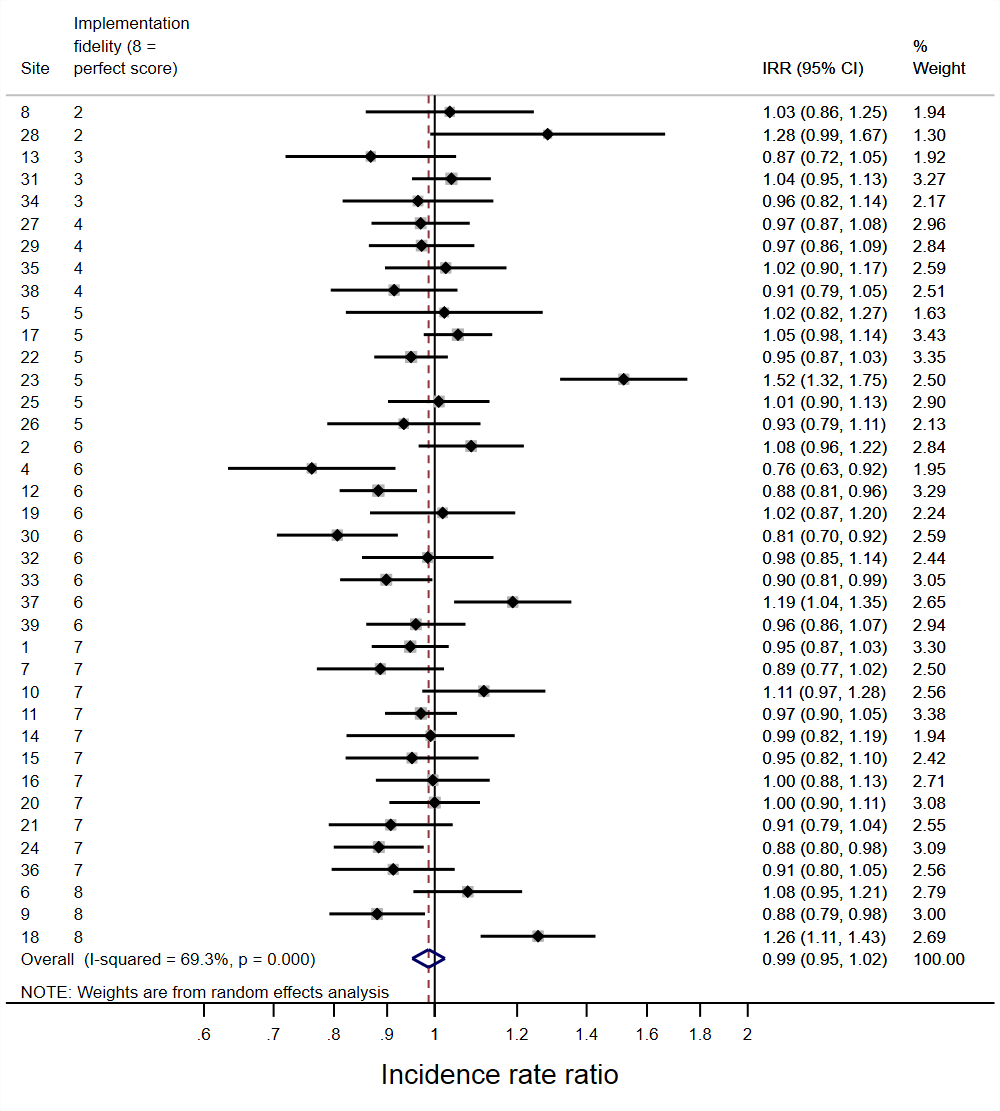 | **D** | **Oral DDD/admission**  **Change in trend post vs pre-implementation**  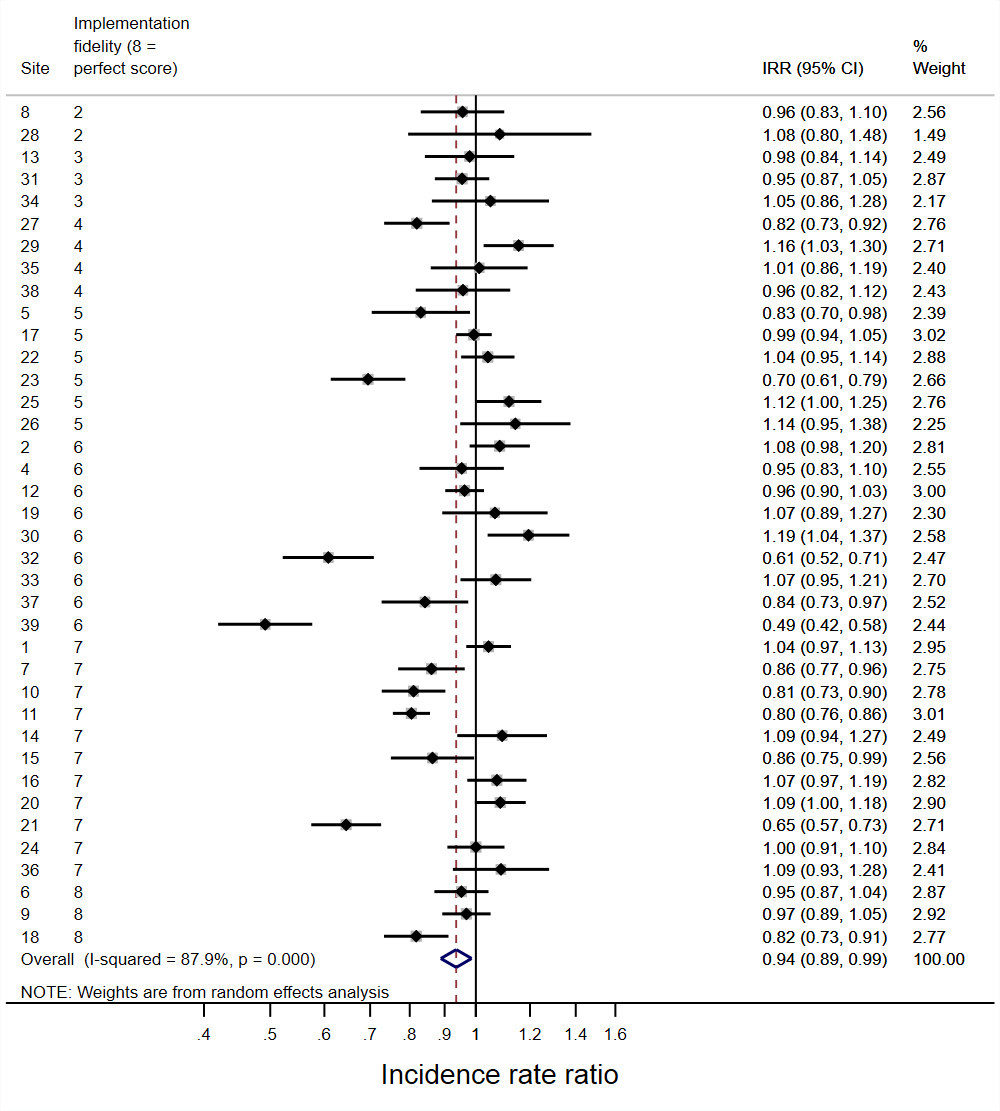 |

#### Supplementary Figure S12: Carbapenem DDD/admission (secondary outcome); **immediate impact of implementation parenteral antibiotics (A), change in trend post vs pre-implementation parenteral antibiotics (B)**

| **A** | **Immediate implementation effect**  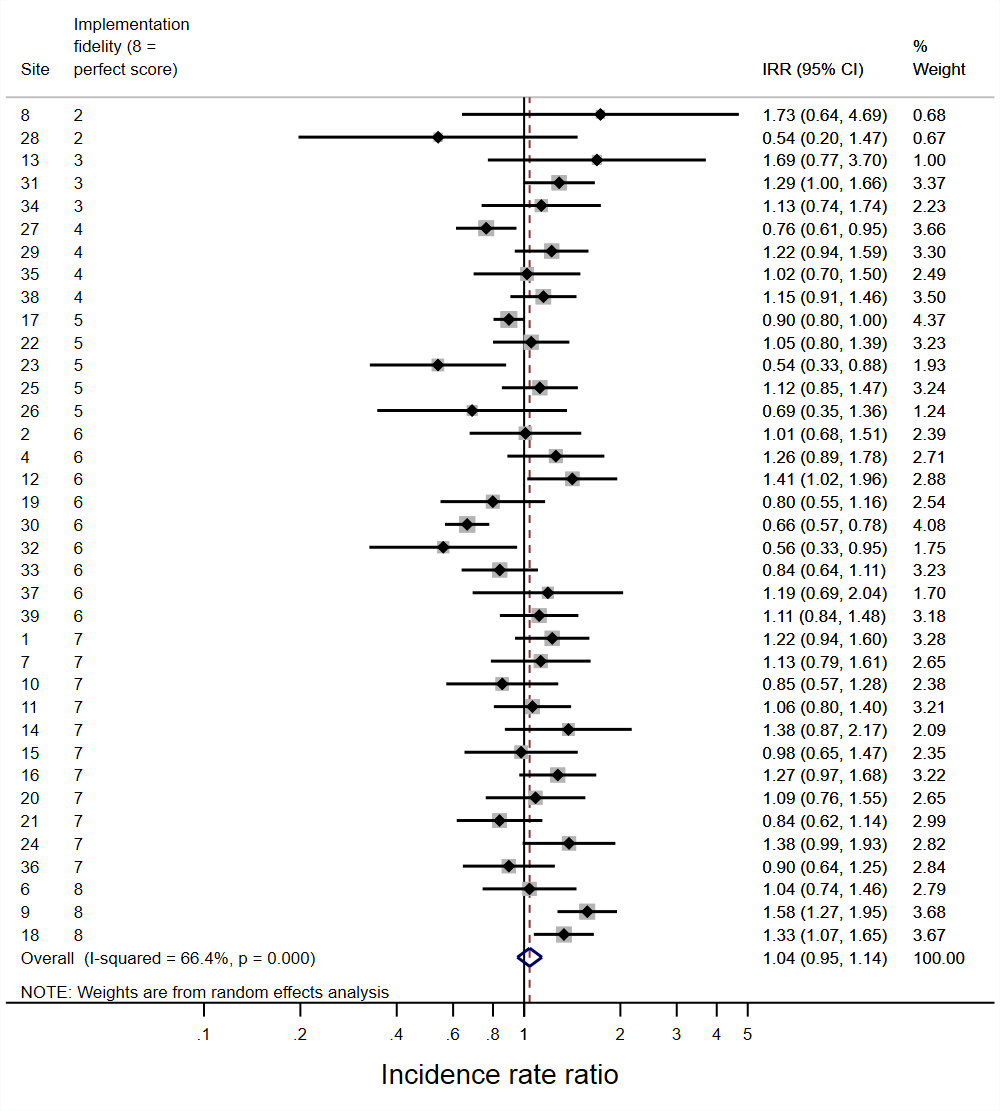 | **B** | **Change in trend post vs pre-implementation**  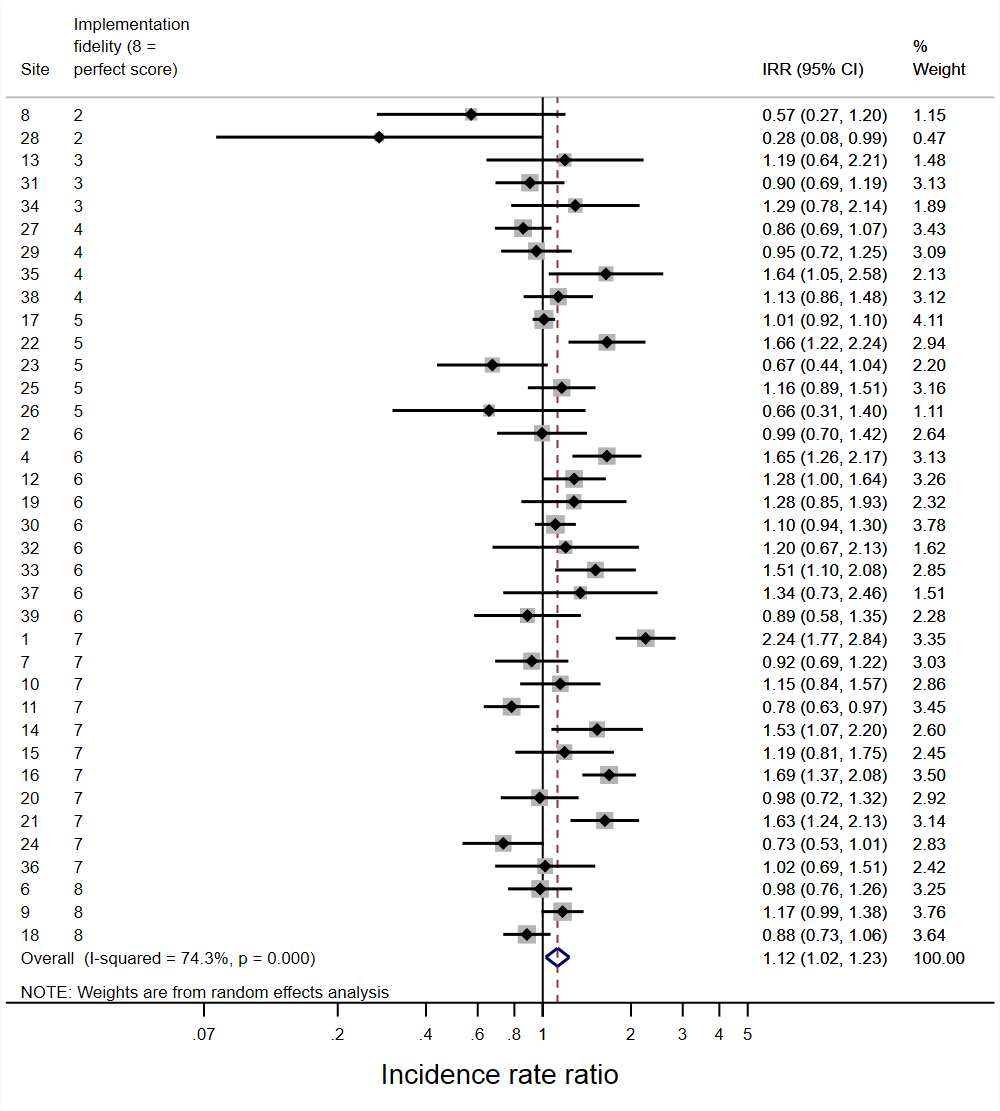 |
| --- | --- | --- | --- |

#### Supplementary Figure S13: Intervention effects on 30-day mortality (co-primary outcome)

Spearman's rho: -0.282, p=0.082
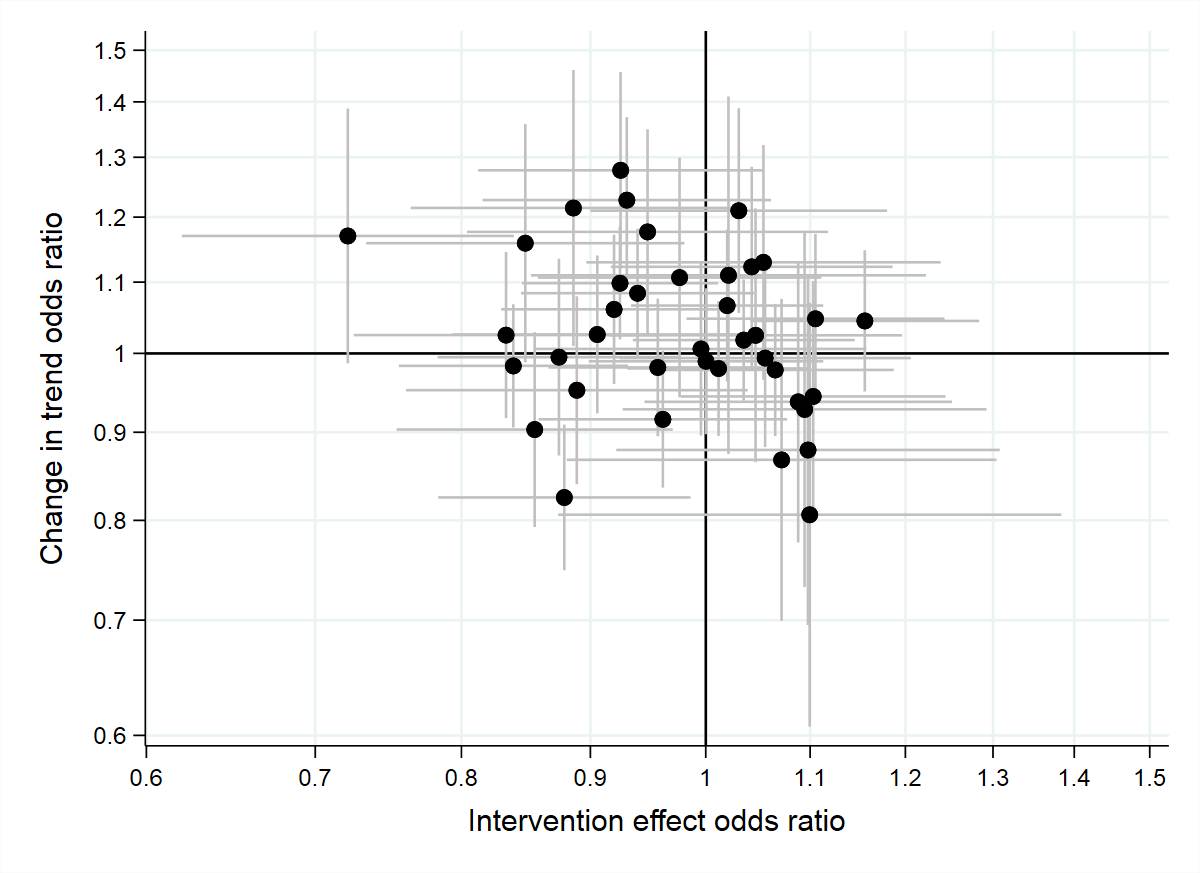

#### Supplementary Figure S14: 30-day mortality (co-primary endpoint); change over time pre- (A) and post-implementation (B)

| **A** | **Pre-implementation trend (per year)**  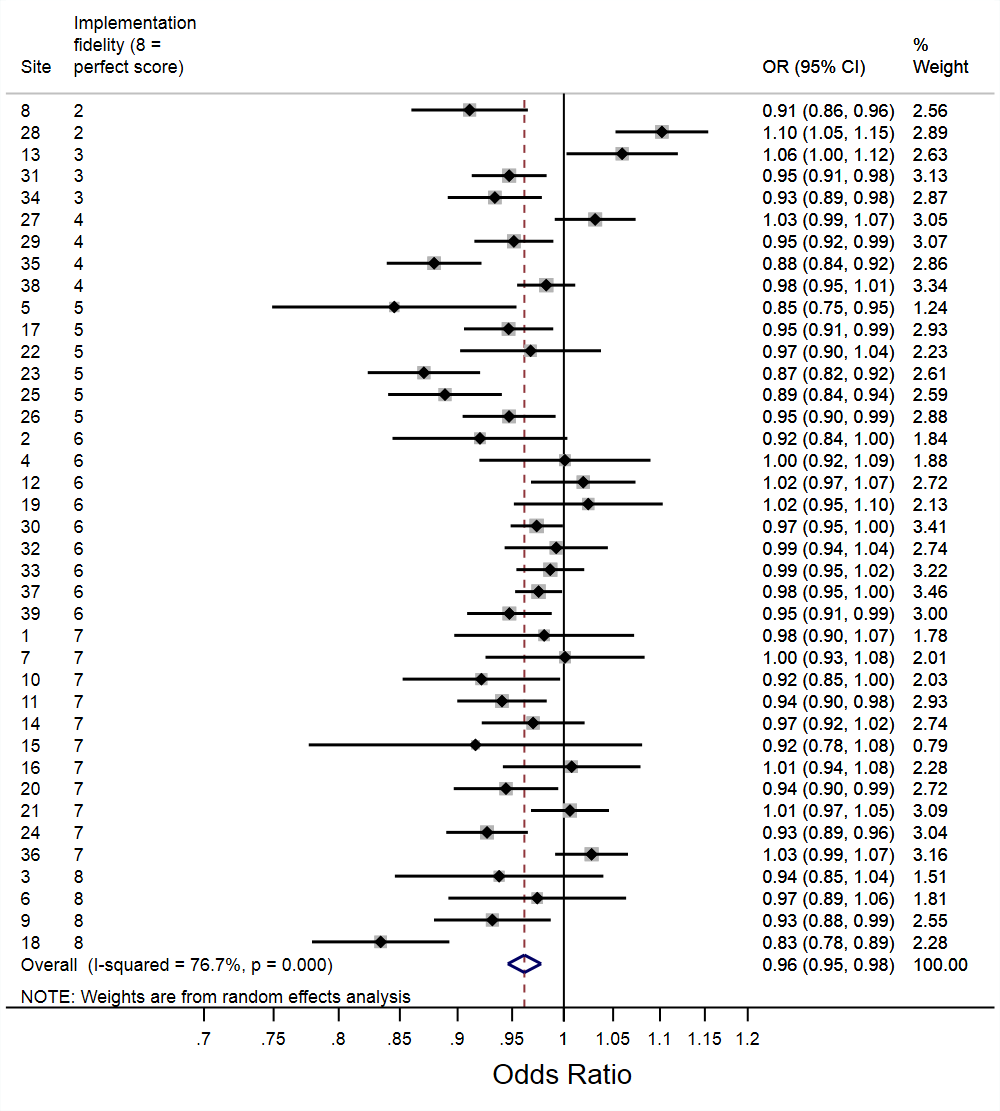 | **B** | **Post-implementation trend (per year)**  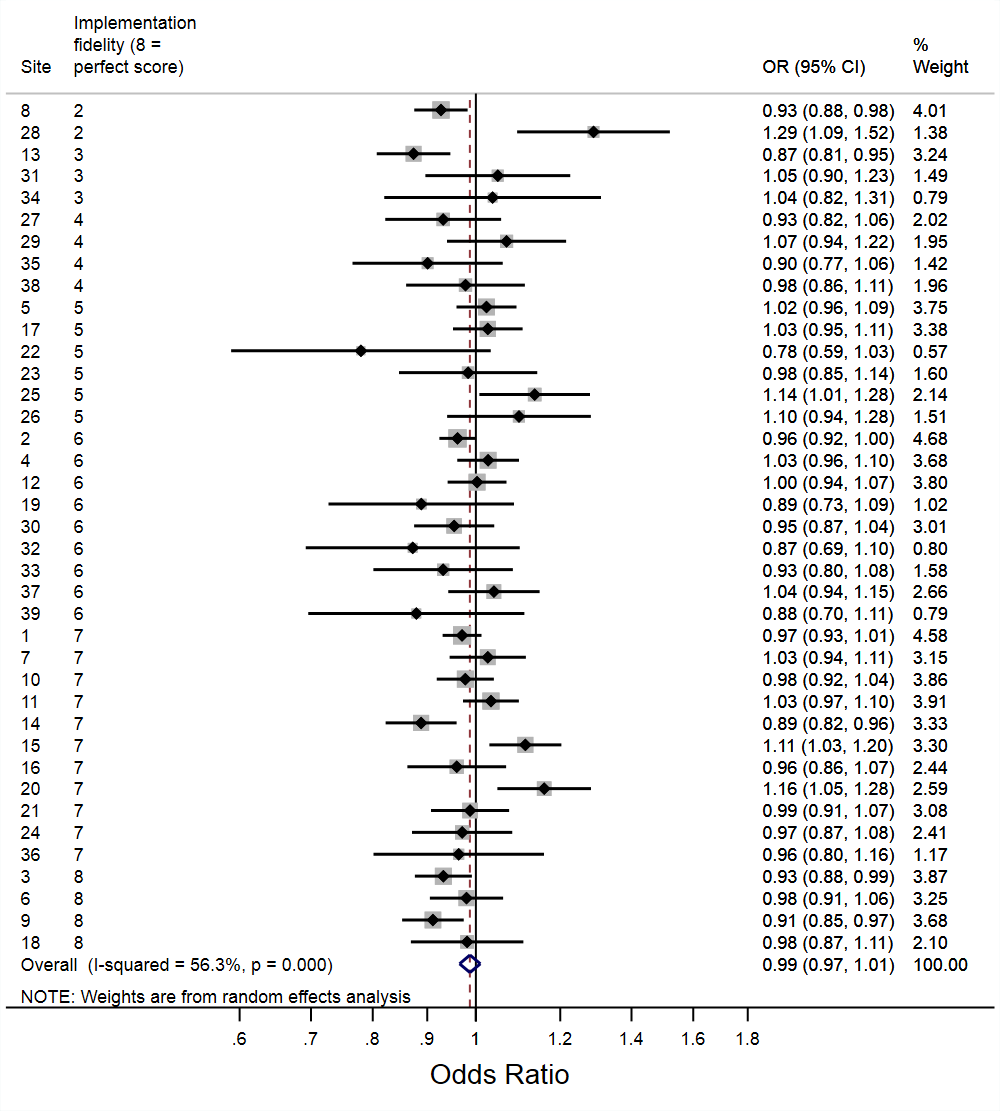 |
| --- | --- | --- | --- |

#### Supplementary Figure S15: 90-day mortality (co-primary endpoint); change over time pre- (A) and post-implementation (B)

| **A** | **Pre-implementation trend (per year)**  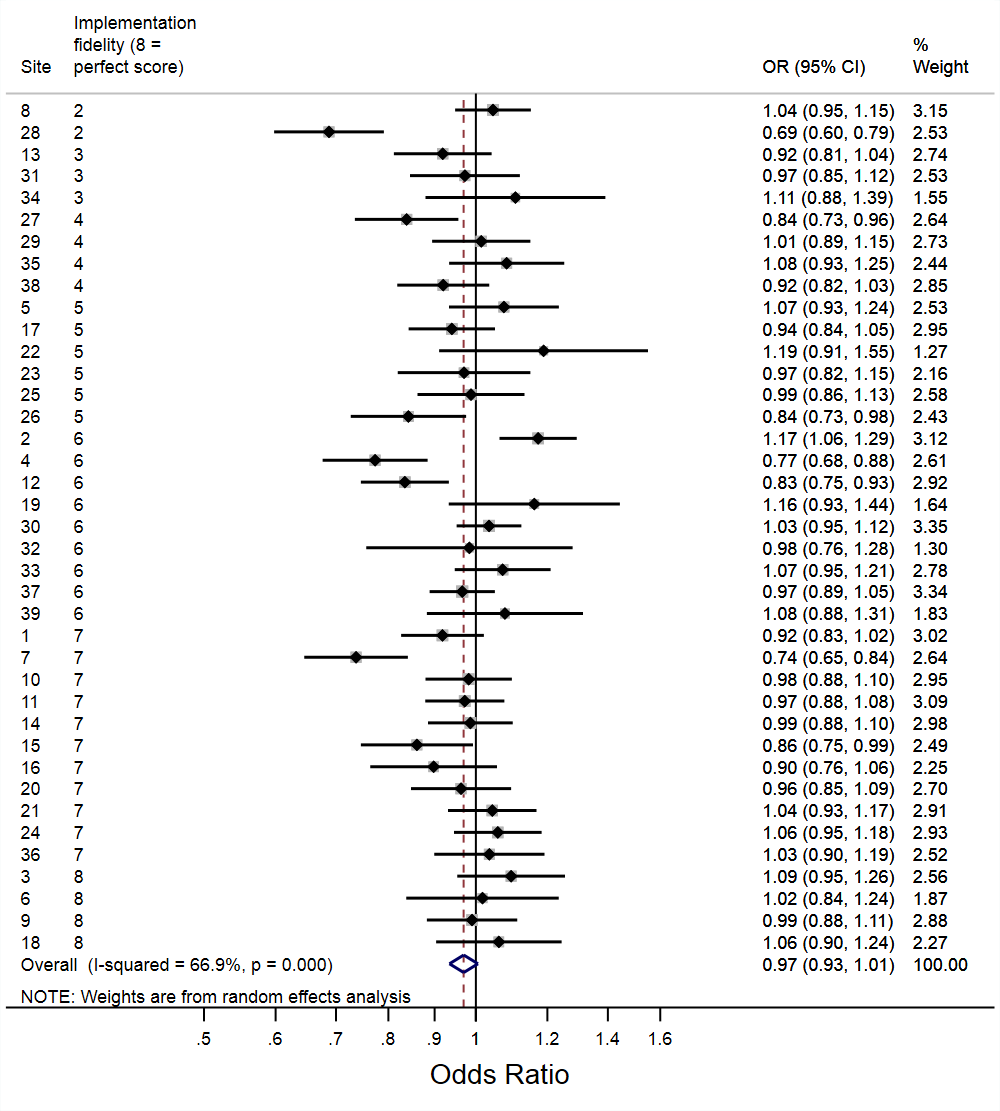 | **B** | **Post-implementation trend (per year)**  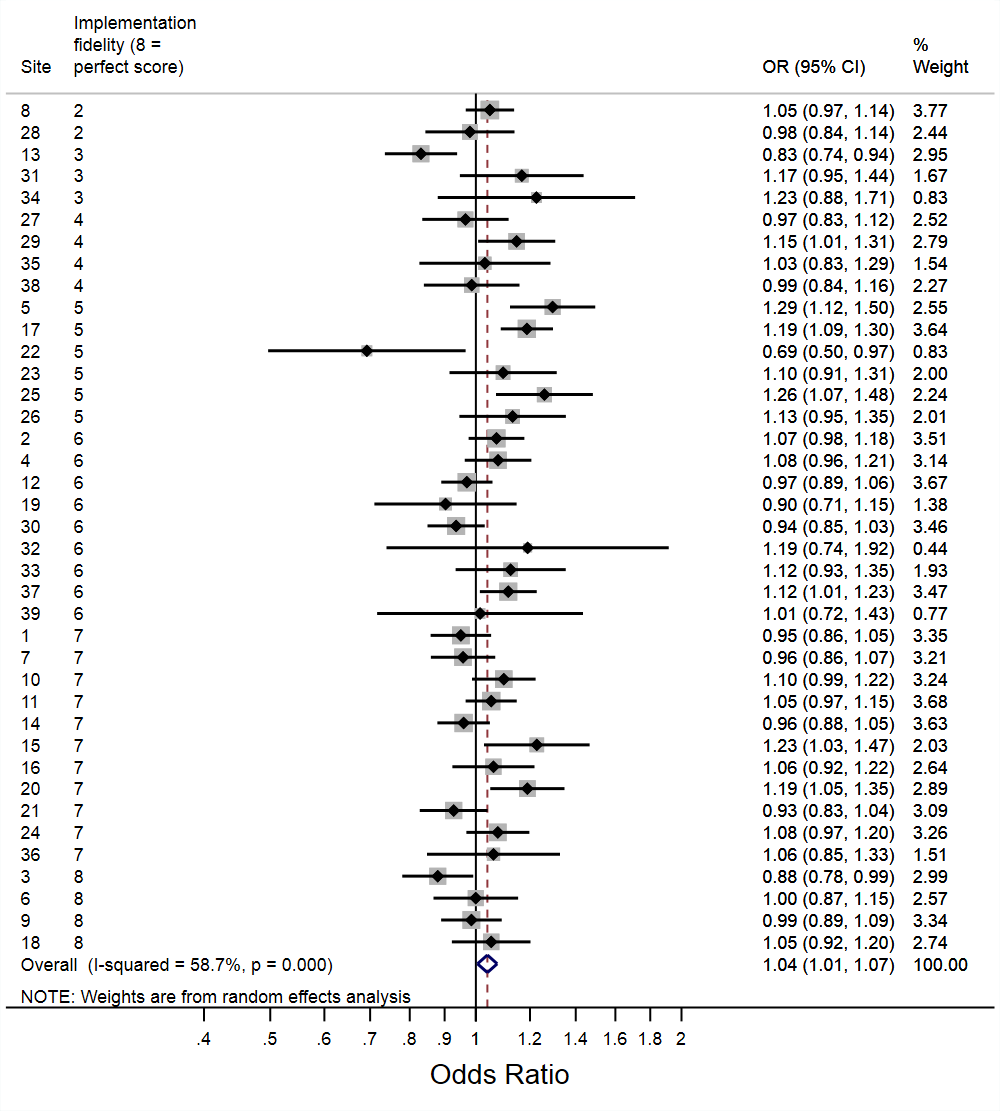 |
| --- | --- | --- | --- |

#### Supplementary Figure S16: ICU admission (secondary outcome); immediate impact of implementation (A), infection change in trend post- vs pre-implementation (B)

| **A** | **Immediate implementation effect**  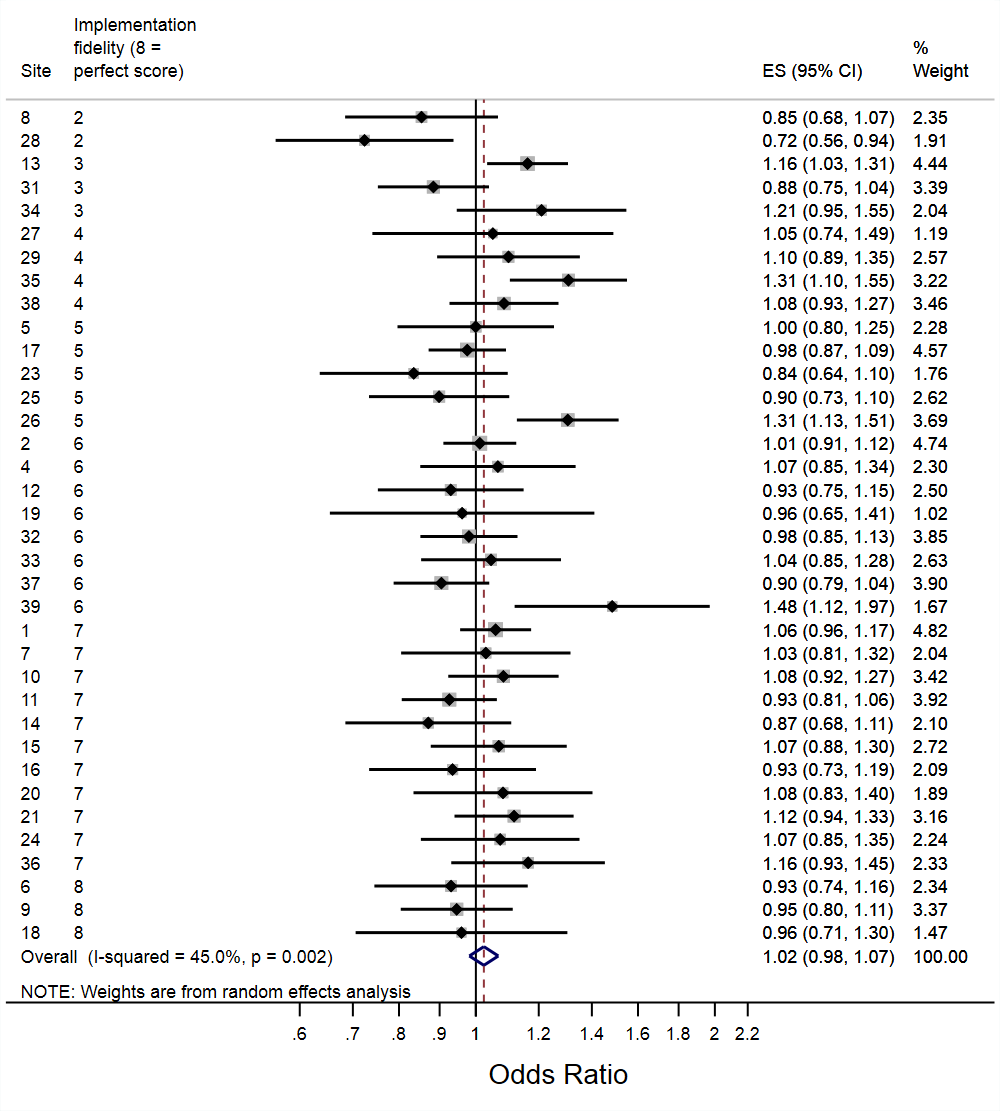 | **B** | **Change in trend post vs pre-implementation**  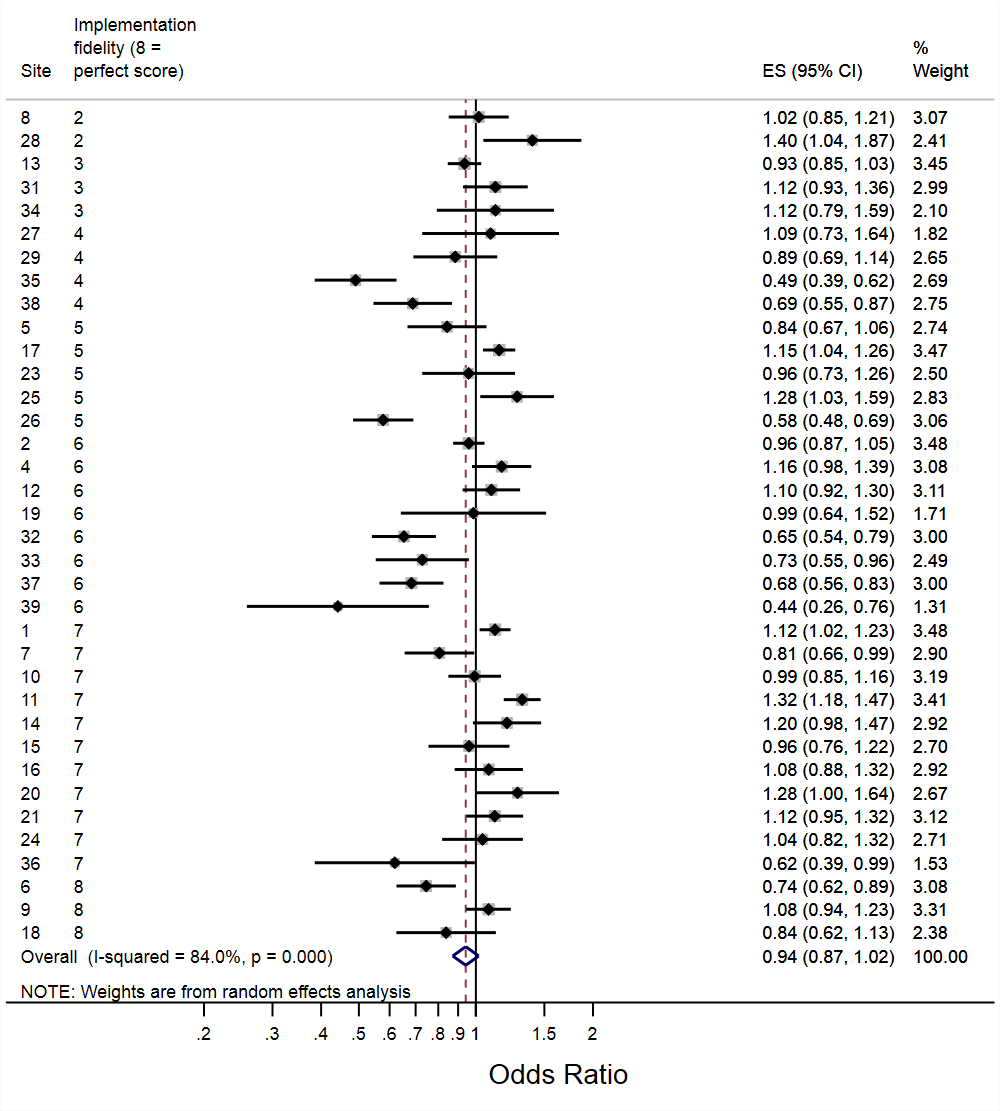 |
| --- | --- | --- | --- |

#### Supplementary Figure S17: Length of hospital stay (days); **immediate impact of implementation (A), change in trend post vs pre-implementation (B)**

| **A** | **Immediate implementation effect**  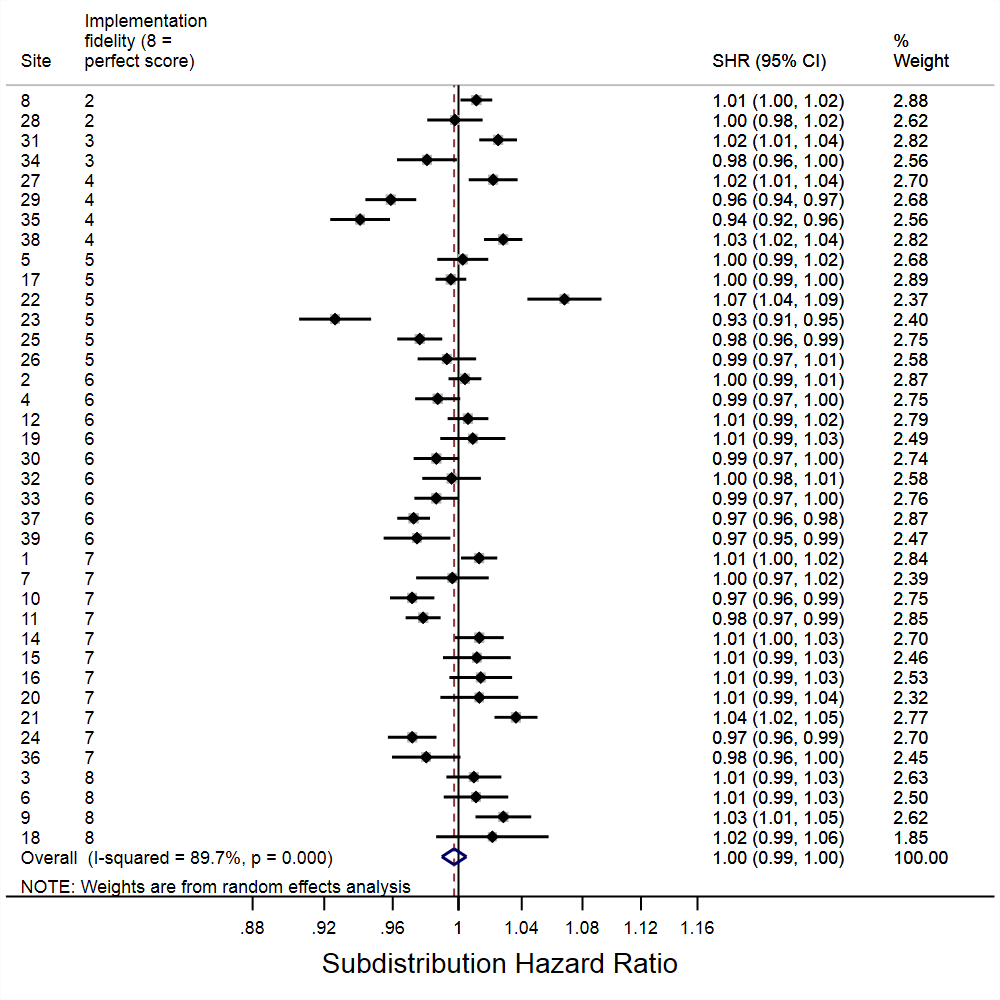 | **B** | **Change in trend post vs pre-implementation**  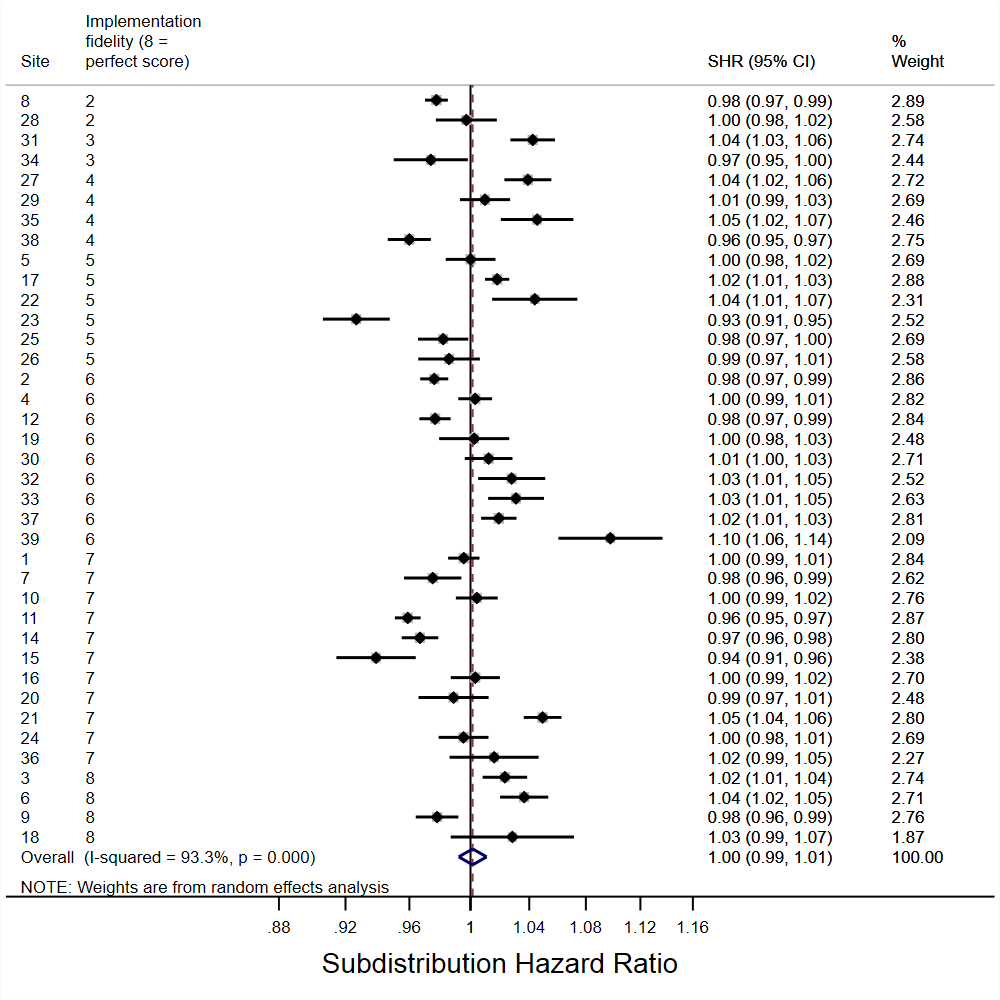 |
| --- | --- | --- | --- |

#### Supplementary Figure S18: Emergency readmission to hospital within 30 days of discharge (secondary outcome); immediate impact of implementation (A), infection change in trend post- vs pre-implementation (B)

| **A** | **Immediate implementation effect**  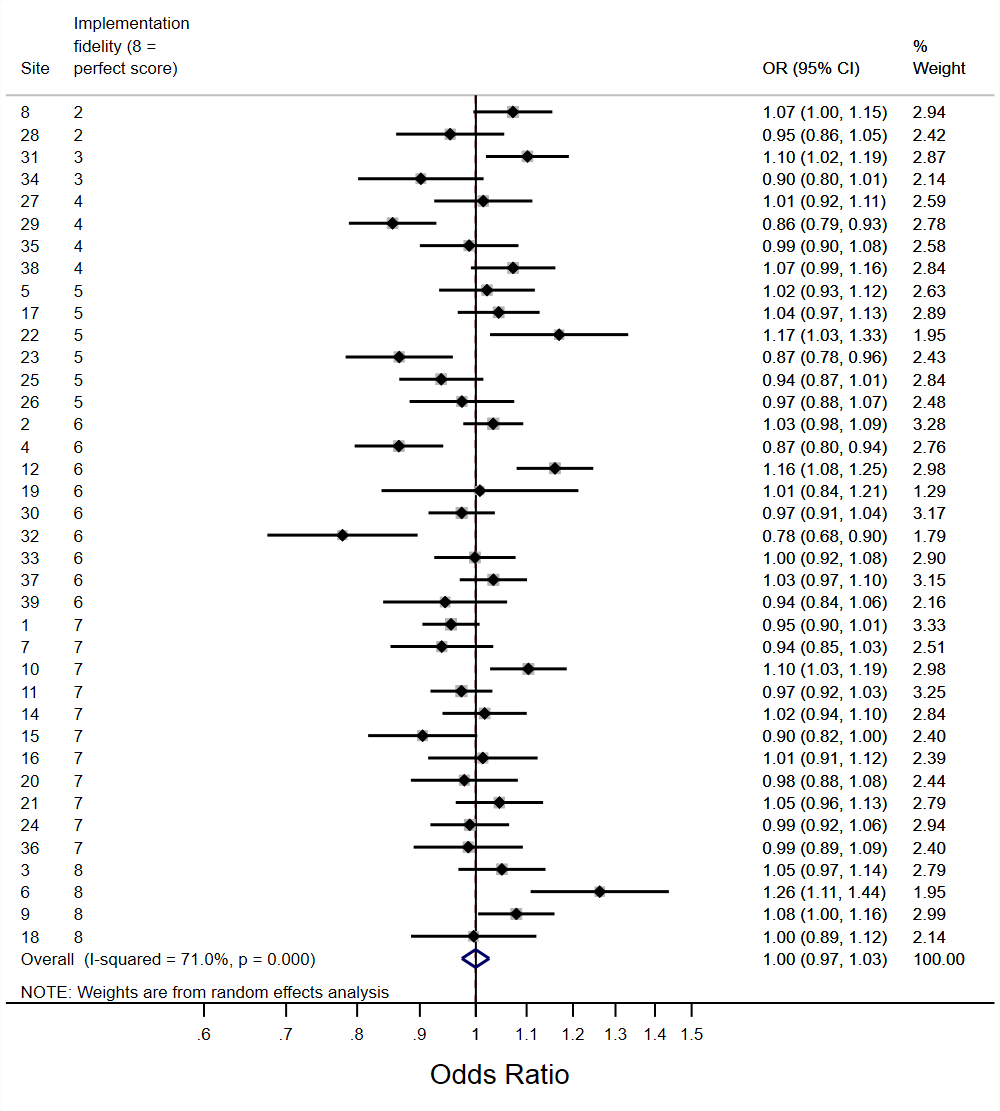 | **B** | **Change in trend post vs pre-implementation**  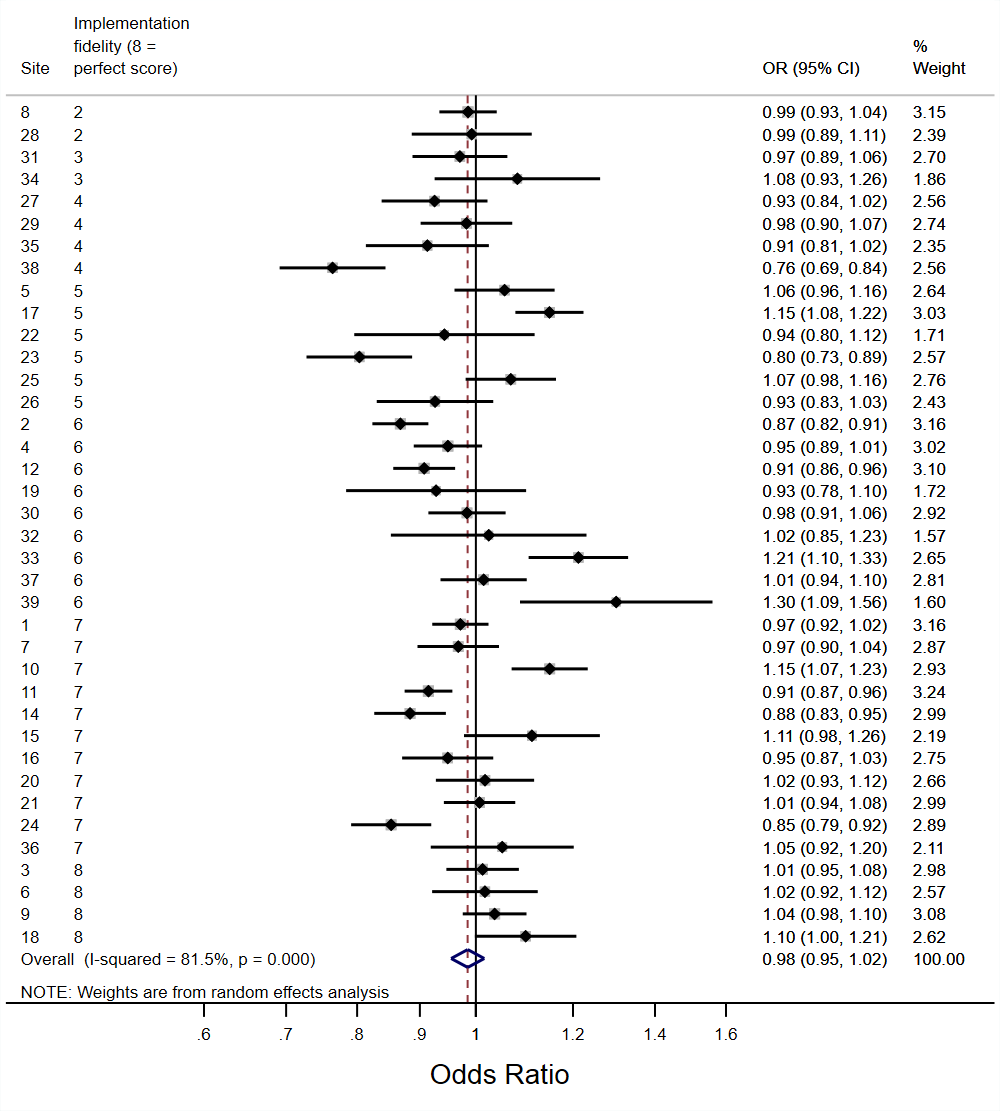 |
| --- | --- | --- | --- |

#### Supplementary Figure S19: *C difficile* (secondary outcome); infection immediate impact of implementation (A), infection change in trend post vs pre implementation (B), colonisation implementation effect (C), colonisation change in trend post vs pre implementation (D)

| **A** | ***C difficile* infection: Immediate implementation effect**  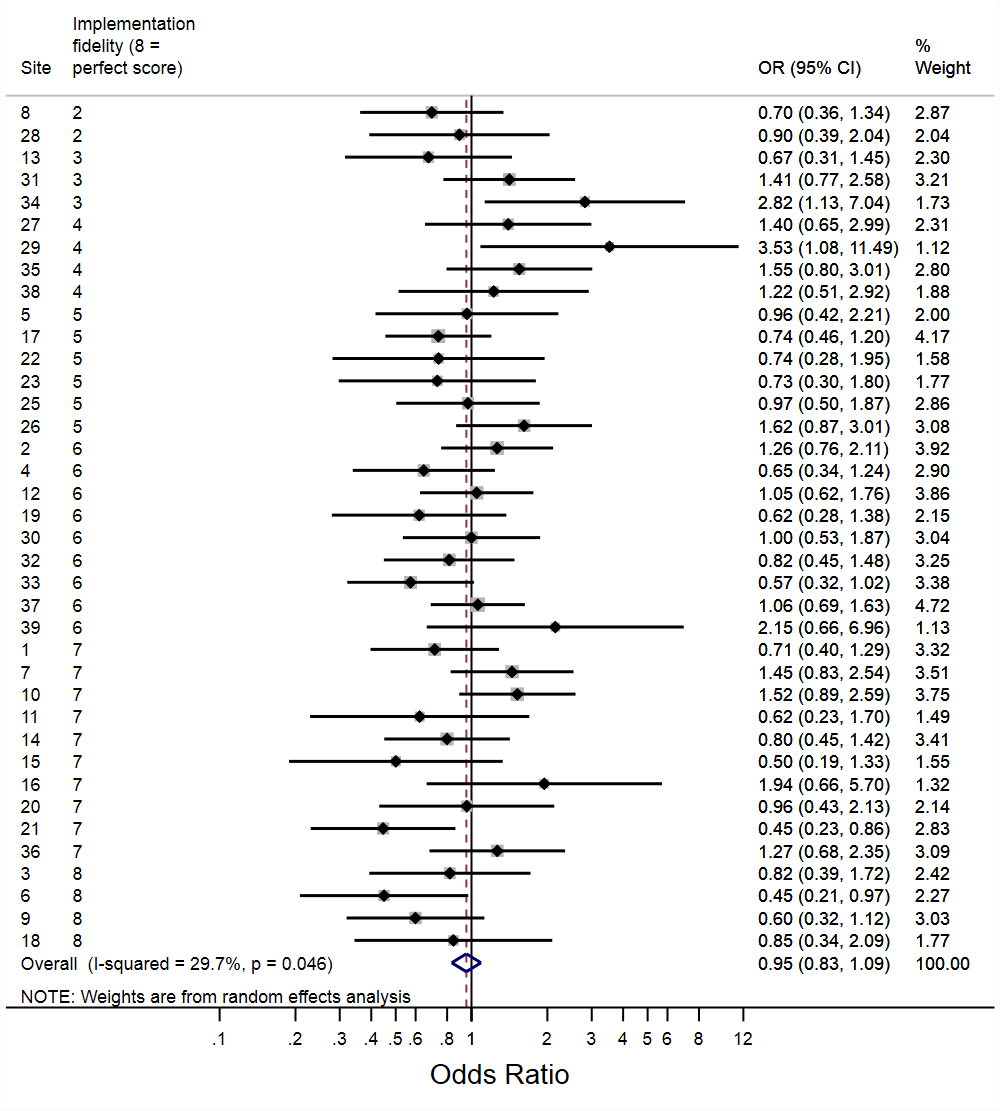 | **B** | ***C difficile* infection: Change in trend post vs pre-implementation**  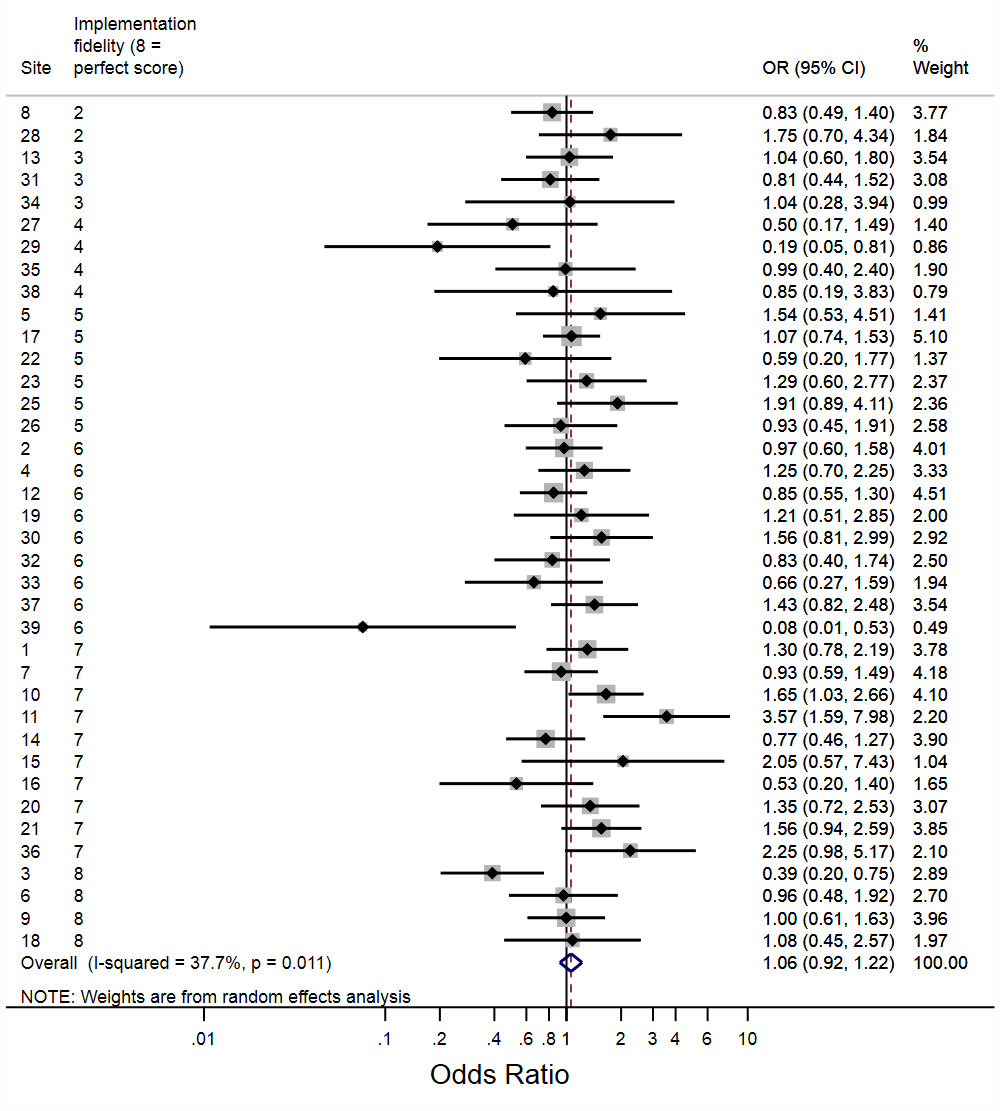 |
| --- | --- | --- | --- |
| **C** | ***C difficile* colonisation: Immediate implementation effect**  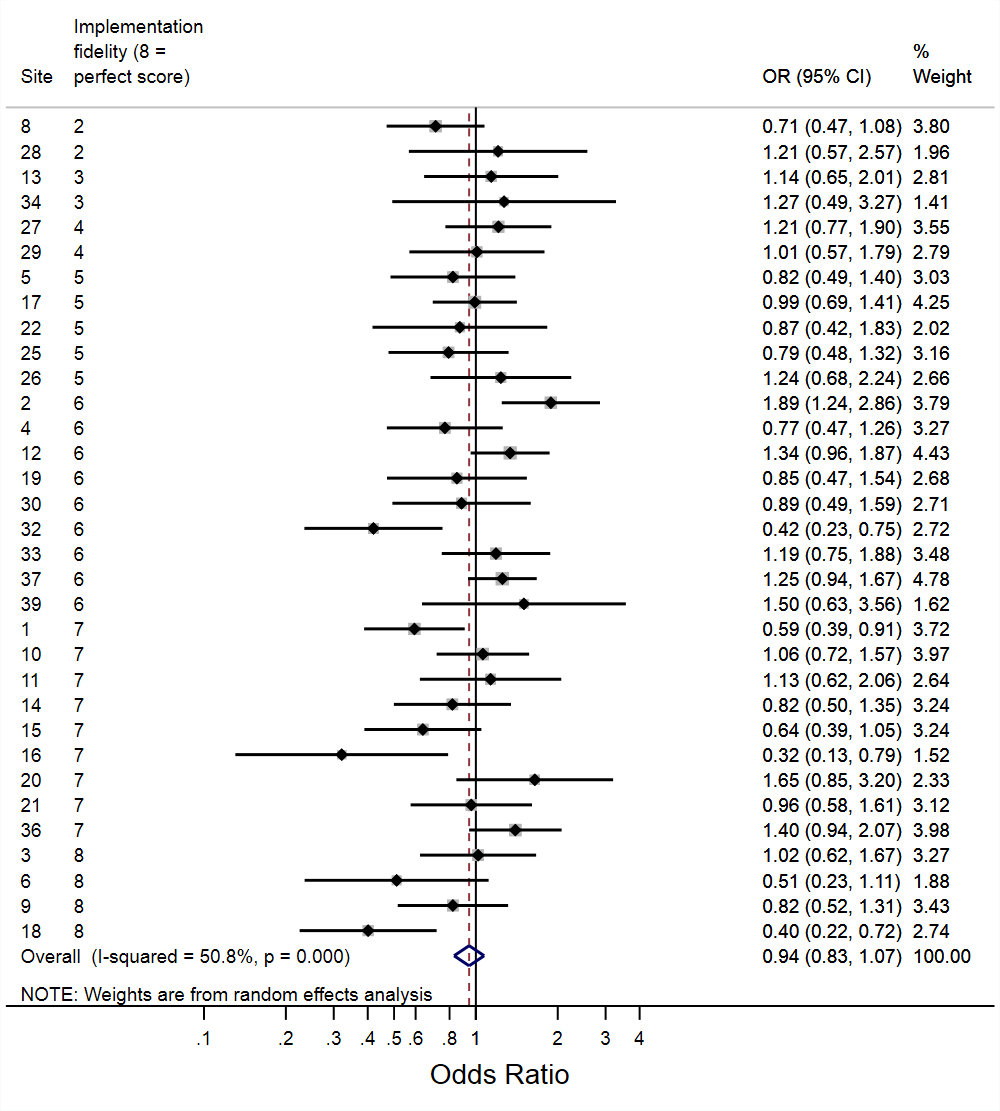 | **D** | ***C difficile* colonisation: Change in trend post vs pre-implementation**  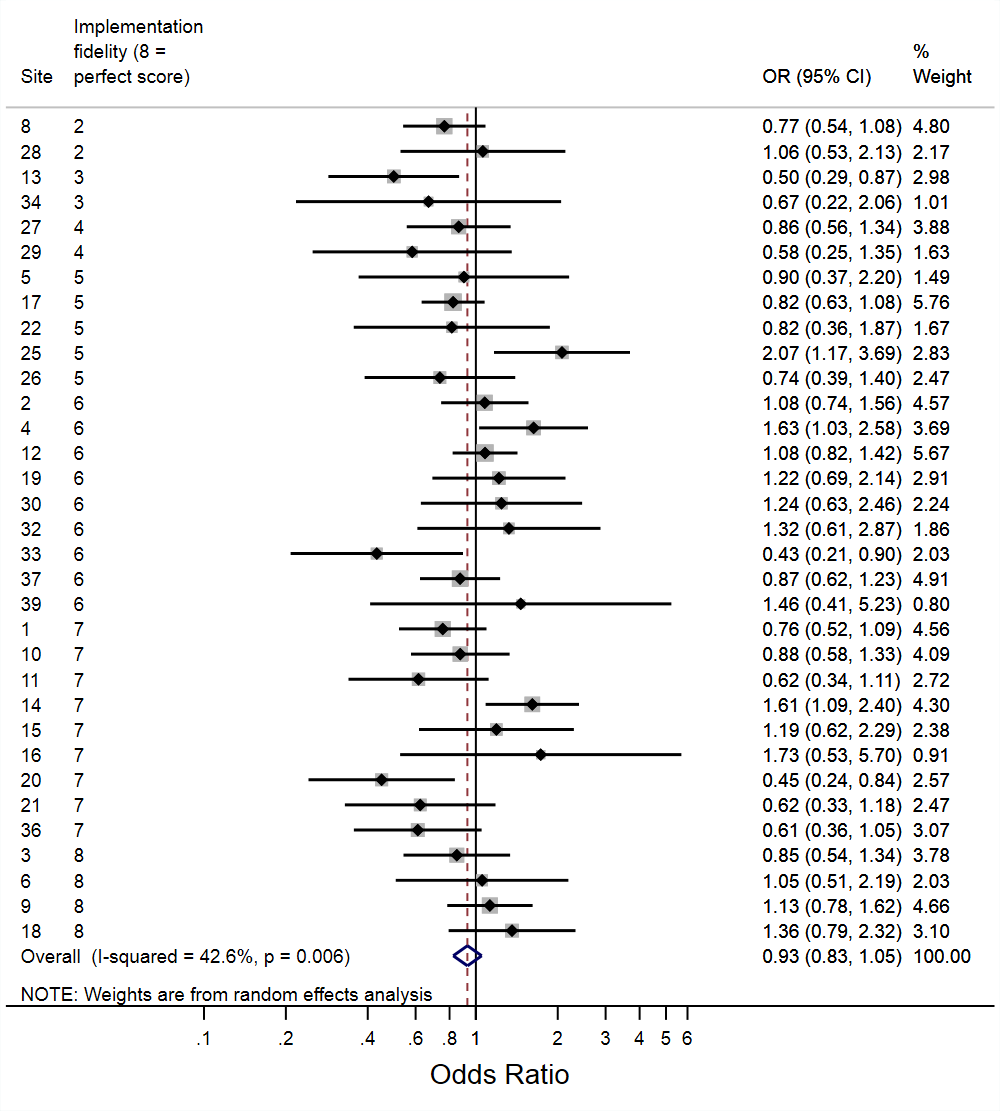 |
